# Exposure to violence, chronic stress, nasal DNA methylation, and atopic asthma in children

**DOI:** 10.1101/2020.11.03.20225250

**Authors:** Qi Yan, Erick Forno, Andres Cardenas, Cancan Qi, Yueh-Ying Han, Edna Acosta-Pérez, Soyeon Kim, Rong Zhang, Nadia Boutaoui, Glorisa Canino, Judith M. Vonk, Cheng-jian Xu, Wei Chen, Emily Oken, Diane R. Gold, Gerard H. Koppelman, Juan C. Celedón

## Abstract

**Background:** Exposure to violence (ETV) or stress may cause asthma through unclear mechanisms.

**Methods:** Epigenome-wide association study (EWAS) of DNA methylation in nasal epithelium and four ETV or chronic stress measures in 487 Puerto Ricans aged 9-20 years who participated in the Epigenetic Variation and Childhood Asthma in Puerto Ricans study [EVA-PR]). We assessed measures of ETV or chronic stress in children (ETV scale, gun violence, and perceived stress) and their mothers (perceived stress). Each EWAS was conducted using linear regression, with CpGs as dependent variables and the stress/violence measure as a predictor, adjusting for age, sex, the top five principal components, and SVA latent factors. We then selected the top 100 CpGs (by P-value) associated with each stress/violence measure in EVA-PR and conducted a meta-analysis of the selected CpGs and atopic asthma using data from EVA-PR and two additional cohorts (Project Viva and PIAMA).

**Results:** In the EWAS of stress/violence in EVA-PR, gun violence was associated with methylation of cg18961589 in *LINC01164* (β=0.03, *P*=1.28×10^−7^), and maternal stress was associated with methylation of cg03402351 in *SNN* (β=0.04, *P*=1.69×10^−7^) and cg19064846 in *PTPRN2* (β=0.03, *P*=3.36×10^−7^). In a meta-analysis of three cohorts, which included the top CpGs associated with stress/violence in EVA-PR, CpGs in *STARD3NL, SLC35F4, TSR3, CDC42SE2, KLHL25, PLCB1, BUD13, OR2B3, GALR1, TMEM196, TEAD4* and *ANAPC13* were associated with atopic asthma at FDR-*P* < 0.05.

**Conclusions:** ETV and chronic stress may increase the risk of atopic asthma through DNA methylation in airway epithelium, though this needs confirmation in future longitudinal studies.

## INTRODUCTION

The U.S. has the highest rate of firearm-related deaths of all industrialized countries ^1^. Exposure to violence (ETV), including gun violence, affects stress responses and may influence asthma pathogenesis ^2–4^. Prolonged ETV can lead to chronic and frequent activation of the body’s stress responses, disrupting neuroendocrine, autonomic nervous system, and immune responses. Chronic stress, both related and unrelated to ETV, may cause or worsen asthma by altering circulating levels of and response to catecholamines and glucocorticoids, as well as by affecting immune responses, leading to airway inflammation and airflow obstruction ^2^. ETV and chronic stress may also cause or worsen asthma through indirect effects mediated by other risk factors. For example, community violence may decrease outdoor physical activity, leading to obesity ^3^.

Puerto Ricans are often exposed to violence in their communities ^5–7^. ETV is associated with asthma in Puerto Rican children ^7^, in whom ETV is also associated with DNA methylation of the promoter of the gene for the pituitary adenylate cyclase activating polypeptide 1 type 1 receptor (*ADCYAP1R1*) in white blood cells. Moreover, such methylation has been associated with childhood asthma in Puerto Ricans.

Based on prior findings, we hypothesized that both ETV and chronic stress would increase asthma risk through DNA methylation of genes regulating autonomic, neuroendocrine and immune responses in airway epithelium, a tissue relevant to asthma. To examine this hypothesis, we first tested for association between ETV or chronic stress measures and genome-wide DNA methylation in nasal epithelium, as DNA methylation in nasal (airway) epithelium is well correlated with that in bronchial (airway) epithelium ^8–11^, and epigenetic regulation of airway epithelial gene expression has been implicated in asthma pathogenesis ^12^. We then examined whether the top CpGs associated with ETV or chronic stress were also associated with atopic asthma in Puerto Rican children and in two independent cohorts. We focused on atopic asthma, both because atopic asthma is the most common type of childhood asthma and because we previously showed that DNA methylation profiles can accurately classify children according to the presence of atopic asthma ^12^.

## METHODS

### Study population

Subject recruitment and study procedures for the Epigenetic Variation and Childhood Asthma in Puerto Ricans study (EVA-PR) have been previously described ^12^. In brief, EVA-PR is a case-control study of asthma in subjects aged 9 to 20 years. Participants with and without asthma were recruited from households in San Juan (PR) from February 2014 through May 2017, using multistage probability sampling; 638 households had ≥1 eligible child, and 543 (85.1%) children (one per household) agreed to participate. The study was approved by the institutional review boards of the University of Puerto Rico (San Juan, PR) and the University of Pittsburgh (Pittsburgh, PA). Written parental consent and assent were obtained from participants <18 years old, and consent was obtained from participants ≥18 years old.

The study protocol included administration of questionnaires on ETV and chronic stress in the child and her/his mother, measurement of serum allergen-specific IgEs, and collection of nasal epithelial samples for DNA and RNA extraction. Participating children completed a modified version of the ETV Scale ^13–15^, a questionnaire used to assess ETV in children ≥8 years. The ETV Scale measures both witnessing and direct victimization for five events: shoving, punching, or kicking; knife attacks; shootings; hearing gunshots; and witnessing verbal abuse of the child’s primary caregiver. An ETV score (range=0-15) is obtained by summing the scores for each section. Internal consistency and test-retest reliability and validity have been established for the English and Spanish versions. The child’s exposure to gun violence was derived from the ETV scale ^16^. Participating children also completed the Checklist of Children’s Distress Symptoms (CCDS), a 28-item scale to assess stress symptoms in the previous six months as a result of ETV ^14, 17^. Answers for each question in the CCDS range from 1 to 5. An overall score is obtained by summing the scores for all 28 questions, and then dividing by the number of questions, so that the score ranges between 1 and 5. Mothers of participating children completed the perceived stress scale (PSS) questionnaire, which includes 4 items and measures the degree to which mothers believed that their lives were unpredictable, uncontrollable, or overwhelming in the preceding month ^18^. Each question in the maternal PSS ranges between 0 and 4, and a total score ranges between 0 and 16 ^18^.

Serum IgEs to each of five common allergens in Puerto Rico (dust mite [Der p 1], cockroach [Bla g 2], cat dander [Fel d 1], dog dander [Can f 1], and mouse urinary protein [Mus m 1]) were measured using the UniCAP 100 system (Pharmacia & Upjohn, Kalamazoo, Mich). Atopy was defined as an IgE ≥0.35 IU/mL to ≥1 of the allergens tested. Asthma was defined as a physician’s diagnosis plus ≥1 episode of wheeze in the previous year. Atopic asthma was defined as the presence of both atopy and asthma.

Whole-genome methylation in nasal epithelium was measured using Infinium HumanMethylation450 BeadChip arrays (Illumina, San Diego, CA). Methylation β-values were calculated as a percentage: β=M/(M+U+α), where M and U represent methylated and unmethylated signal intensities, respectively, and α is an arbitrary offset to stabilize β-values where fluorescent intensities are low. The β-values were used in all downstream analyses. We performed the same preprocessing and quality control procedures as in our previous study ^12^. In brief, Raw IDAT files were loaded using the R package *minfi* ^19^. Samples with low detection values (>10% CpG probes with a detection *P* >0.01) were removed. The R package *ENmix* ^*20*^ was used to perform background correction and normalization. Cross-reactive (n=26,098) and SNP-containing probes ^21^ (n=15,834), sex chromosomal probes (n=9,490), and low-quality probes (>10% of samples with detection *P* >0.01; n=21,174) were also removed. After further filtering, CpG sites with an overall mean β-value of greater than 0.9 or less than 0.1 were removed, leaving 227,901 CpG probes in the analysis.

RNA-Seq was performed using the Illumina NextSeq 500 platform, with paired-end reads at 75 cycles and 80 million reads per sample; reads were aligned to reference human genome (hg19) ^22^ and TPM (Transcripts Per Kilobase Million) were used as proxy for gene expression level. After QC, 16,737 genes were retained for the analyses of gene expression, which focused on genes in *cis* (within 1 Mb on each side of the transcription start site) with the top CpGs from the methylation analyses. Genome-wide genotyping was conducted using the HumanOmni2.5 BeadChip platform, (Illumina, San Diego, CA), as previously described.

### Study cohorts included in the meta-analysis of atopic asthma

#### PIAMA

PIAMA is a birth cohort study of children born in the Netherlands in 1996 and 1997. The study protocol has been previously described ^23^. The Medical Ethical Committees of the participating institutions approved the study, and the parents and legal guardians of all participants, as well as the participants themselves, gave written informed consent. At age 16 years, nasal epithelial cells were collected at two study centers (Groningen and Utrecht) ^24^ by brushing the lateral area underneath the right inferior turbinate. Serum IgEs to each of four common allergens (house dust mite, cat, dactylis (grass) and birch) were measured, and atopy was defined as an IgE ≥0.35 IU/mL to any of the allergens tested. Asthma was defined as physician-diagnosed asthma and either wheeze in the previous year or use of medications for respiratory or lung problems. Atopic asthma was defined as the presence of both asthma and atopy.

In total, 479 nasal epithelial samples were hybridized to the Infinium HumanMethylation450 BeadChip arrays. DNA methylation data were pre-processed with Bioconductor package *minfi* ^19^. Samples with call rate <99% were removed. 65 SNP probes were used to check for concordance between paired DNA samples (nasal and blood DNA samples from the same subjects were hybridized in the same experiments); paired samples with Pearson correlation coefficient <0.9 were excluded, as were probes on sex chromosomes, probes that mapped to multiple loci, 65 SNP-probes, and probes containing SNPs at the target CpG sites with a MAF>5% ^21^. “DASEN” ^25^ was used to perform signal correction and normalization. After QC, 455 samples and 436,824 probes remained in the analysis, as previously described ^26^. Of these 455 samples, 246 samples corresponded to subjects with atopic asthma and non-atopic control subjects, and were thus included in the meta-analysis.

#### Project Viva

Project Viva is a prospective pre-birth cohort study of mother-child pairs recruited between 1999 and 2002 during the mothers’ first prenatal visits at Atrius Harvard Vanguard Medical Associates, a medical practice in eastern Massachusetts (U.S.) ^27^. The Institutional Review Board of Harvard Pilgrim Health Care reviewed and approved all study protocols. Asthma and atopy were assessed at the same time-point as collection of nasal samples. Atopy was defined as ≥1 IgE ≥0.35 IU/mL to the allergens tested (Dermatophagoides farinae, cat dander, dog dander, Alternaria or Aspergillus species, rye grass, ragweed, and oak and silver birch).

Nasal swabs were collected from the anterior nares, previously demonstrated to yield respiratory epithelial cells ^28^. Epigenome-wide methylation was measured on DNA extracted from nasal samples, using Infinium MethylationEPIC BeadChip arrays (Illumina). DNA methylation data was imported into the *R* statistical software for preprocessing using *minfi* ^19^. QC was first performed at the sample level, excluding samples that: had overall low intensity (<10.5; n=3), were mismatched on recorded sex (n=4), or had mixed genotype distributions on the measured SNP probes (n=59) indicating possible sample contamination (n=8). In addition, 35 technical duplicates were excluded. Of 547 high-quality samples, 156 corresponded to subjects with atopic asthma and non-atopic controls and were thus retained for the meta-analysis. QC was then performed at the probe level by computing a detection *P*-value relative to control probes. In total, 40,377 cross-reactive probes previously identified in the MethylationEPIC BeadChip ^29^ were excluded, leaving 719,075 high-quality probes included in the analysis. DNA methylation heterogeneity was estimated by including potential cell-type variation, using 10 principal components derived via ReFACTor ^30^.

### Statistical analysis

We analyzed four measures of ETV and chronic stress. As in prior work, the child’s ETV scale was analyzed as continuous, the child’s exposure to gun violence was analyzed as binary (having heard gunshots at least twice vs. no more than once) ^16^, the child’s perceived stress scale was analyzed as binary (upper quartile [≥2.6 points] vs. lowest three quartiles [<2.6 points]) ^31^, and maternal perceived stress (assessed by the PSS) was analyzed as binary (upper quartile [>7 points] vs. lowest three quartiles [≤7 points]) ^31^. Of the 487 participants in EVA-PR who had nasal genome-wide methylation (GWM) data and complete data on relevant covariates, 470 had ETV data, 475 had data on gun violence, 471 had CCDS data, and 476 had maternal PSS data.

We conducted epigenome-wide association studies (EWAS) of ETV or chronic stress measures among subjects in EVA-PR using linear regression models, as follows:

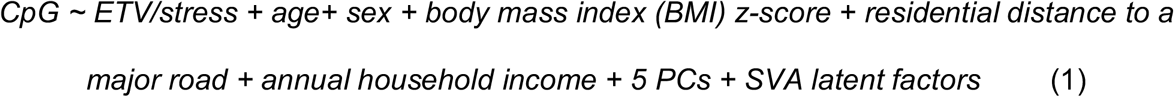

where CpG is the β-value for a CpG methylation, with known batches adjusted by R function *Combat* ^*32*^, BMI z-score is calculated using CDC growth charts, residential distance to a road (a proxy for traffic-related air pollution) is a binary covariate (as in prior work, < 444.7 vs. ≥ 444.7 meters or quartiles 1-3 vs. quartile 4), annual household income is a binary covariate (≥ $15,000 vs. < $15,000 per year, near the median household income in Puerto Rico in 2008-2009), 5 PCs are the top five principal components from genome-wide genotypic data, and latent factors (LFs) are estimated by *sva* ^33^ (which capture unknown data heterogeneity).

As determined *a priori* (because of our sample size and limited statistical power in EVA-PR), we selected the top 100 CpG sites from each of the four EWAS (one for each measure of ETV or chronic stress) for the analysis of atopic asthma. For this analysis in EVA-PR, we included 272 participants (167 subjects with atopic asthma and 105 non-atopic controls). Next, we used linear regression modeling to test whether the top methylation signals for ETV or chronic stress were also associated with atopic asthma in each of the three study cohorts (EVA-PR, Viva, and PIAMA), as follows:

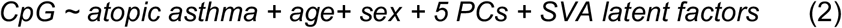

We then conducted a meta-analysis of the three EWAS of atopic asthma. In this meta-analysis, P-values from each of the three independent studies were taken as input, with effect direction considered. First, a Z-score was calculated based on the P-value and direction of effect in each study, as follows:

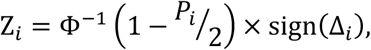

where Z _*i*_ is the Z-score for study *i, P*_*i*_ is the P-value for study *i*, Δ_*i*_ is the direction of effect for the study *i*, and Φ^−1^ gives the percentile of a standard normal distribution. Combined coefficients were calculated by averaging the study-specific coefficients, with weights reflecting the standard errors (SE) from each study, as follows:

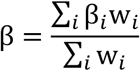

where β is the combined coefficient, β _*i*_ is the study-specific coefficient, and 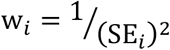 is the weight for study *i*. An overall P-value was then calculated as:

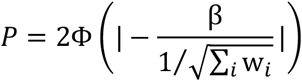

where *P* is the overall *P*-value. The false discovery rate (FDR) approach was used to adjust for multiple testing.

To assess the biological relevance of the top CpG sites for atopic asthma, we conducted an expression quantitative trait methylation (eQTM) analysis, to test whether those CpGs affected the expression of *cis*-genes (i.e. those with transcription start sites located within 1 Mb of the CpG, on each side) in EVA-PR. This analysis was adjusted for age, sex, asthma and atopy status, the top five PCs, RNA sample sorting protocol (i.e., whole-cells or CD326-positive nasal epithelial cells) ^37^, methylation and RNA-Seq batch, and LFs estimated from *sva* ^33^.

## RESULTS

The main characteristics of the EVA-PR participants included in the four EWAS of ETV and stress measures (ETV, gun violence, CCDS and PSS) are shown in **Supplementary Table S1**. In reviewing the results for each of these EWAS, the Q-Q plots showed no evidence of inflation (**Figure 1**). In these EWAS, methylation of cg18961589 in *LINC01164* (on chromosome 10) was associated with gun violence exposure (β=0.03, *P*=1.28×10^−7^ at FDR*-P* < 0.05), and methylation of cg03402351 in *SNN* (on chromosome 16, β=0.04, *P*=1.69×10^−7^) and cg19064846 in *PTPRN2* (on chromosome 7, β=0.03, *P*=3.36×10^−7^) were associated with maternal perceived stress at FDR*-P* < 0.05 (**Figure 2**).

**Figure 1.**
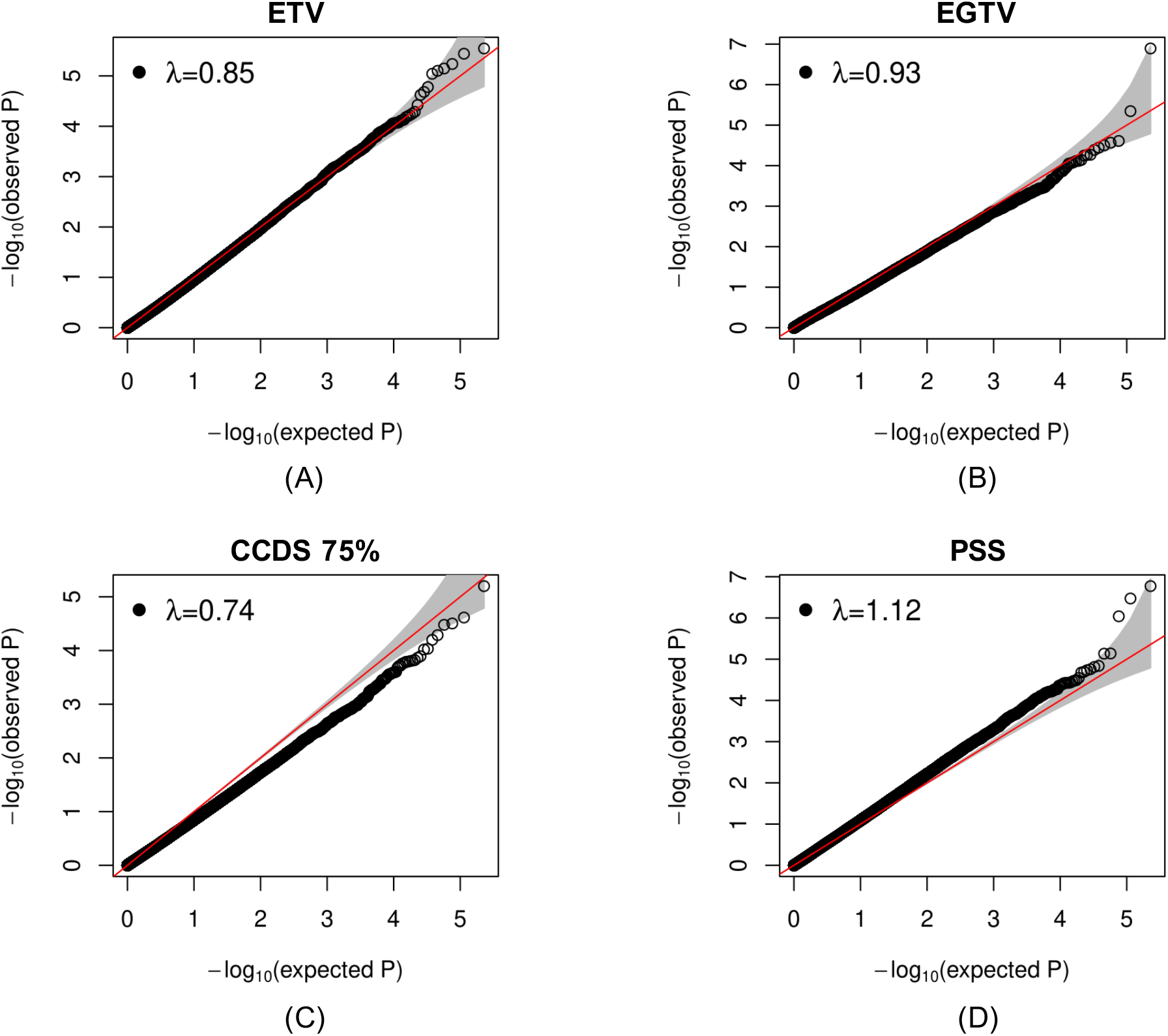
Q-Q plots of four epigenome-wide analyses of stress/violence measures in nasal epithelium from participants in the Epigenetic Variation of Childhood Asthma in Puerto Ricans study (EVA-PR). ETV: Exposure to violence. ETGV: Exposure to gun violence. CCDS: Checklist of Children’s Distress Symptoms. PSS: Perceived Stress Scale.

**Figure 2.**
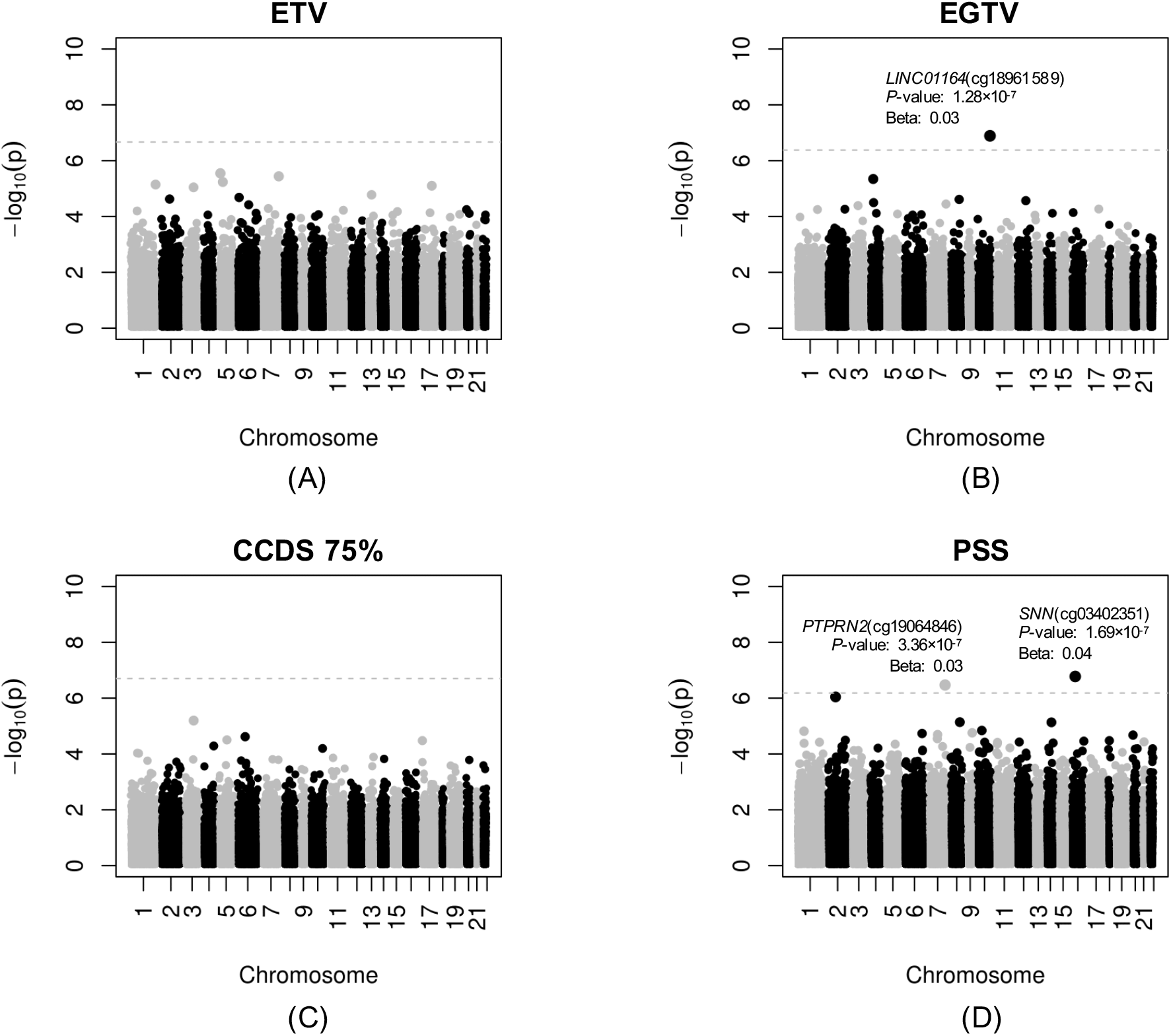
Manhattan plots for the epigenome-wide association studies (EWAS) of four ETV/stress measures in nasal epithelial samples from participants in the Epigenetic Variation of Childhood Asthma in Puerto Ricans study (EVA-PR). (A) Exposure to Violence scale (ETV), (B) Exposure to gun violence (ETGV), (C) Checklist of Child Distress Symptoms (CCDS), and (D) maternal perceived stress symptoms (PSS). The chromosomal position of each CpG site is displayed along the X-axis and the negative logarithm of the association *P*-value is displayed on the Y-axis. The red line represents the threshold for genome-wide significance line (false-discovery rate-adjusted *P* < 0.05).

We then selected the top 100 CpG sites (by P-value) from each of the four EWAS in EVA-PR for testing for association with atopic asthma in a meta-analysis including data from EVA-PR, PIAMA, and Project Viva. A total of 336 CpG sites were included in this meta-analysis, as 4 CpG sites overlapped across EWAS and 60 CpG sites were missing in Project Viva; see **Supplementary Tables S2-S3**). The main characteristics of participants in the three cohorts included in the meta-analysis of atopic asthma and nasal epithelial DNA methylation are shown in **Table 1**. As expected, EVA-PR (a case-control study of asthma) had a larger proportion of participants with atopic asthma than PIAMA or Viva (both unselected for asthma). The studies were conducted in different geographic locations, ranging from Puerto Rico to Boston to the Netherlands. While all participants in EVA-PR were Puerto Rican and most participants in PIAMA were European, Viva was ethnically diverse.

**Table 1.**
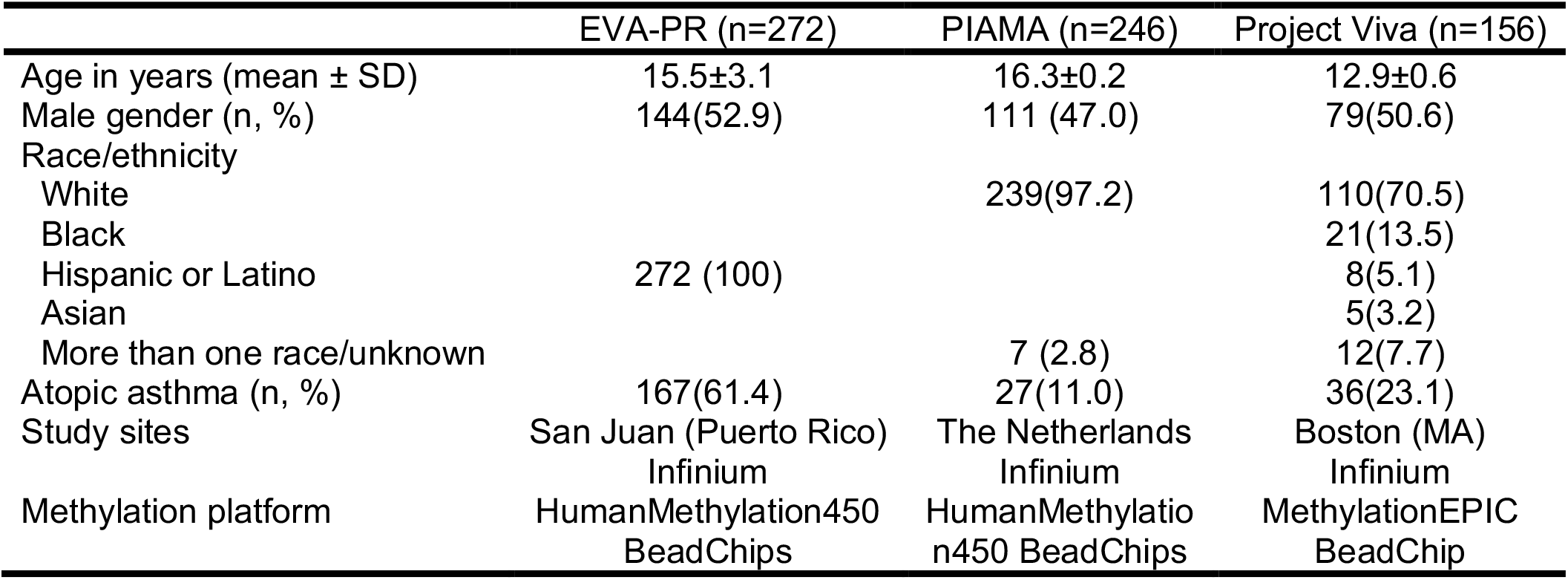
Summary of characteristics of participants in the studies included in the meta-analysis of CpG sites (associated with stress or violence measures in EVA-PR) and atopic asthma.

We first performed separate analyses of the 336 CpG sites and atopic asthma in each of the three study cohorts (EVA-PR, PIAMA and Viva), and then combined the results from the three cohorts in a meta-analysis. In this meta-analysis, we identified 7 CpGs (cg02695349 in *STARD3NL*, cg18146152 in *TSR3*, cg03541903 in *CDC42SE2*, cg21376795 in *KLHL25*, cg26772788 in *GALR1*, cg03729152 in *TMEM196* and cg16848072 in *ANAPC13*) associated with higher odds of atopic asthma, and 5 CpGs (cg04990977 in *SLC35F4*, cg27178677 in *PLCB1*, cg17076485 in *BUD13*, cg17335499 in *OR2B3*, and cg01039401 in *TEAD4*) associated with lower odds of atopic asthma, all at FDR-*P* < 0.05 (**Table 2**).

**Table 2.**
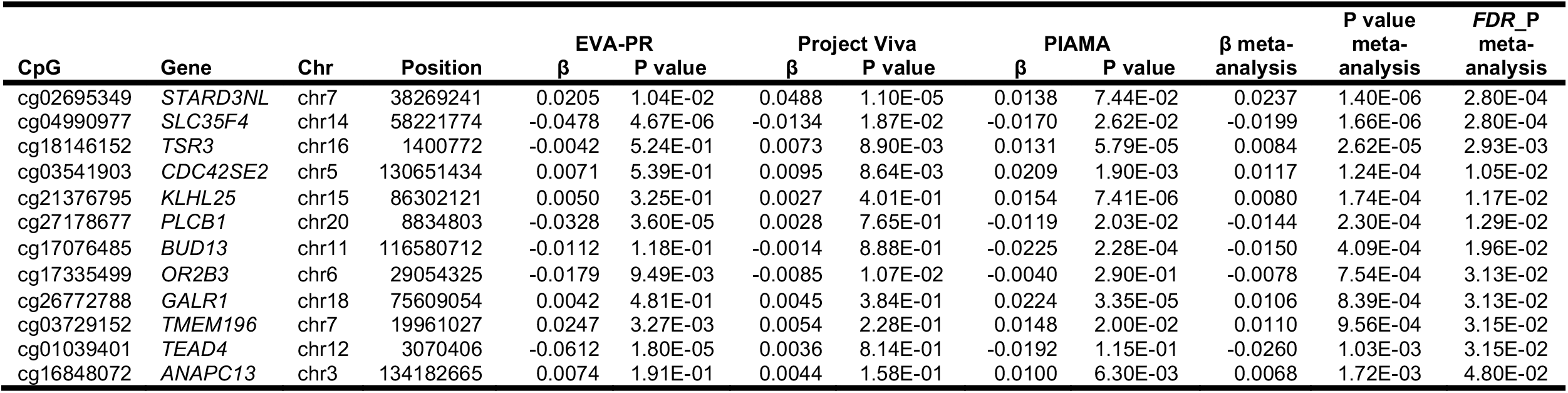
CpG sites associated with atopic asthma (at *FDR-P* < 0.05) in a meta-analysis of data from three cohorts (EVA-PR, Project Viva, and PIAMA)

We then conducted a *cis*-expression quantitative trait methylation (*cis*-eQTM) analysis of the 12 CpGs associated with atopic asthma in nasal epithelial cells from participants in EVA-PR. In this analysis, we identified 16 nominally significant *cis*-acting eQTM probes in nasal epithelium (**Supplementary Table 4**).

Our EWAS of ETV or stress measures in EVA-PR included children with and without asthma. We did not adjust for asthma status in these analyses, as this could have attenuated our effect estimates for the analysis of atopic asthma. In a sensitivity analysis, we repeated the four EWAS of ETV or stress measures after additional adjustment for asthma status, obtaining similar results (**Supplementary Figures S1 and S2**). Moreover, when the top 100 CpGs from each of these four EWAS (adjusted for asthma status) were tested for association with atopic asthma in a meta-analysis of the three study cohorts, 7 of the 12 CpGs from the primary analysis were also significant at FDR-*P* <0.05 (in *STARD3NL, TSR3, CDC42SE2, PLCB1, OR2B3, GALR1* and *TMEM196*).

## DISCUSSION

ETV and chronic stress have been linked to childhood asthma ^34^. We previously showed that ETV and gun violence are associated with childhood asthma in Puerto Ricans, a high-risk group ^4, 7, 35^. To our knowledge, this is the first report of an association between chronic stress or ETV and DNA methylation in nasal epithelium, as well as the first association study of stress- or violence-related methylation markers in nasal epithelium and atopic asthma.

Among Puerto Rican children, exposure to gun violence was significantly associated with increased methylation of a CpG site (cg18961589) in the gene for long intergenic non-protein coding 1164 (*LINC01164). LINC01164* is most highly expressed in brain tissue from GTEx ^36^ (**Figure 3**), and SNPs in this gene have been associated with cognitive function and educational attainment ^37^. In our recent EWAS of atopic asthma, methylation of another CpG site in *LINC01164* (cg15491439) was significantly associated with atopic asthma in EVA-PR (FDR-*P* = 4.20×10^−4^) ^12^. Moreover, this long non-coding RNA is adjacent (∼128Kbp) to *PPP2R2D*, a gene that codes for a regulatory subunit of the PPP2A phosphatase family that has been linked to T-cell proliferation and apoptosis in mice ^38^.

**Figure 3.**
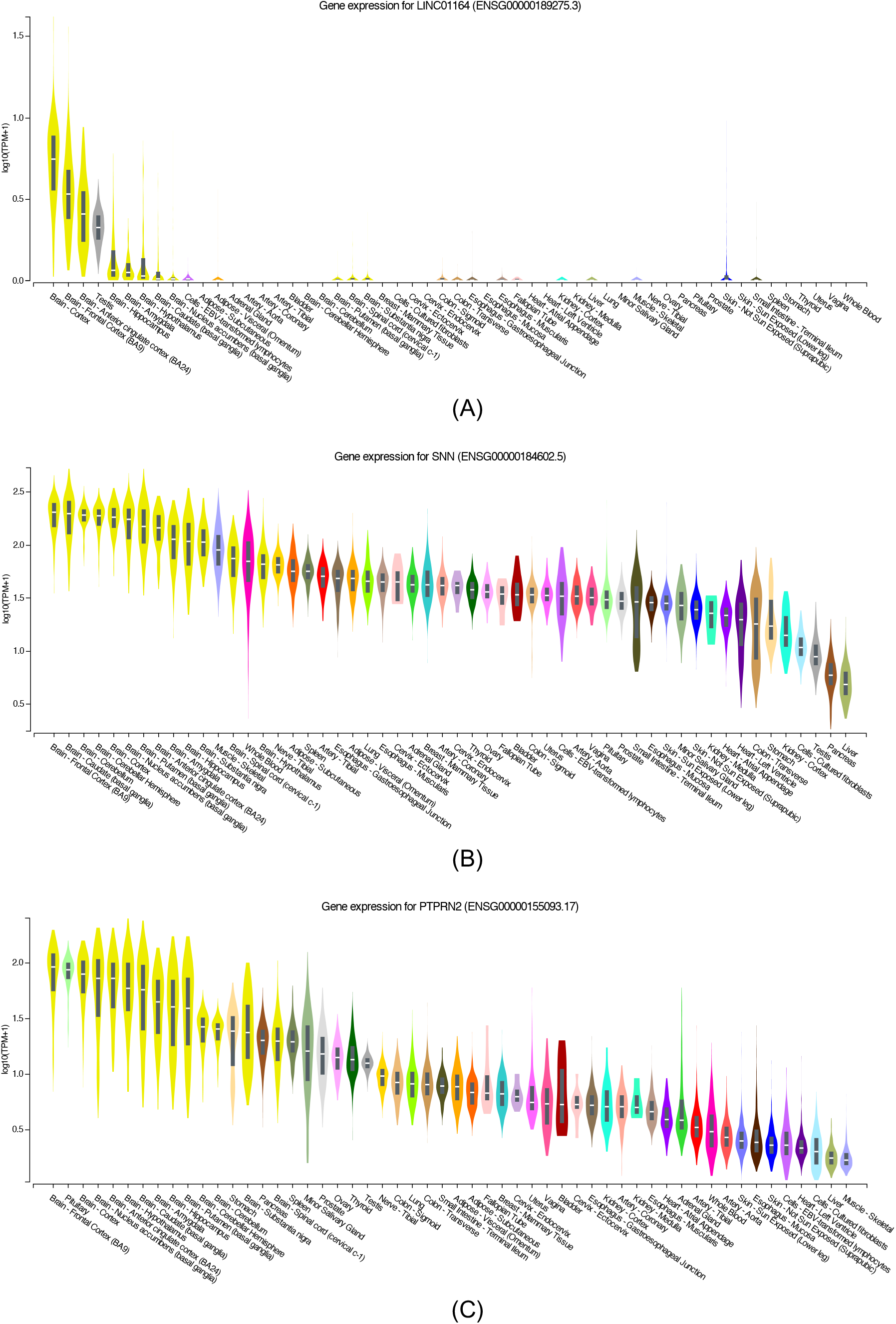
*LINC01164, SNN* and *PTPRN2* expression in different tissues from GTEx Increased methylation of cg03402351 in *SNN* (stannin) and increased methylation of cg19064846 in *PTPRN2* (receptor-type tyrosine-protein phosphatase N2) in nasal epithelium were associated with maternal perceived stress. Both *SNN* and *PTPRN2* are highly expressed in brain tissue from GTEx ^36^ (**Figure 3**). Stannin is a marker for neuronal cell apoptosis induced by Trimethyltin intoxication, where Trimethyltin is known to inflict injury to specific regions of the brain ^39^. *PTPRN2* methylation and expression have been associated with Parkinson’s Disease ^40–42^, and a meta-analysis of epigenome-wide data showed that a differentially methylated region in *PTPRN2* was associated with childhood asthma ^43^.

In the meta-analysis of selected stress- or violence-associated CpG sites in the study cohorts, we identified 12 CpG sites associated with atopic asthma (in genes *STARD3NL, TSR3, CDC42SE2, KLHL25, GALR1, TMEM196, ANAPC13, SLC35F4, PLCB1, BUD13, OR2B3*, and *TEAD4*). Of note, expression levels of *STARD3NL* (log2FC [fold-change] = −0.16; FDR-*P* = 6.24×10^−6^), *TSR3* (log2FC=0.07; FDR-*P*=0.04), *CDC42SE2* (log2FC= −0.18; FDR-*P*=2.62×10^−4^), *KLHL25* (log2FC=0.21; FDR-*P*=3.11×10^−6^), *PLCB1* (log2FC= −0.31; FDR-*P*=0.01), and *ANAPC13* (log2FC = −0.13; FDR-*P*=1.27×10^−3^) were associated with atopic asthma in our recent transcriptome-wide association study using nasal epithelial samples from Puerto Ricans ^44^.

*PLCB1* has been associated with bronchodilator response in children with asthma ^45^, and is differentially expressed in children with treatment-resistant asthma compared with children with controlled persistent asthma or healthy controls ^46^. Silencing *PLCB1* can attenuate lipopolysaccharide-induced endothelial cell inflammation by inhibiting expression of proinflammatory cytokines ^47^. In the brain, *PLCB1* is expressed mainly in the cortex, hippocampus and amygdala; the protein it codes for, PLCB1, is a mediator of synaptic plasticity that plays an important role in cognitive behavior and emotions ^48^ and may influence the pathogenesis of stress-related disorders. Indeed, expression of *PLCB1* is decreased in the frontal cortex of rats subjected to chronic stress, and this downregulation is reversed by quetiapine, a drug used to treat major depression ^49^. *PLCB1* has also been implicated in depression, bipolar disorder, epilepsy, and schizophrenia ^50^.

Four SNPs in *TMEM196* have been shown to be associated with decreased odds of nonsteroidal anti-inflammatory drug (NSAID)-exacerbated respiratory disease (NERD) in asthma ^51^. Moreover, *TMEM196* has been shown to regulate autophagy and apoptosis of cancer and inflammatory cells ^52^. Of note, disruption of the balance between autophagy and apoptosis induces hyperplasia of the airways, a typical feature of nasal polyps and airway eosinophilia in NERD ^53–55^.

We recognize several study limitations. First, we had limited statistical power to examine ETV or chronic stress and DNA methylation, as only one cohort (EVA-PR) had such data. Although top methylation markers in the EWAS of stress/violence measures were associated with atopic asthma in the meta-analysis of three study cohorts, none achieved genome-wide significance for ETV or stress measures in EVA-PR, and thus this needs follow-up in future studies. Second, we cannot assess temporal relationships in this cross-sectional study. Third, covariates correlated with stress or violence may partly explain our results. However, our EWAS of stress or violence measures in EVA-PR was adjusted for indicators of socioeconomic status (annual income), adiposity (BMI), and traffic-related air pollution. Moreover, some of the top genes associated with atopic asthma have been implicated in neuropsychiatric function or disorders (i.e. *PLCB1)*.

In summary, we show that measures of chronic stress or violence are associated with nasal DNA methylation markers in Puerto Rican youth. Moreover, we show that some stress- or violence-related methylation markers are also associated with atopic asthma in a meta-analysis of data from three cohorts of youth in diverse racial/ethnic groups. Although our findings are preliminary and await confirmation in other studies, they suggest that chronic stress or violence may alter DNA methylation in nasal epithelium, and that stress- or violence-linked methylation markers may increase the risk of atopic asthma.

## Data Availability

The DNA methylation data supporting the conclusions of this manuscript will be made available by the authors upon request to any qualified researcher, pending ethical approval.

## Acknowledgments

We thank all the participating children and families.

