## Supplemental Tables and Figures for "Exposure to violence, chronic stress, nasal DNA methylation, and atopic asthma in children"

**Figure S1. Manhattan plots of epigenome-wide association analyses (EWAS) of four ETV/stress EWAS results with additional adjustment for asthma status in model (1).**

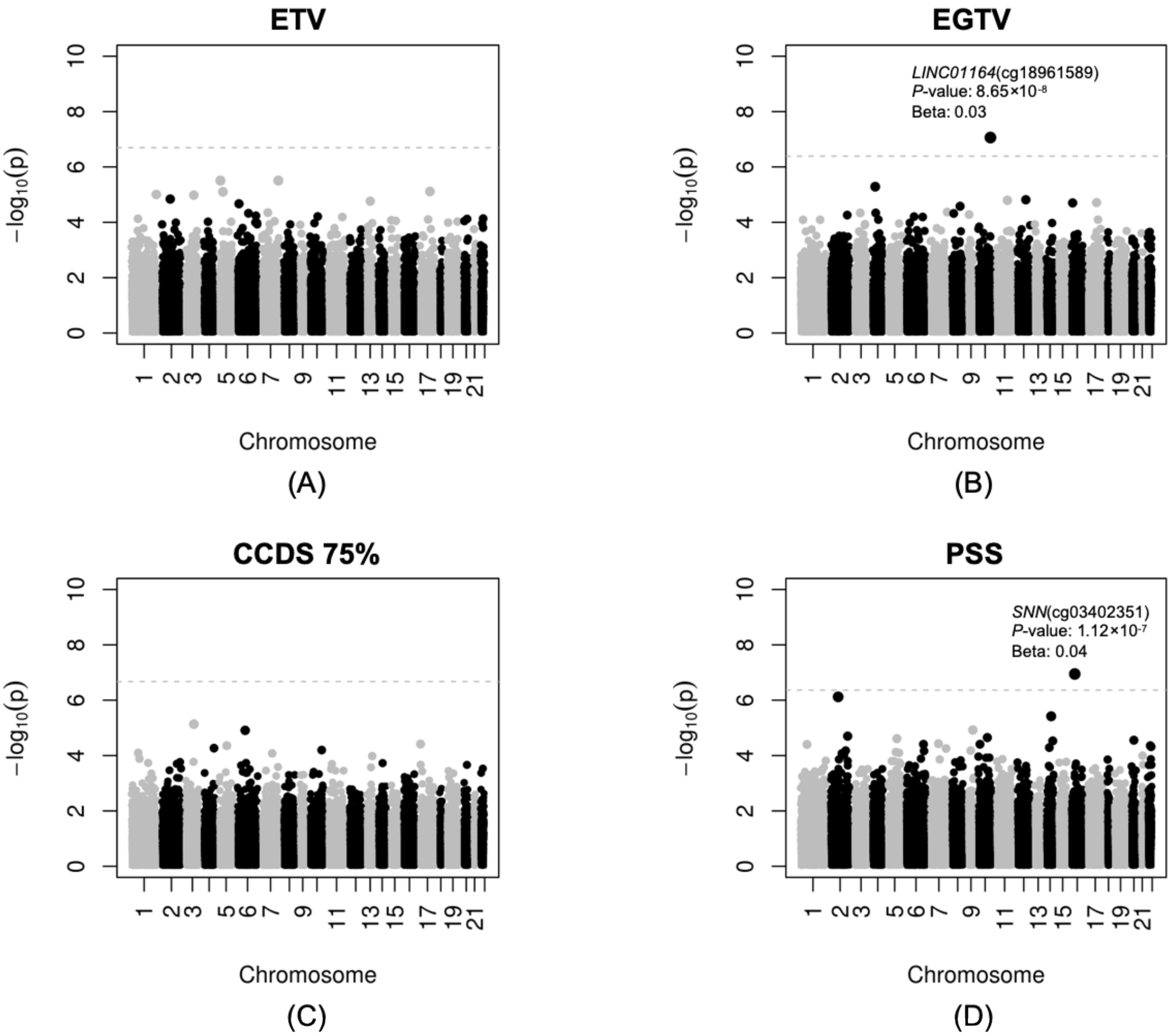


Manhattan plots showing the EWAS results of (A) ETV, (B) ETGV, (C) CCDS, and (D) PSS in EVA-PR nasal epithelial cells. The chromosomal position of each CpG site is displayed along the X-axis and the negative logarithm of the association P-value is displayed on the Y-axis. The red line represents the genome-wide significance line (FDR < 0.05). ETV: Exposure to violence. ETGV: Exposure to gun violence. CCDS: Checklist of Children's Distress Symptoms. PSS: Perceived Stress Scale.

**Figure S2. Q-Q plots of four epigenome-wide analyses of stress/violence measures with additional adjustment for asthma status in model (1) in nasal epithelium from participants in EVA-PR.**

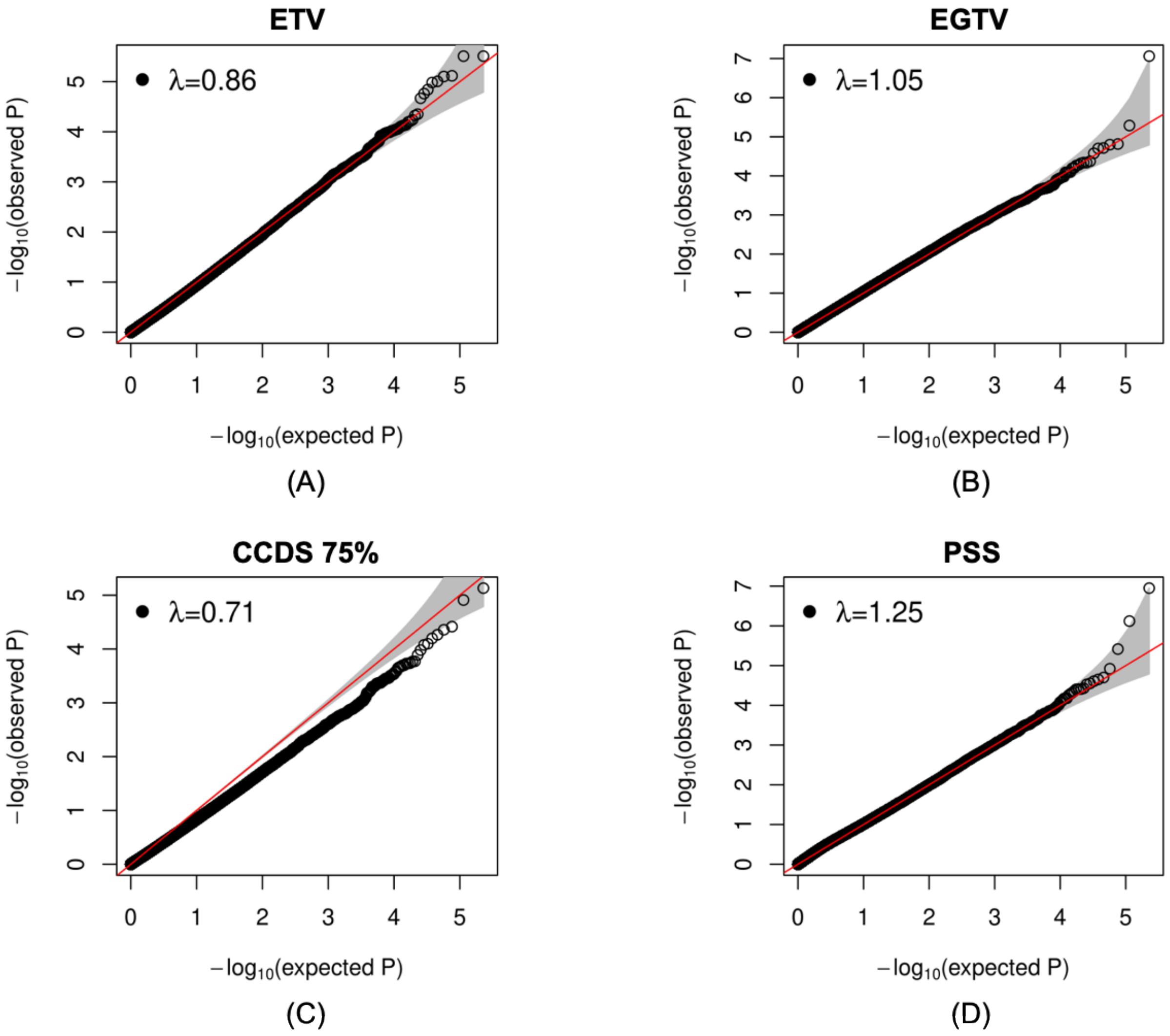


ETV: Exposure to violence. ETGV: Exposure to gun violence. CCDS: Checklist of Children's Distress Symptoms. PSS: Perceived Stress Scale.

**Table S1. Summary of characteristics of participants included in the four ETV/stress EWAS**

| | **ETV** (n=470) | **ETGV** (n=475) | **CCDS** (n=471) | **PSS** (n=476) |
|---|---|---|---|---|
| | 3.8±2.2 | 338 (71.2) | 127 (27.0) | 123 (25.8) |
| Age in years (mean ± SD) | 15.4±2.9 | 15.4±2.9 | 15.4±3.0 | 15.4±2.9 |
| Male gender (n, %) | 243 (51.7) | 245 (51.6) | 244 (51.8) | 246 (51.7) |
| Household income (≥ $15,000 per year; n, %) | 210 (44.7) | 210 (44.2) | 209 (44.4) | 210 (44.1) |
| BMI z-score (mean ± SD) | 0.6±1.1 | 0.6±1.1 | 0.6±1.1 | 0.6±1.1 |
| Distance to roadway (< 444.7 meters; n, %) | 355 (75.5) | 358 (75.4) | 356 (75.6) | 359 (75.4) |

ETV: Exposure to violence. ETGV: Exposure to gun violence. CCDS: Checklist of Children's Distress Symptoms. PSS: Perceived Stress Scale.

**Table S2. Top 100 CpG sites from each of the four ETV/stress EWAS (total 396 CpGs = 400 CpGs – 4 CpGs overlapped in two EWAS) and their corresponding results**

| CpG | Gene | CHR | Position | Beta_ETV | *P*_ETV | Beta_ETGV | *P*_ETGV | Beta_CCDS75 | *P*_CCDS75 | Beta_PSS | *P*_PSS |
|---|---|---|---|---|---|---|---|---|---|---|---|
| cg15009913 | *MORN1* | 1 | 2287741 | -0.0009 | 2.55E-01 | 0.0022 | 5.64E-01 | -0.0121 | 1.11E-03 | 0.0048 | 2.10E-01 |
| cg01150351 | *LINC01346* | 1 | 4220528 | -0.0038 | 3.47E-04 | -0.0074 | 1.45E-01 | -0.0080 | 1.13E-01 | 0.0098 | 5.49E-02 |
| cg16264537 | *LOC284661* | 1 | 4468051 | 0.0016 | 6.92E-02 | 0.0165 | 1.04E-04 | 0.0029 | 4.93E-01 | 0.0028 | 5.24E-01 |
| cg04124888 | *CAPZB* | 1 | 19788288 | -0.0040 | 2.36E-04 | -0.0099 | 5.60E-02 | -0.0039 | 4.48E-01 | -0.0010 | 8.43E-01 |
| cg09683730 | *USP48* | 1 | 22013127 | -0.0036 | 6.30E-05 | -0.0004 | 9.34E-01 | -0.0127 | 2.71E-03 | 0.0010 | 8.24E-01 |
| cg09154062 | *WNT4* | 1 | 22582789 | -0.0008 | 5.47E-01 | -0.0059 | 4.07E-01 | 0.0039 | 5.39E-01 | 0.0214 | 1.54E-05 |
| cg20991983 | *EPHA8* | 1 | 22927528 | -0.0031 | 1.63E-03 | -0.0037 | 4.38E-01 | -0.0181 | 9.25E-05 | 0.0055 | 2.44E-01 |
| cg17104151 | *C1QC* | 1 | 22970132 | -0.0004 | 7.46E-01 | 0.0043 | 4.80E-01 | -0.0044 | 4.53E-01 | 0.0214 | 4.16E-05 |
| cg01974817 | *PTPRU* | 1 | 29802252 | -0.0031 | 3.75E-02 | -0.0023 | 7.53E-01 | -0.0277 | 9.54E-05 | -0.0002 | 9.81E-01 |
| cg17445948 | *LINC01226* | 1 | 31997420 | 0.0009 | 5.73E-01 | 0.0285 | 3.74E-04 | 0.0053 | 5.07E-01 | 0.0056 | 4.86E-01 |
| cg00499831 | *CSMD2* | 1 | 34627314 | -0.0043 | 2.54E-04 | -0.0101 | 6.76E-02 | -0.0095 | 8.51E-02 | -0.0062 | 2.52E-01 |
| cg02865595 | *SH3D21* | 1 | 36771714 | 0.0000 | 9.90E-01 | 0.0046 | 7.61E-01 | 0.0029 | 8.43E-01 | -0.0170 | 1.07E-04 |
| cg20806264 | *MKNK1-AS1* | 1 | 47005728 | -0.0006 | 4.58E-01 | -0.0012 | 7.61E-01 | -0.0130 | 9.53E-04 | -0.0006 | 8.86E-01 |
| cg12708287 | *DIO1* | 1 | 54358446 | -0.0019 | 1.49E-01 | -0.0215 | 5.09E-04 | 0.0012 | 8.41E-01 | -0.0053 | 3.83E-01 |
| cg22361604 | *NFIA* | 1 | 61435148 | -0.0091 | 1.67E-04 | -0.0129 | 2.59E-01 | -0.0118 | 3.00E-01 | 0.0109 | 3.47E-01 |
| cg03050981 | *LEPR* | 1 | 65906670 | -0.0080 | 3.65E-04 | -0.0038 | 7.24E-01 | -0.0070 | 5.12E-01 | 0.0000 | 1.00E+00 |
| cg00116430 | *BCAR3* | 1 | 94188268 | 0.0005 | 8.01E-01 | 0.0047 | 6.35E-01 | 0.0117 | 2.27E-01 | 0.0203 | 1.27E-04 |
| cg16501235 | *C1orf54* | 1 | 150245988 | -0.0015 | 1.41E-01 | -0.0006 | 9.10E-01 | -0.0159 | 9.36E-04 | 0.0035 | 4.79E-01 |
| cg10224088 | *S100A13* | 1 | 153597891 | -0.0010 | 2.04E-01 | NA | NA | -0.0141 | 1.75E-04 | NA | NA |
| cg01134274 | *CLK2* | 1 | 155239493 | -0.0003 | 7.41E-01 | 0.0041 | 3.56E-01 | 0.0145 | 7.79E-04 | 0.0010 | 8.15E-01 |
| cg10518144 | *NUF2* | 1 | 163581520 | 0.0008 | 3.76E-01 | 0.0140 | 5.84E-04 | -0.0008 | 8.39E-01 | 0.0086 | 3.39E-02 |
| cg13550751 | *ABL2* | 1 | 179194565 | 0.0006 | 7.27E-01 | 0.0300 | 5.26E-04 | 0.0019 | 8.26E-01 | -0.0011 | 9.03E-01 |
| cg19827716 | *GM140* | 1 | 181287967 | -0.0014 | 3.19E-01 | -0.0259 | 5.65E-05 | 0.0026 | 6.86E-01 | 0.0082 | 2.07E-01 |
| cg15078825 | *GLUL* | 1 | 182204191 | -0.0025 | 7.76E-03 | -0.0040 | 3.87E-01 | -0.0158 | 4.73E-04 | -0.0033 | 4.67E-01 |
| cg26430597 | *GLUL* | 1 | 182354699 | -0.0011 | 4.73E-01 | -0.0002 | 9.82E-01 | -0.0249 | 5.73E-04 | 0.0002 | 9.70E-01 |
| cg13177053 | *MFSD4* | 1 | 205564088 | -0.0007 | 5.26E-01 | 0.0048 | 3.90E-01 | -0.0191 | 5.08E-04 | 0.0120 | 3.33E-02 |
| cg22032961 | *FCAMR* | 1 | 207152909 | 0.0010 | 4.99E-01 | 0.0113 | 1.13E-01 | -0.0001 | 9.83E-01 | 0.0275 | 3.79E-05 |
| cg16109731 | *DUSP10* | 1 | 221891307 | 0.0020 | 5.17E-01 | 0.0035 | 8.13E-01 | -0.0023 | 8.71E-01 | 0.0337 | 1.66E-04 |
| cg24488001 | *LINC01139* | 1 | 238644629 | -0.0071 | 7.17E-06 | -0.0081 | 2.91E-01 | -0.0201 | 7.61E-03 | -0.0056 | 4.69E-01 |
| cg23616126 | *ZNF496* | 1 | 247463678 | 0.0007 | 3.91E-01 | -0.0013 | 7.38E-01 | -0.0017 | 6.72E-01 | 0.0156 | 9.76E-05 |
| cg26146455 | *PXDN* | 2 | 1681820 | -0.0031 | 1.24E-04 | -0.0068 | 7.75E-02 | -0.0041 | 2.85E-01 | 0.0047 | 2.21E-01 |
| cg11902863 | *TSSC1* | 2 | 3044737 | -0.0015 | 4.48E-01 | -0.0322 | 6.20E-04 | 0.0080 | 3.86E-01 | 0.0092 | 3.27E-01 |
| cg03365705 | *GREB1* | 2 | 11751071 | -0.0033 | 2.22E-04 | -0.0027 | 5.41E-01 | -0.0095 | 2.55E-02 | 0.0030 | 4.94E-01 |
| cg18703983 | *KCNS3* | 2 | 18097337 | 0.0014 | 3.38E-01 | 0.0230 | 6.77E-04 | -0.0025 | 7.12E-01 | 0.0062 | 3.38E-01 |
| cg23221603 | *KCNK3* | 2 | 26963883 | -0.0023 | 1.67E-01 | -0.0115 | 1.51E-01 | -0.0112 | 1.56E-01 | 0.0226 | 1.91E-04 |
| cg18883643 | *ZNF512* | 2 | 27845233 | 0.0012 | 2.60E-01 | 0.0172 | 4.64E-04 | 0.0022 | 6.47E-01 | 0.0054 | 2.81E-01 |
| cg19489704 | *EHD3* | 2 | 31508053 | 0.0008 | 4.67E-01 | 0.0167 | 6.79E-04 | 0.0059 | 2.32E-01 | -0.0032 | 5.19E-01 |
| cg27368726 | *CYP1B1-AS1* | 2 | 38385687 | -0.0021 | 4.83E-02 | -0.0184 | 3.71E-04 | -0.0034 | 5.10E-01 | 0.0173 | 8.99E-04 |
| cg04732596 | *MEIS1* | 2 | 66789740 | 0.0042 | 1.03E-01 | 0.0060 | 6.34E-01 | 0.0397 | 1.12E-03 | 0.0163 | 1.31E-01 |
| cg07720990 | *ADD2* | 2 | 70998383 | -0.0027 | 9.66E-02 | 0.0006 | 9.43E-01 | -0.0269 | 5.24E-04 | -0.0142 | 6.15E-02 |
| cg26907313 | *ATP6V1B1* | 2 | 71167117 | -0.0014 | 1.51E-01 | -0.0029 | 5.36E-01 | -0.0151 | 9.53E-04 | 0.0083 | 7.69E-02 |
| cg02987054 | *INO80B-WBP1* | 2 | 74680627 | 0.0014 | 2.11E-01 | 0.0193 | 2.50E-04 | -0.0015 | 7.78E-01 | -0.0010 | 8.56E-01 |
| cg24434987 | *REG3G* | 2 | 79221130 | 0.0039 | 3.94E-02 | 0.0320 | 4.58E-04 | 0.0081 | 3.65E-01 | -0.0151 | 9.35E-02 |
| cg06827611 | *ITPRIPL1* | 2 | 96993311 | -0.0008 | 3.39E-01 | -0.0060 | 1.20E-01 | 0.0032 | 3.94E-01 | 0.0174 | 9.09E-07 |
| cg25162794 | *ANKRD23* | 2 | 97509831 | 0.0030 | 1.88E-03 | 0.0023 | 6.26E-01 | 0.0166 | 3.08E-04 | 0.0042 | 3.76E-01 |
| cg16573374 | *FAHD2B* | 2 | 97761334 | 0.0022 | 5.19E-02 | 0.0185 | 6.69E-04 | 0.0067 | 2.14E-01 | -0.0016 | 7.72E-01 |
| cg14445389 | *AFF3* | 2 | 100426973 | 0.0031 | 5.22E-01 | 0.0051 | 8.25E-01 | 0.0066 | 7.72E-01 | -0.0363 | 1.25E-04 |
| cg15678392 | *TMEM182* | 2 | 103378135 | -0.0097 | 2.39E-05 | 0.0039 | 7.24E-01 | -0.0081 | 4.55E-01 | 0.0234 | 3.31E-02 |
| cg16099169 | *UXS1* | 2 | 106886729 | -0.0034 | 4.41E-03 | -0.0206 | 2.83E-04 | 0.0006 | 9.09E-01 | 0.0010 | 8.59E-01 |
| cg07461215 | *CNTNAP5* | 2 | 124444215 | -0.0013 | 4.36E-01 | -0.0276 | 4.41E-04 | 0.0053 | 5.00E-01 | 0.0060 | 4.53E-01 |
| cg08231096 | *THSD7B* | 2 | 137748460 | -0.0021 | 1.48E-01 | 0.0003 | 9.62E-01 | -0.0078 | 2.48E-01 | 0.0254 | 1.65E-04 |
| cg00980804 | *RND3* | 2 | 150977441 | 0.0010 | 4.19E-01 | 0.0052 | 3.97E-01 | -0.0024 | 6.92E-01 | 0.0230 | 1.54E-04 |
| cg25417917 | *RND3* | 2 | 151452323 | 0.0011 | 5.03E-01 | 0.0267 | 5.42E-04 | -0.0033 | 6.73E-01 | -0.0105 | 1.61E-01 |
| cg07321354 | *DLX2* | 2 | 173078588 | 0.0030 | 7.11E-02 | 0.0270 | 6.62E-04 | 0.0048 | 5.42E-01 | 0.0030 | 7.10E-01 |
| cg02749511 | *SP3* | 2 | 174696522 | -0.0043 | 1.86E-04 | -0.0017 | 7.55E-01 | -0.0068 | 2.12E-01 | 0.0027 | 5.66E-01 |
| cg18003659 | *CHRNA1* | 2 | 175629267 | 0.0028 | 5.30E-02 | 0.0243 | 4.51E-04 | 0.0102 | 1.39E-01 | -0.0031 | 6.38E-01 |
| cg18448949 | *HOXD8* | 2 | 176993089 | 0.0012 | 1.84E-01 | 0.0149 | 4.74E-04 | -0.0016 | 6.98E-01 | 0.0032 | 4.50E-01 |
| cg02552137 | *ITGA4* | 2 | 182320901 | 0.0001 | 8.78E-01 | 0.0160 | 3.44E-04 | 0.0031 | 4.82E-01 | 0.0028 | 5.42E-01 |
| cg01494441 | *CERKL* | 2 | 182523146 | -0.0036 | 4.22E-02 | 0.0118 | 1.64E-01 | -0.0269 | 1.09E-03 | 0.0104 | 2.15E-01 |
| cg24775027 | *ZNF804A* | 2 | 185459721 | -0.0017 | 1.61E-01 | 0.0001 | 9.88E-01 | -0.0188 | 1.05E-03 | 0.0055 | 3.42E-01 |
| cg09825429 | *ZC3H15* | 2 | 187385867 | -0.0016 | 1.36E-01 | 0.0010 | 8.46E-01 | -0.0039 | 4.45E-01 | -0.0200 | 1.15E-04 |
| cg15599483 | *SLC40A1* | 2 | 190442823 | -0.0016 | 2.06E-01 | -0.0033 | 5.84E-01 | 0.0049 | 4.14E-01 | -0.0221 | 7.80E-05 |
| cg10625666 | *SLC39A10* | 2 | 196521354 | -0.0015 | 5.85E-01 | -0.0065 | 6.29E-01 | -0.0016 | 9.06E-01 | 0.0367 | 5.42E-05 |
| cg18189288 | *SPATS2L* | 2 | 201262081 | -0.0067 | 1.20E-04 | -0.0101 | 2.26E-01 | -0.0148 | 7.53E-02 | -0.0062 | 4.56E-01 |
| cg14905665 | *NRP2* | 2 | 206661494 | -0.0015 | 1.45E-01 | -0.0177 | 4.12E-04 | 0.0043 | 3.87E-01 | 0.0017 | 7.42E-01 |

| | | | | | | | | | | | |
|---|---|---|---|---|---|---|---|---|---|---|---|
| cg24545125 | *TNP1* | 2 | 217724866 | -0.0024 | 7.36E-02 | -0.0074 | 2.54E-01 | -0.0237 | 1.91E-04 | -0.0059 | 3.64E-01 |
| cg23349982 | *DIRC3* | 2 | 218135498 | -0.0024 | 7.62E-02 | -0.0101 | 1.22E-01 | 0.0001 | 9.94E-01 | -0.0264 | 5.42E-05 |
| cg15080232 | *TM4SF20* | 2 | 228228336 | -0.0072 | 2.67E-04 | -0.0093 | 3.25E-01 | 0.0005 | 9.60E-01 | 0.0009 | 9.22E-01 |
| cg13996593 | *DNER* | 2 | 230574596 | -0.0007 | 8.13E-01 | -0.0021 | 8.78E-01 | -0.0150 | 2.59E-01 | 0.0479 | 1.58E-04 |
| cg01011757 | *EIF4E2* | 2 | 233416425 | 0.0001 | 8.97E-01 | 0.0194 | 5.46E-05 | -0.0059 | 2.17E-01 | -0.0030 | 5.30E-01 |
| cg12037784 | *SH3BP4* | 2 | 235962605 | 0.0000 | 9.95E-01 | 0.0017 | 7.21E-01 | 0.0067 | 1.43E-01 | 0.0179 | 1.15E-04 |
| cg06946314 | *SH3BP4* | 2 | 236092957 | -0.0010 | 3.07E-01 | 0.0010 | 8.37E-01 | -0.0072 | 1.26E-01 | 0.0186 | 3.24E-05 |
| cg07611816 | *AGAP1* | 2 | 236760371 | -0.0045 | 3.61E-04 | 0.0010 | 8.69E-01 | 0.0008 | 8.96E-01 | 0.0043 | 4.81E-01 |
| cg18552983 | *ASB18* | 2 | 237128101 | 0.0002 | 7.91E-01 | 0.0071 | 9.43E-02 | -0.0141 | 5.65E-04 | 0.0016 | 7.03E-01 |
| cg10629128 | *HDAC4* | 2 | 240280038 | 0.0014 | 3.69E-01 | -0.0041 | 5.83E-01 | 0.0266 | 2.74E-04 | 0.0000 | 9.99E-01 |
| cg01255646 | *AGXT* | 2 | 241819381 | -0.0018 | 4.49E-02 | -0.0142 | 6.80E-04 | 0.0027 | 5.12E-01 | -0.0012 | 7.85E-01 |
| cg00513781 | *THAP4* | 2 | 242571652 | -0.0039 | 3.81E-04 | 0.0034 | 5.15E-01 | -0.0127 | 1.48E-02 | 0.0027 | 6.17E-01 |
| cg22435335 | *RTP5* | 2 | 242841392 | 0.0020 | 7.13E-02 | 0.0007 | 8.88E-01 | 0.0185 | 3.41E-04 | -0.0065 | 2.13E-01 |
| cg08649360 | *TADA3* | 3 | 9833841 | -0.0013 | 2.42E-01 | 0.0100 | 6.60E-02 | -0.0178 | 6.74E-04 | 0.0056 | 2.92E-01 |
| cg23028654 | *GRIP2* | 3 | 14597729 | -0.0003 | 7.12E-01 | 0.0013 | 7.79E-01 | -0.0012 | 7.85E-01 | 0.0105 | 1.92E-04 |
| cg12382418 | *METTL6* | 3 | 15466952 | -0.0048 | 9.29E-03 | -0.0113 | 2.06E-01 | -0.0296 | 7.60E-04 | -0.0021 | 8.16E-01 |
| cg11863854 | *RBMS3* | 3 | 29471980 | 0.0013 | 7.50E-01 | -0.0017 | 9.32E-01 | -0.0012 | 9.49E-01 | 0.0379 | 7.49E-05 |
| cg08381504 | *PRSS50* | 3 | 46792357 | -0.0035 | 5.02E-02 | -0.0293 | 5.77E-04 | -0.0114 | 1.82E-01 | 0.0041 | 6.39E-01 |
| cg19513744 | *ARHGEF3* | 3 | 56836209 | 0.0002 | 7.38E-01 | 0.0034 | 3.31E-01 | -0.0113 | 9.09E-04 | 0.0018 | 6.08E-01 |
| cg00196474 | *EIF4E3* | 3 | 71702735 | 0.0014 | 1.66E-01 | 0.0200 | 4.09E-05 | -0.0045 | 3.52E-01 | -0.0043 | 3.86E-01 |
| cg26154235 | *PDZRN3* | 3 | 73622059 | 0.0007 | 5.92E-01 | -0.0021 | 7.50E-01 | -0.0043 | 4.94E-01 | 0.0229 | 7.41E-05 |
| cg20982112 | *PVRL3* | 3 | 110911278 | 0.0034 | 3.35E-04 | 0.0158 | 4.07E-04 | 0.0000 | 9.91E-01 | 0.0036 | 4.25E-01 |
| cg26084141 | *PDIA5* | 3 | 122786895 | -0.0004 | 8.17E-01 | 0.0072 | 3.61E-01 | -0.0280 | 1.57E-04 | 0.0048 | 5.24E-01 |
| cg08027748 | *UROC1* | 3 | 126237319 | -0.0033 | 9.07E-06 | -0.0046 | 2.02E-01 | -0.0159 | 6.31E-06 | -0.0017 | 6.32E-01 |
| cg19234171 | *LOC90246* | 3 | 128226071 | 0.0005 | 6.80E-01 | -0.0012 | 8.41E-01 | -0.0007 | 9.02E-01 | 0.0187 | 1.90E-04 |
| cg16848072 | *ANAPC13* | 3 | 134182665 | -0.0012 | 2.35E-01 | -0.0171 | 4.01E-04 | -0.0058 | 2.23E-01 | -0.0016 | 7.48E-01 |
| cg05487105 | *CLDN18* | 3 | 137729296 | -0.0023 | 3.70E-03 | -0.0052 | 1.67E-01 | -0.0031 | 4.06E-01 | 0.0140 | 1.32E-04 |
| cg21924070 | *SPSB4* | 3 | 140788453 | 0.0013 | 8.45E-02 | 0.0127 | 5.94E-04 | 0.0009 | 8.02E-01 | 0.0046 | 2.18E-01 |
| cg15423862 | *KCNAB1* | 3 | 155838109 | 0.0016 | 2.55E-01 | 0.0250 | 1.44E-04 | -0.0026 | 6.86E-01 | -0.0008 | 9.03E-01 |
| cg22481770 | *SSR3* | 3 | 156324118 | 0.0010 | 4.92E-01 | 0.0241 | 4.37E-04 | -0.0012 | 8.63E-01 | -0.0005 | 9.38E-01 |
| cg02235663 | *B3GALNT1* | 3 | 160825607 | -0.0033 | 3.09E-04 | -0.0091 | 3.74E-02 | -0.0115 | 7.72E-03 | 0.0052 | 2.39E-01 |
| cg08827764 | *TNIK* | 3 | 171181416 | -0.0041 | 1.71E-04 | -0.0050 | 3.49E-01 | 0.0049 | 3.54E-01 | -0.0066 | 2.17E-01 |
| cg26832915 | *FAM53A* | 4 | 1577932 | -0.0016 | 1.73E-01 | -0.0051 | 3.73E-01 | -0.0202 | 2.78E-04 | 0.0043 | 4.38E-01 |
| cg05887630 | *SH3TC1* | 4 | 8256315 | 0.0024 | 2.59E-02 | 0.0234 | 4.51E-06 | 0.0054 | 2.82E-01 | -0.0021 | 6.81E-01 |
| cg10169812 | *CD38* | 4 | 15779392 | 0.0023 | 1.65E-02 | 0.0188 | 3.16E-05 | 0.0048 | 2.82E-01 | 0.0083 | 6.96E-02 |
| cg07410217 | *TBC1D1* | 4 | 38153736 | -0.0035 | 2.04E-04 | -0.0098 | 3.44E-02 | -0.0031 | 4.93E-01 | 0.0056 | 2.22E-01 |
| cg02086125 | *LIAS* | 4 | 39463608 | -0.0045 | 8.72E-05 | -0.0037 | 5.07E-01 | -0.0008 | 8.90E-01 | -0.0017 | 7.61E-01 |
| cg09816180 | *GC* | 4 | 72650088 | -0.0040 | 3.04E-04 | 0.0018 | 7.43E-01 | -0.0052 | 3.11E-01 | 0.0000 | 9.93E-01 |
| cg27639662 | *PRDM8* | 4 | 81111393 | 0.0020 | 3.33E-01 | 0.0361 | 3.60E-04 | 0.0090 | 3.56E-01 | 0.0072 | 4.37E-01 |
| cg06440348 | *PRDM8* | 4 | 81111527 | 0.0009 | 4.76E-01 | 0.0250 | 7.65E-05 | -0.0013 | 8.25E-01 | -0.0009 | 8.88E-01 |
| cg02847897 | *KLHL8* | 4 | 88142275 | 0.0044 | 4.64E-02 | 0.0370 | 4.27E-04 | 0.0093 | 3.70E-01 | -0.0085 | 4.10E-01 |
| cg14375050 | *CYP2U1* | 4 | 108848533 | 0.0002 | 9.12E-01 | 0.0281 | 2.83E-04 | 0.0078 | 3.11E-01 | 0.0000 | 9.98E-01 |
| cg11950778 | *UGT8* | 4 | 115519006 | 0.0038 | 4.26E-02 | 0.0305 | 6.10E-04 | -0.0077 | 3.80E-01 | 0.0022 | 8.04E-01 |
| cg17200999 | *SYNPO2* | 4 | 119902588 | 0.0026 | 3.30E-01 | 0.0100 | 4.41E-01 | 0.0118 | 3.56E-01 | 0.0315 | 6.18E-05 |
| cg19463078 | *TNIP3* | 4 | 122099598 | -0.0026 | 2.17E-02 | -0.0188 | 4.77E-04 | 0.0029 | 5.88E-01 | -0.0013 | 8.05E-01 |
| cg26518884 | *CPE* | 4 | 166322679 | 0.0012 | 2.82E-01 | 0.0199 | 3.34E-04 | -0.0028 | 6.12E-01 | -0.0023 | 6.83E-01 |
| cg15948536 | *PALLD* | 4 | 169770092 | -0.0018 | 5.45E-01 | 0.0153 | 2.83E-01 | -0.0451 | 1.22E-03 | -0.0012 | 9.32E-01 |
| cg00612828 | *DCTD* | 4 | 183815971 | -0.0016 | 5.17E-02 | 0.0014 | 7.26E-01 | -0.0161 | 5.17E-05 | 0.0008 | 8.40E-01 |
| cg11869150 | *EXOC3* | 5 | 465665 | -0.0024 | 1.11E-02 | -0.0083 | 6.55E-02 | -0.0148 | 7.79E-04 | -0.0037 | 4.11E-01 |
| cg17851021 | *SLC12A7* | 5 | 1097374 | -0.0002 | 9.06E-01 | 0.0035 | 6.56E-01 | -0.0263 | 5.81E-04 | -0.0022 | 7.74E-01 |
| cg19508967 | *NDUFS6* | 5 | 1840347 | 0.0057 | 2.85E-06 | 0.0096 | 1.05E-01 | 0.0030 | 6.06E-01 | 0.0066 | 2.56E-01 |
| cg09632585 | *ZNF131* | 5 | 43125540 | -0.0052 | 5.78E-06 | -0.0036 | 5.08E-01 | -0.0061 | 2.62E-01 | -0.0095 | 8.22E-02 |
| cg22810546 | *PAIP1* | 5 | 43559688 | -0.0058 | 1.76E-04 | -0.0034 | 6.44E-01 | -0.0142 | 5.29E-02 | -0.0094 | 1.91E-01 |
| cg16506432 | *CDC20B* | 5 | 54428863 | -0.0012 | 2.26E-01 | 0.0036 | 4.57E-01 | 0.0013 | 7.89E-01 | -0.0184 | 1.49E-04 |
| cg10510558 | *MTX3* | 5 | 79290786 | -0.0026 | 1.42E-02 | -0.0085 | 9.14E-02 | -0.0076 | 1.14E-01 | 0.0182 | 1.05E-04 |
| cg17491365 | *TMEM232* | 5 | 109962391 | -0.0030 | 1.26E-02 | -0.0054 | 3.52E-01 | -0.0208 | 2.49E-04 | -0.0045 | 4.46E-01 |
| cg03541903 | *CDC42SE2* | 5 | 130651434 | -0.0055 | 3.38E-03 | -0.0023 | 8.00E-01 | -0.0366 | 3.13E-05 | -0.0157 | 7.96E-02 |
| cg11429111 | *TIFAB* | 5 | 134813329 | -0.0014 | 1.44E-01 | -0.0057 | 2.20E-01 | 0.0043 | 3.38E-01 | 0.0175 | 1.25E-04 |
| cg00513205 | *NEUROG1* | 5 | 134872649 | 0.0011 | 6.49E-01 | -0.0036 | 7.63E-01 | 0.0192 | 1.01E-01 | 0.0279 | 1.25E-04 |
| cg23932798 | *MYOT* | 5 | 137142029 | 0.0007 | 4.21E-01 | 0.0150 | 6.15E-04 | 0.0013 | 7.59E-01 | -0.0052 | 2.46E-01 |
| cg11937703 | *PCDHA1* | 5 | 140307640 | -0.0004 | 8.04E-01 | -0.0024 | 7.42E-01 | -0.0042 | 5.56E-01 | 0.0283 | 4.43E-05 |
| cg03504078 | *PCDHB3* | 5 | 140480218 | 0.0016 | 4.60E-01 | -0.0087 | 4.09E-01 | 0.0340 | 9.11E-04 | 0.0006 | 9.58E-01 |
| cg02563137 | *PCDHGA1* | 5 | 140778546 | -0.0052 | 2.12E-04 | 0.0065 | 3.36E-01 | -0.0118 | 7.83E-02 | -0.0018 | 7.96E-01 |
| cg20963030 | *ATP10B* | 5 | 160280315 | 0.0009 | 5.17E-01 | 0.0006 | 9.24E-01 | 0.0118 | 5.96E-02 | 0.0237 | 1.40E-04 |
| cg08734395 | *MAT2B* | 5 | 163086064 | 0.0002 | 8.98E-01 | 0.0293 | 8.12E-05 | -0.0046 | 5.36E-01 | 0.0009 | 9.06E-01 |
| cg23184477 | *DOCK2* | 5 | 169064897 | 0.0011 | 7.39E-01 | 0.0004 | 9.78E-01 | -0.0230 | 1.33E-01 | 0.0489 | 1.04E-04 |
| cg11927233 | *NPM1* | 5 | 170816542 | -0.0058 | 1.27E-04 | -0.0131 | 6.88E-02 | -0.0011 | 8.78E-01 | -0.0067 | 3.49E-01 |
| cg11892307 | *EXOC2* | 6 | 470595 | 0.0002 | 7.97E-01 | -0.0056 | 2.11E-01 | 0.0142 | 1.08E-03 | 0.0004 | 9.23E-01 |
| cg02409090 | *EXOC2* | 6 | 709725 | -0.0043 | 3.07E-04 | -0.0089 | 1.18E-01 | -0.0066 | 2.39E-01 | 0.0051 | 3.79E-01 |
| cg19521311 | *F13A1* | 6 | 6322347 | 0.0023 | 5.81E-02 | 0.0206 | 3.62E-04 | 0.0089 | 1.20E-01 | -0.0013 | 8.24E-01 |

| | | | | | | | | | | | |
|---|---|---|---|---|---|---|---|---|---|---|---|
| cg23626986 | *TMEM170B* | 6 | 11487376 | -0.0034 | 2.08E-05 | -0.0048 | 2.19E-01 | -0.0015 | 6.93E-01 | -0.0014 | 7.15E-01 |
| cg04611912 | *DTNBP1* | 6 | 15538569 | -0.0007 | 3.60E-01 | -0.0029 | 4.31E-01 | -0.0005 | 9.04E-01 | 0.0141 | 1.84E-04 |
| cg23205775 | *HIST1H2AD* | 6 | 26199576 | 0.0003 | 7.49E-01 | -0.0177 | 2.19E-04 | 0.0008 | 8.69E-01 | 0.0072 | 1.37E-01 |
| cg26529239 | *ZBED9* | 6 | 28550343 | -0.0067 | 1.59E-04 | -0.0063 | 4.62E-01 | -0.0206 | 1.44E-02 | -0.0096 | 2.59E-01 |
| cg17335499 | *OR2B3* | 6 | 29054325 | -0.0005 | 7.09E-01 | 0.0016 | 7.79E-01 | -0.0211 | 1.71E-04 | 0.0038 | 5.07E-01 |
| cg12083232 | *GABBR1* | 6 | 29571438 | 0.0013 | 1.98E-01 | 0.0186 | 1.16E-04 | -0.0011 | 8.12E-01 | -0.0065 | 1.84E-01 |
| cg18298197 | *MRPS18B* | 6 | 30593314 | -0.0031 | 1.79E-04 | -0.0066 | 1.04E-01 | -0.0035 | 3.80E-01 | 0.0005 | 8.97E-01 |
| cg21606953 | *NRM* | 6 | 30657384 | -0.0015 | 1.10E-01 | -0.0025 | 5.79E-01 | -0.0153 | 5.37E-04 | 0.0031 | 5.01E-01 |
| cg02668819 | *TNXB* | 6 | 32057053 | 0.0000 | 9.70E-01 | 0.0155 | 4.86E-04 | -0.0047 | 2.86E-01 | -0.0051 | 2.60E-01 |
| cg21189146 | *TNXB* | 6 | 32069841 | -0.0008 | 3.78E-01 | 0.0038 | 3.78E-01 | -0.0177 | 2.42E-05 | 0.0014 | 7.44E-01 |
| cg20891116 | *BTNL2* | 6 | 32363228 | 0.0003 | 7.68E-01 | 0.0200 | 8.78E-05 | -0.0040 | 4.31E-01 | 0.0014 | 7.89E-01 |
| cg00402668 | *HLA-DOB* | 6 | 32768079 | -0.0022 | 2.65E-02 | 0.0036 | 4.56E-01 | -0.0171 | 2.57E-04 | -0.0010 | 8.31E-01 |
| cg11313335 | *TAP1* | 6 | 32814921 | -0.0019 | 2.78E-02 | -0.0033 | 4.40E-01 | -0.0153 | 2.07E-04 | -0.0085 | 4.42E-02 |
| cg26858238 | *DAXX* | 6 | 33289208 | -0.0003 | 8.58E-01 | 0.0052 | 4.89E-01 | 0.0255 | 4.57E-04 | -0.0044 | 5.59E-01 |
| cg27449651 | *SYNGAP1* | 6 | 33395433 | 0.0046 | 2.86E-04 | 0.0092 | 1.30E-01 | 0.0000 | 9.95E-01 | -0.0077 | 2.12E-01 |
| cg13075951 | *KCNK16* | 6 | 39282393 | 0.0009 | 7.71E-01 | -0.0059 | 6.94E-01 | 0.0108 | 4.66E-01 | 0.0301 | 1.85E-04 |
| cg20377447 | *UNC5CL* | 6 | 41010217 | -0.0036 | 3.80E-05 | -0.0159 | 1.37E-04 | -0.0021 | 6.09E-01 | -0.0007 | 8.73E-01 |
| cg03991152 | *PGC* | 6 | 41715322 | 0.0016 | 6.77E-02 | 0.0145 | 6.34E-04 | -0.0001 | 9.81E-01 | -0.0060 | 1.56E-01 |
| cg21657043 | *MRPL14* | 6 | 44035552 | 0.0002 | 8.13E-01 | 0.0029 | 4.75E-01 | -0.0132 | 7.50E-04 | -0.0020 | 6.25E-01 |
| cg09582880 | *IL17A* | 6 | 52050133 | 0.0014 | 1.06E-01 | 0.0002 | 9.69E-01 | 0.0142 | 4.26E-04 | -0.0005 | 9.04E-01 |
| cg15217784 | *FAM83B* | 6 | 54722227 | 0.0040 | 1.83E-02 | 0.0293 | 3.01E-04 | -0.0114 | 1.48E-01 | 0.0075 | 3.14E-01 |
| cg00988179 | *HMGCLL1* | 6 | 55314478 | -0.0065 | 3.54E-04 | -0.0134 | 1.29E-01 | 0.0023 | 7.82E-01 | -0.0066 | 4.43E-01 |
| cg03922126 | *KCNQ5* | 6 | 73330297 | 0.0061 | 3.89E-04 | -0.0002 | 9.84E-01 | 0.0009 | 9.07E-01 | 0.0152 | 3.74E-02 |
| cg05376362 | *CD109* | 6 | 74826447 | -0.0042 | 3.20E-04 | -0.0114 | 4.02E-02 | -0.0044 | 4.28E-01 | 0.0004 | 9.42E-01 |
| cg08792416 | *SH3BGRL2* | 6 | 80404639 | -0.0005 | 5.22E-01 | 0.0067 | 9.41E-02 | -0.0126 | 1.22E-03 | -0.0020 | 6.15E-01 |
| cg13674558 | *UBE3D* | 6 | 83775899 | 0.0037 | 1.90E-01 | 0.0475 | 5.82E-04 | 0.0152 | 2.60E-01 | 0.0011 | 9.34E-01 |
| cg03076855 | *CEP162* | 6 | 85132322 | 0.0035 | 3.29E-01 | 0.0247 | 1.53E-01 | 0.0252 | 1.37E-01 | 0.0470 | 1.77E-04 |
| cg22884082 | *SMIM8* | 6 | 88039023 | 0.0012 | 3.42E-01 | -0.0039 | 5.23E-01 | 0.0198 | 1.06E-03 | 0.0017 | 7.81E-01 |
| cg00029821 | *STXBP5-AS1* | 6 | 147374493 | 0.0027 | 9.46E-02 | 0.0309 | 8.49E-05 | 0.0074 | 3.34E-01 | 0.0137 | 6.39E-02 |
| cg11071155 | *ARID1B* | 6 | 157012049 | -0.0003 | 7.82E-01 | -0.0005 | 9.09E-01 | 0.0018 | 6.79E-01 | 0.0194 | 1.86E-05 |
| cg16312872 | *TULP4* | 6 | 158923181 | -0.0038 | 1.37E-04 | -0.0133 | 6.33E-03 | -0.0040 | 4.12E-01 | 0.0014 | 7.64E-01 |
| cg01832757 | *FNDC1* | 6 | 159547894 | 0.0019 | 2.82E-01 | 0.0030 | 7.14E-01 | -0.0021 | 7.97E-01 | 0.0234 | 7.73E-05 |
| cg19166759 | *IGF2R* | 6 | 160501174 | -0.0020 | 1.76E-01 | -0.0156 | 2.77E-02 | 0.0093 | 1.87E-01 | 0.0282 | 7.21E-05 |
| cg03034180 | *PACRG* | 6 | 163721894 | -0.0035 | 7.51E-05 | -0.0085 | 4.89E-02 | 0.0008 | 8.52E-01 | 0.0052 | 2.26E-01 |
| cg14078070 | *RPS6KA2* | 6 | 167051518 | -0.0014 | 2.04E-01 | -0.0191 | 2.10E-04 | 0.0101 | 4.64E-02 | 0.0047 | 3.70E-01 |
| cg04471092 | *SMOC2* | 6 | 169246889 | -0.0032 | 1.18E-04 | -0.0030 | 4.49E-01 | -0.0018 | 6.50E-01 | 0.0014 | 7.33E-01 |
| cg21985862 | *THBS2* | 6 | 169559628 | -0.0033 | 1.11E-04 | -0.0098 | 1.85E-02 | -0.0001 | 9.85E-01 | 0.0015 | 7.20E-01 |
| cg20926049 | *DLL1* | 6 | 170591749 | 0.0010 | 2.91E-01 | 0.0107 | 1.38E-02 | 0.0144 | 7.46E-04 | -0.0039 | 3.71E-01 |
| cg25974888 | *FAM20C* | 7 | 84435 | -0.0038 | 3.16E-04 | -0.0121 | 1.78E-02 | 0.0002 | 9.67E-01 | 0.0092 | 6.08E-02 |
| cg18838059 | *PDGFA* | 7 | 534834 | -0.0040 | 3.51E-04 | -0.0129 | 1.54E-02 | -0.0106 | 4.42E-02 | 0.0026 | 6.23E-01 |
| cg05812430 | *INTS1* | 7 | 1542218 | 0.0009 | 2.22E-01 | 0.0129 | 3.52E-04 | 0.0013 | 7.20E-01 | 0.0036 | 3.14E-01 |
| cg02072875 | *IQCE* | 7 | 2606796 | -0.0066 | 2.77E-04 | -0.0109 | 2.15E-01 | -0.0224 | 9.52E-03 | 0.0094 | 2.78E-01 |
| cg03729152 | *TMEM196* | 7 | 19961027 | -0.0014 | 3.34E-01 | -0.0066 | 3.56E-01 | -0.0234 | 7.17E-04 | -0.0093 | 1.92E-01 |
| cg17701991 | *CHN2* | 7 | 29305699 | -0.0035 | 3.80E-04 | -0.0091 | 5.40E-02 | -0.0015 | 7.51E-01 | -0.0031 | 5.16E-01 |
| cg02695349 | *STARD3NL* | 7 | 38269241 | -0.0060 | 5.18E-05 | 0.0010 | 8.91E-01 | -0.0220 | 1.70E-03 | 0.0011 | 8.76E-01 |
| cg18034995 | *VPS41* | 7 | 38805409 | 0.0010 | 4.25E-01 | 0.0212 | 5.43E-04 | -0.0040 | 5.19E-01 | 0.0064 | 3.08E-01 |
| cg06278714 | *HECW1* | 7 | 43287968 | -0.0041 | 3.76E-04 | -0.0090 | 1.13E-01 | -0.0115 | 3.47E-02 | 0.0048 | 3.80E-01 |
| cg14896948 | *COBL* | 7 | 51096885 | -0.0034 | 2.31E-04 | -0.0001 | 9.74E-01 | -0.0044 | 3.07E-01 | 0.0005 | 9.06E-01 |
| cg19616851 | *LINC01005* | 7 | 63363811 | 0.0084 | 1.56E-04 | 0.0249 | 1.94E-02 | 0.0267 | 1.09E-02 | -0.0063 | 4.94E-01 |
| cg11639168 | *POM121* | 7 | 72393693 | 0.0009 | 5.76E-01 | 0.0081 | 2.83E-01 | -0.0067 | 3.62E-01 | 0.0313 | 2.84E-05 |
| cg05857060 | *HIP1* | 7 | 75267630 | -0.0006 | 7.48E-01 | -0.0010 | 9.10E-01 | 0.0085 | 3.36E-01 | 0.0256 | 2.02E-05 |
| cg14392168 | *GAL3ST4* | 7 | 99766665 | 0.0027 | 3.06E-01 | 0.0090 | 4.86E-01 | 0.0473 | 1.54E-04 | -0.0059 | 6.44E-01 |
| cg08986871 | *GIGYF1* | 7 | 100279334 | 0.0001 | 9.40E-01 | -0.0008 | 8.76E-01 | -0.0154 | 1.21E-03 | -0.0012 | 8.08E-01 |
| cg24837397 | *CUX1* | 7 | 101632568 | -0.0009 | 1.87E-01 | -0.0116 | 1.71E-04 | 0.0020 | 5.23E-01 | -0.0053 | 9.14E-02 |
| cg09425926 | *MEST* | 7 | 130124971 | -0.0037 | 8.49E-05 | -0.0082 | 6.98E-02 | -0.0027 | 5.48E-01 | 0.0049 | 2.62E-01 |
| cg15950251 | *TBXAS1* | 7 | 139658345 | 0.0013 | 3.21E-01 | 0.0084 | 1.89E-01 | -0.0135 | 2.97E-02 | 0.0232 | 5.40E-05 |
| cg04388919 | *ZNF777* | 7 | 149119133 | 0.0036 | 2.12E-02 | 0.0252 | 6.16E-04 | -0.0013 | 8.56E-01 | 0.0026 | 7.02E-01 |
| cg12556325 | *LRRC61* | 7 | 150026731 | -0.0025 | 7.36E-02 | -0.0042 | 5.23E-01 | -0.0212 | 1.04E-03 | 0.0125 | 5.91E-02 |
| cg09125924 | *PRKAG2* | 7 | 151452190 | 0.0005 | 7.13E-01 | 0.0084 | 1.76E-01 | 0.0195 | 1.21E-03 | -0.0037 | 5.47E-01 |
| cg19064846 | *PTPRN2* | 7 | 157273391 | -0.0003 | 8.08E-01 | 0.0033 | 6.01E-01 | -0.0091 | 1.40E-01 | 0.0283 | 3.36E-07 |
| cg17255108 | *PTPRN2* | 7 | 157338433 | -0.0012 | 2.68E-01 | -0.0008 | 8.79E-01 | -0.0191 | 1.61E-04 | -0.0075 | 1.46E-01 |
| cg02323550 | *PTPRN2* | 7 | 157660321 | -0.0038 | 3.63E-06 | 0.0001 | 9.82E-01 | -0.0087 | 2.69E-02 | -0.0011 | 7.77E-01 |
| cg08530934 | *PTPRN2* | 7 | 158236160 | -0.0037 | 3.41E-04 | -0.0045 | 3.63E-01 | -0.0069 | 1.56E-01 | -0.0022 | 6.47E-01 |
| cg13632655 | *PTPRN2* | 7 | 158266097 | -0.0017 | 2.94E-01 | 0.0009 | 9.03E-01 | -0.0004 | 9.52E-01 | 0.0176 | 1.10E-04 |
| cg12283658 | *WDR60* | 7 | 158702581 | -0.0009 | 4.60E-01 | 0.0246 | 3.60E-05 | 0.0050 | 3.98E-01 | -0.0064 | 2.94E-01 |
| cg10300293 | *ERICH1* | 8 | 643753 | 0.0001 | 9.13E-01 | 0.0012 | 7.79E-01 | 0.0024 | 5.79E-01 | 0.0142 | 1.34E-04 |
| cg07399532 | *MSRA* | 8 | 10001612 | -0.0018 | 1.07E-01 | -0.0069 | 1.84E-01 | 0.0026 | 6.07E-01 | 0.0184 | 1.24E-04 |
| cg23883874 | *PRSS55* | 8 | 10405415 | -0.0009 | 5.65E-01 | 0.0079 | 3.06E-01 | -0.0108 | 1.49E-01 | 0.0233 | 1.62E-04 |
| cg21799053 | *RHOBTB2* | 8 | 22856801 | -0.0012 | 1.48E-01 | -0.0140 | 3.66E-04 | 0.0015 | 6.90E-01 | -0.0061 | 1.23E-01 |
| cg02501086 | *ADRA1A* | 8 | 26717875 | -0.0006 | 5.45E-01 | -0.0068 | 1.29E-01 | 0.0021 | 6.43E-01 | 0.0173 | 1.41E-04 |

| | | | | | | | | | | | |
|---|---|---|---|---|---|---|---|---|---|---|---|
| cg05276226 | *ADAM18* | 8 | 39441968 | -0.0071 | 1.98E-04 | -0.0099 | 2.85E-01 | -0.0278 | 2.19E-03 | 0.0111 | 2.29E-01 |
| cg05072559 | *SNAI2* | 8 | 49813855 | -0.0077 | 3.34E-04 | -0.0092 | 3.74E-01 | -0.0023 | 8.24E-01 | -0.0051 | 6.25E-01 |
| cg21449463 | *MRPL15* | 8 | 55091759 | -0.0086 | 2.79E-03 | -0.0085 | 5.39E-01 | -0.0444 | 1.05E-03 | 0.0077 | 5.55E-01 |
| cg27506254 | *XKR4* | 8 | 56436472 | -0.0009 | 3.95E-01 | 0.0025 | 6.40E-01 | -0.0186 | 3.55E-04 | 0.0059 | 2.65E-01 |
| cg08035479 | *LOC286177* | 8 | 58172643 | 0.0019 | 2.55E-01 | -0.0014 | 8.57E-01 | 0.0250 | 1.12E-03 | 0.0033 | 6.69E-01 |
| cg14012365 | *FAM110B* | 8 | 59059187 | -0.0016 | 5.55E-02 | -0.0061 | 1.38E-01 | -0.0133 | 8.87E-04 | 0.0037 | 3.68E-01 |
| cg13580380 | *MSC* | 8 | 72667937 | 0.0000 | 9.78E-01 | 0.0017 | 7.24E-01 | -0.0048 | 3.19E-01 | 0.0159 | 8.92E-05 |
| cg27642416 | *REXO1L2P* | 8 | 86554026 | -0.0058 | 1.08E-04 | -0.0168 | 1.84E-02 | -0.0021 | 7.62E-01 | 0.0008 | 9.09E-01 |
| cg05314679 | *PABPC1* | 8 | 101821998 | 0.0005 | 6.58E-01 | -0.0032 | 5.92E-01 | -0.0045 | 4.11E-01 | 0.0166 | 1.90E-04 |
| cg04440283 | *TRHR* | 8 | 110098923 | -0.0058 | 3.29E-03 | -0.0329 | 4.12E-04 | -0.0071 | 4.46E-01 | 0.0172 | 6.69E-02 |
| cg07147958 | *LINC00536* | 8 | 116822442 | 0.0021 | 2.22E-01 | 0.0347 | 2.45E-05 | -0.0028 | 7.37E-01 | -0.0028 | 7.40E-01 |
| cg01542423 | *TRIB1* | 8 | 126668119 | -0.0001 | 9.28E-01 | -0.0012 | 8.36E-01 | -0.0008 | 8.90E-01 | 0.0237 | 7.28E-06 |
| cg24314877 | *MYC* | 8 | 128656055 | 0.0006 | 6.05E-01 | -0.0010 | 8.65E-01 | 0.0022 | 7.03E-01 | 0.0189 | 1.58E-04 |
| cg02546828 | *FAM49B* | 8 | 130838883 | 0.0041 | 6.62E-03 | 0.0271 | 1.80E-04 | 0.0084 | 2.41E-01 | -0.0012 | 8.64E-01 |
| cg25799241 | *FAM83H* | 8 | 144805740 | -0.0032 | 1.60E-02 | -0.0001 | 9.88E-01 | -0.0212 | 5.38E-04 | -0.0013 | 8.32E-01 |
| cg05623411 | *ZDHHC21* | 9 | 14691289 | -0.0105 | 1.13E-04 | -0.0166 | 2.12E-01 | -0.0376 | 3.41E-03 | 0.0014 | 9.10E-01 |
| cg13804467 | *LINGO2* | 9 | 28907222 | -0.0005 | 7.77E-01 | 0.0184 | 2.31E-02 | -0.0266 | 9.09E-04 | 0.0109 | 1.64E-01 |
| cg21199398 | *TRPM6* | 9 | 77440667 | 0.0007 | 4.32E-01 | 0.0059 | 1.40E-01 | -0.0141 | 3.45E-04 | 0.0011 | 7.81E-01 |
| cg00306063 | *UNQ6494* | 9 | 92289643 | -0.0028 | 2.65E-04 | -0.0066 | 7.29E-02 | -0.0027 | 4.49E-01 | -0.0022 | 5.43E-01 |
| cg14220055 | *LOC158434* | 9 | 98885705 | 0.0017 | 1.85E-01 | 0.0211 | 5.91E-04 | -0.0031 | 6.06E-01 | 0.0014 | 8.24E-01 |
| cg06119575 | *TAL2* | 9 | 108424905 | 0.0008 | 4.64E-01 | 0.0189 | 3.56E-04 | -0.0047 | 3.72E-01 | -0.0007 | 8.97E-01 |
| cg14260530 | *AKNA* | 9 | 117157871 | -0.0009 | 4.78E-01 | 0.0003 | 9.65E-01 | -0.0052 | 3.83E-01 | 0.0204 | 5.80E-05 |
| cg13483797 | *PRRX2* | 9 | 132431581 | -0.0032 | 2.93E-02 | -0.0034 | 6.33E-01 | -0.0246 | 3.76E-04 | 0.0041 | 5.57E-01 |
| cg00648203 | *NACC2* | 9 | 138967195 | -0.0005 | 5.99E-01 | -0.0015 | 7.32E-01 | 0.0021 | 6.37E-01 | 0.0190 | 1.75E-05 |
| cg26725665 | *DIP2C* | 10 | 636140 | -0.0032 | 2.27E-04 | -0.0159 | 1.24E-04 | 0.0035 | 3.97E-01 | 0.0059 | 1.60E-01 |
| cg24910943 | *CELF2* | 10 | 11206320 | 0.0005 | 5.63E-01 | -0.0015 | 7.41E-01 | -0.0018 | 6.75E-01 | 0.0158 | 8.26E-05 |
| cg22601058 | *LOC283070* | 10 | 12874207 | -0.0021 | 4.00E-01 | -0.0013 | 9.13E-01 | -0.0167 | 1.66E-01 | 0.0237 | 8.68E-05 |
| cg17482424 | *BAMBI* | 10 | 28970998 | 0.0003 | 7.98E-01 | -0.0062 | 3.17E-01 | 0.0203 | 8.03E-04 | 0.0053 | 3.84E-01 |
| cg12810264 | *BMS1* | 10 | 43329092 | 0.0003 | 7.32E-01 | 0.0019 | 6.81E-01 | -0.0154 | 5.61E-04 | 0.0008 | 8.64E-01 |
| cg12433084 | *GDF10* | 10 | 48430289 | 0.0011 | 6.69E-01 | 0.0221 | 7.52E-02 | -0.0097 | 4.30E-01 | 0.0234 | 1.45E-05 |
| cg13558105 | *C10orf71* | 10 | 50530868 | -0.0040 | 9.90E-05 | -0.0024 | 6.32E-01 | -0.0038 | 4.30E-01 | 0.0018 | 7.01E-01 |
| cg11709763 | *COL13A1* | 10 | 71570077 | 0.0040 | 4.48E-02 | 0.0335 | 4.10E-04 | -0.0062 | 5.06E-01 | 0.0071 | 3.91E-01 |
| cg13585930 | *NPFFR1* | 10 | 72027357 | -0.0007 | 6.31E-01 | 0.0045 | 5.20E-01 | -0.0049 | 4.71E-01 | 0.0267 | 9.37E-05 |
| cg18195276 | *P4HA1* | 10 | 74857663 | 0.0039 | 2.98E-04 | 0.0109 | 3.67E-02 | 0.0070 | 1.76E-01 | 0.0081 | 1.26E-01 |
| cg18783033 | *LIPK* | 10 | 90488297 | 0.0017 | 4.62E-01 | 0.0108 | 3.28E-01 | -0.0007 | 9.45E-01 | 0.0246 | 3.77E-05 |
| cg09154166 | *SLC35G1* | 10 | 95608979 | -0.0037 | 8.53E-05 | -0.0051 | 2.62E-01 | -0.0039 | 3.93E-01 | -0.0001 | 9.79E-01 |
| cg06569542 | *ARHGAP19-SLIT1* | 10 | 98946673 | -0.0002 | 8.04E-01 | -0.0067 | 1.32E-01 | 0.0029 | 5.03E-01 | 0.0152 | 1.99E-04 |
| cg04368939 | *KCNIP2* | 10 | 103588914 | 0.0004 | 6.95E-01 | 0.0060 | 2.45E-01 | 0.0043 | 4.01E-01 | 0.0144 | 6.55E-05 |
| cg18188150 | *PPRC1* | 10 | 103895431 | -0.0003 | 8.47E-01 | 0.0049 | 4.69E-01 | -0.0223 | 5.29E-04 | 0.0110 | 8.11E-02 |
| cg04688330 | *SH3PXD2A* | 10 | 105451802 | -0.0003 | 7.26E-01 | 0.0003 | 9.26E-01 | -0.0019 | 5.77E-01 | 0.0135 | 5.25E-05 |
| cg01266390 | *BUB3* | 10 | 125034002 | 0.0003 | 8.76E-01 | -0.0043 | 6.08E-01 | -0.0094 | 2.51E-01 | 0.0304 | 5.89E-05 |
| cg19686940 | *C10orf90* | 10 | 128193332 | 0.0001 | 8.78E-01 | -0.0048 | 2.47E-01 | 0.0060 | 1.41E-01 | 0.0162 | 6.14E-05 |
| cg03344290 | *MGMT* | 10 | 131448630 | 0.0017 | 1.88E-02 | 0.0010 | 7.75E-01 | 0.0139 | 6.33E-05 | 0.0009 | 8.05E-01 |
| cg02094360 | *TCERG1L* | 10 | 133326929 | -0.0028 | 7.13E-03 | -0.0173 | 6.74E-04 | -0.0039 | 4.36E-01 | 0.0018 | 7.24E-01 |
| cg18961589 | *LINC01164* | 10 | 133598669 | 0.0019 | 1.03E-01 | 0.0294 | 1.28E-07 | -0.0017 | 7.63E-01 | -0.0072 | 2.03E-01 |
| cg15486233 | *SPRN* | 10 | 135237205 | -0.0014 | 1.09E-01 | 0.0093 | 2.64E-02 | -0.0135 | 1.09E-03 | 0.0019 | 6.49E-01 |
| cg20522006 | *MUC2* | 11 | 1097259 | 0.0013 | 1.87E-01 | 0.0016 | 7.39E-01 | -0.0008 | 8.63E-01 | 0.0184 | 8.81E-05 |
| cg19547124 | *MOB2* | 11 | 1669376 | -0.0018 | 1.80E-01 | -0.0065 | 3.10E-01 | -0.0034 | 5.89E-01 | 0.0243 | 4.37E-05 |
| cg19290938 | *MRPL23* | 11 | 1988758 | -0.0012 | 4.17E-01 | -0.0042 | 5.39E-01 | -0.0042 | 5.26E-01 | 0.0226 | 3.83E-05 |
| cg25204743 | *KCNQ1* | 11 | 2715837 | -0.0001 | 9.68E-01 | -0.0034 | 6.42E-01 | 0.0055 | 4.51E-01 | 0.0219 | 1.07E-04 |
| cg22366698 | *ZNF195* | 11 | 3400543 | -0.0023 | 3.69E-02 | -0.0026 | 6.25E-01 | -0.0181 | 4.15E-04 | 0.0037 | 4.84E-01 |
| cg17025642 | *TRIM5* | 11 | 5709468 | -0.0074 | 2.39E-04 | -0.0155 | 1.10E-01 | -0.0101 | 2.87E-01 | -0.0169 | 7.90E-02 |
| cg06595154 | *MRVI1* | 11 | 10716164 | -0.0051 | 3.69E-02 | -0.0094 | 4.31E-01 | -0.0437 | 1.37E-04 | 0.0122 | 2.71E-01 |
| cg27062927 | *USH1C* | 11 | 17555142 | -0.0007 | 4.13E-01 | 0.0056 | 1.57E-01 | -0.0133 | 6.21E-04 | -0.0027 | 4.99E-01 |
| cg11302992 | *ELP4* | 11 | 31789446 | 0.0010 | 4.56E-01 | 0.0239 | 2.88E-04 | 0.0022 | 7.34E-01 | -0.0024 | 7.22E-01 |
| cg02801405 | *LOC100507384* | 11 | 45727997 | -0.0042 | 1.30E-04 | -0.0107 | 4.32E-02 | -0.0041 | 4.37E-01 | -0.0015 | 7.77E-01 |
| cg05149097 | *OSBP* | 11 | 59325534 | -0.0006 | 7.56E-01 | 0.0094 | 2.86E-01 | -0.0305 | 4.15E-04 | 0.0061 | 4.79E-01 |
| cg09801334 | *LTBP3* | 11 | 65315625 | -0.0039 | 2.48E-04 | -0.0127 | 1.30E-02 | 0.0032 | 5.16E-01 | 0.0009 | 8.56E-01 |
| cg23839479 | *MYEOV* | 11 | 69181986 | 0.0022 | 4.93E-02 | 0.0185 | 5.42E-04 | 0.0087 | 9.98E-02 | 0.0054 | 3.22E-01 |
| cg02104112 | *CCND1* | 11 | 69418071 | 0.0029 | 9.73E-02 | 0.0097 | 2.48E-01 | 0.0031 | 7.05E-01 | 0.0232 | 1.78E-04 |
| cg08458678 | *SHANK2* | 11 | 70565177 | -0.0006 | 5.49E-01 | -0.0169 | 1.56E-04 | 0.0049 | 2.62E-01 | 0.0024 | 5.87E-01 |
| cg19942977 | *DLG2* | 11 | 83394759 | 0.0000 | 9.69E-01 | 0.0056 | 3.27E-01 | -0.0184 | 1.00E-03 | -0.0039 | 4.93E-01 |
| cg03537802 | *CEP126* | 11 | 101868388 | 0.0016 | 3.16E-01 | 0.0264 | 6.19E-04 | -0.0013 | 8.61E-01 | 0.0012 | 8.79E-01 |
| cg17076485 | *BUD13* | 11 | 116580712 | 0.0008 | 5.44E-01 | 0.0064 | 3.09E-01 | -0.0063 | 3.09E-01 | 0.0224 | 6.09E-05 |
| cg26502105 | *FXYD6-FXYD2* | 11 | 117701909 | -0.0041 | 6.01E-05 | -0.0090 | 6.48E-02 | -0.0015 | 7.61E-01 | 0.0033 | 4.84E-01 |
| cg27314569 | *BSX* | 11 | 122852461 | 0.0014 | 9.36E-02 | 0.0146 | 2.23E-04 | -0.0027 | 4.91E-01 | 0.0018 | 6.51E-01 |
| cg03455964 | *PATE1* | 11 | 125615317 | 0.0001 | 9.32E-01 | 0.0061 | 1.57E-01 | -0.0150 | 3.32E-04 | 0.0015 | 7.21E-01 |
| cg16358741 | *ST14* | 11 | 130076257 | -0.0044 | 3.59E-04 | -0.0149 | 1.17E-02 | -0.0049 | 4.01E-01 | 0.0086 | 1.35E-01 |
| cg20872252 | *LOC283177* | 11 | 134287326 | -0.0051 | 3.25E-04 | -0.0020 | 7.69E-01 | -0.0031 | 6.45E-01 | 0.0106 | 1.05E-01 |
| cg08726801 | *CACNA2D4* | 12 | 1974122 | -0.0010 | 4.94E-01 | 0.0079 | 2.76E-01 | -0.0004 | 9.56E-01 | -0.0272 | 1.79E-04 |

| | | | | | | | | | | | |
|---|---|---|---|---|---|---|---|---|---|---|---|
| cg01039401 | *TEAD4* | 12 | 3070406 | 0.0000 | 9.99E-01 | 0.0035 | 7.72E-01 | -0.0386 | 1.03E-03 | 0.0053 | 5.65E-01 |
| cg02140508 | *RAD51AP1* | 12 | 4659038 | -0.0054 | 3.21E-04 | 0.0017 | 8.13E-01 | -0.0086 | 2.20E-01 | 0.0064 | 3.71E-01 |
| cg01181415 | *LMO3* | 12 | 16757954 | 0.0022 | 5.09E-01 | 0.0069 | 6.58E-01 | 0.0043 | 7.83E-01 | 0.0362 | 3.76E-05 |
| cg16205352 | *PRPH* | 12 | 49688176 | 0.0005 | 7.05E-01 | 0.0049 | 4.42E-01 | 0.0022 | 7.26E-01 | 0.0219 | 1.46E-04 |
| cg19482754 | *TRPV4* | 12 | 110224547 | -0.0024 | 1.66E-02 | -0.0200 | 2.72E-05 | 0.0015 | 7.45E-01 | -0.0011 | 8.12E-01 |
| cg05134054 | *CUX2* | 12 | 111751588 | -0.0010 | 3.91E-01 | 0.0019 | 7.38E-01 | -0.0024 | 6.68E-01 | 0.0219 | 9.16E-05 |
| cg09265016 | *TBX3* | 12 | 115170515 | -0.0015 | 3.98E-01 | -0.0010 | 9.06E-01 | -0.0020 | 8.08E-01 | 0.0236 | 1.23E-04 |
| cg08932683 | *RNFT2* | 12 | 117253648 | 0.0008 | 5.10E-01 | 0.0201 | 3.57E-04 | 0.0027 | 6.31E-01 | 0.0030 | 6.02E-01 |
| cg09296697 | *MSI1* | 12 | 120833809 | 0.0005 | 6.43E-01 | 0.0182 | 6.43E-04 | 0.0019 | 7.10E-01 | 0.0031 | 5.48E-01 |
| cg24309215 | *POP5* | 12 | 121021560 | 0.0023 | 1.20E-01 | 0.0240 | 6.77E-04 | 0.0034 | 6.24E-01 | 0.0137 | 5.40E-02 |
| cg01281501 | *ADGRD1* | 12 | 131494451 | -0.0036 | 1.41E-04 | -0.0062 | 1.81E-01 | -0.0059 | 1.95E-01 | 0.0000 | 9.99E-01 |
| cg15159017 | *GALNT9* | 12 | 132854526 | 0.0042 | 3.64E-04 | 0.0067 | 2.51E-01 | -0.0009 | 8.71E-01 | 0.0040 | 4.38E-01 |
| cg19838358 | *ANKLE2* | 12 | 133304061 | -0.0017 | 3.36E-02 | -0.0142 | 2.69E-04 | 0.0063 | 1.06E-01 | 0.0029 | 4.53E-01 |
| cg10236247 | *IL17D* | 13 | 21276233 | -0.0002 | 8.81E-01 | 0.0218 | 8.93E-05 | 0.0003 | 9.59E-01 | 0.0115 | 3.98E-02 |
| cg11947245 | *SLC46A3* | 13 | 29328979 | -0.0018 | 7.31E-02 | -0.0182 | 1.36E-04 | -0.0084 | 7.23E-02 | -0.0022 | 6.46E-01 |
| cg12017061 | *ALOX5AP* | 13 | 31271512 | -0.0068 | 3.10E-04 | -0.0180 | 4.57E-02 | -0.0088 | 3.26E-01 | 0.0078 | 2.97E-01 |
| cg16525987 | *TEX26-AS1* | 13 | 31506376 | -0.0061 | 1.50E-04 | -0.0092 | 2.42E-01 | -0.0125 | 1.06E-01 | -0.0063 | 4.22E-01 |
| cg25376651 | *EEF1DP3* | 13 | 32505135 | -0.0013 | 6.27E-01 | -0.0462 | 5.00E-04 | -0.0088 | 5.00E-01 | 0.0200 | 1.24E-01 |
| cg00661205 | *VWA8* | 13 | 42400902 | -0.0042 | 2.12E-04 | 0.0037 | 4.94E-01 | -0.0126 | 1.90E-02 | 0.0010 | 8.58E-01 |
| cg23869158 | *LRCH1* | 13 | 47126023 | 0.0011 | 4.47E-01 | 0.0251 | 4.80E-04 | -0.0022 | 7.62E-01 | -0.0014 | 8.50E-01 |
| cg00750481 | *SPRY2* | 13 | 80755568 | -0.0177 | 1.66E-05 | -0.0319 | 1.08E-01 | -0.0114 | 5.61E-01 | -0.0240 | 2.30E-01 |
| cg27300028 | *CLYBL* | 13 | 100312810 | -0.0015 | 1.71E-01 | 0.0006 | 9.05E-01 | -0.0183 | 2.73E-04 | 0.0049 | 3.46E-01 |
| cg25876057 | *CCDC168* | 13 | 103387002 | 0.0006 | 7.00E-01 | -0.0018 | 8.25E-01 | 0.0258 | 1.04E-03 | -0.0080 | 3.17E-01 |
| cg11893488 | *COL4A2* | 13 | 111144464 | -0.0027 | 3.87E-04 | -0.0019 | 6.05E-01 | -0.0093 | 1.02E-02 | 0.0048 | 1.93E-01 |
| cg25247122 | *SOX1* | 13 | 112652556 | -0.0043 | 9.60E-05 | -0.0071 | 1.87E-01 | 0.0029 | 5.82E-01 | 0.0046 | 3.89E-01 |
| cg21186708 | *SOX1* | 13 | 112716728 | -0.0003 | 6.95E-01 | 0.0071 | 8.14E-02 | -0.0151 | 1.28E-04 | 0.0036 | 3.53E-01 |
| cg10327980 | *FITM1* | 14 | 24601001 | -0.0043 | 2.64E-04 | -0.0096 | 8.93E-02 | -0.0011 | 8.35E-01 | 0.0071 | 1.82E-01 |
| cg16577132 | *STRN3* | 14 | 31408344 | -0.0041 | 3.78E-04 | -0.0081 | 1.39E-01 | 0.0008 | 8.87E-01 | 0.0036 | 5.08E-01 |
| cg04990977 | *SLC35F4* | 14 | 58221774 | -0.0008 | 6.37E-01 | 0.0076 | 3.85E-01 | -0.0008 | 9.28E-01 | 0.0270 | 6.12E-05 |
| cg14685575 | *RTN1* | 14 | 60193832 | -0.0034 | 1.69E-04 | -0.0064 | 1.41E-01 | -0.0109 | 9.71E-03 | 0.0019 | 6.48E-01 |
| cg04921109 | *PLEKHD1* | 14 | 69952104 | 0.0030 | 4.70E-02 | 0.0088 | 2.34E-01 | 0.0054 | 4.57E-01 | 0.0280 | 8.38E-05 |
| cg04127998 | *IFT43* | 14 | 76518992 | -0.0003 | 7.53E-01 | 0.0154 | 4.84E-04 | -0.0085 | 4.87E-02 | 0.0033 | 4.50E-01 |
| cg18666249 | *FOXN3* | 14 | 89670502 | -0.0016 | 3.49E-01 | 0.0157 | 6.06E-02 | -0.0310 | 1.51E-04 | 0.0105 | 2.12E-01 |
| cg25090315 | *GPR68* | 14 | 91705005 | -0.0019 | 3.01E-02 | -0.0078 | 7.26E-02 | -0.0137 | 9.53E-04 | -0.0067 | 1.17E-01 |
| cg16244889 | *PRIMA1* | 14 | 94211818 | -0.0012 | 4.05E-01 | -0.0042 | 5.59E-01 | -0.0065 | 3.54E-01 | 0.0304 | 7.35E-06 |
| cg19774899 | *DLK1* | 14 | 101203896 | 0.0014 | 1.28E-01 | 0.0174 | 7.63E-05 | 0.0002 | 9.58E-01 | 0.0019 | 6.61E-01 |
| cg13150061 | *WDR20* | 14 | 102685953 | -0.0014 | 2.04E-01 | -0.0081 | 1.17E-01 | -0.0004 | 9.32E-01 | -0.0210 | 4.01E-05 |
| cg16097532 | *AHNAK2* | 14 | 105416166 | -0.0015 | 4.27E-01 | -0.0011 | 9.04E-01 | -0.0285 | 1.19E-03 | -0.0132 | 1.42E-01 |
| cg08343671 | *SNORD115-7* | 15 | 25427509 | -0.0051 | 1.64E-04 | -0.0123 | 5.78E-02 | -0.0072 | 2.59E-01 | 0.0089 | 1.68E-01 |
| cg24687432 | *ATP10A* | 15 | 26109608 | -0.0052 | 8.88E-05 | -0.0077 | 2.36E-01 | -0.0115 | 6.91E-02 | 0.0073 | 2.42E-01 |
| cg13619277 | *FMN1* | 15 | 33307322 | -0.0003 | 9.43E-01 | 0.0174 | 3.69E-01 | -0.0188 | 3.26E-01 | 0.0505 | 1.92E-04 |
| cg00397575 | *RASL12* | 15 | 65359932 | 0.0008 | 1.84E-01 | 0.0010 | 7.26E-01 | -0.0010 | 7.43E-01 | 0.0108 | 1.62E-04 |
| cg18011253 | *ISLR2* | 15 | 74424054 | -0.0026 | 6.62E-05 | -0.0040 | 2.15E-01 | -0.0075 | 1.55E-02 | 0.0016 | 6.10E-01 |
| cg05537316 | *SEMA7A* | 15 | 74706314 | 0.0012 | 1.68E-01 | 0.0150 | 4.56E-04 | -0.0030 | 4.78E-01 | 0.0023 | 5.82E-01 |
| cg21376795 | *KLHL25* | 15 | 86302121 | -0.0007 | 4.18E-01 | 0.0146 | 5.68E-04 | -0.0011 | 7.98E-01 | 0.0032 | 4.59E-01 |
| cg14378769 | *MCTP2* | 15 | 94443296 | -0.0034 | 3.85E-04 | -0.0025 | 5.89E-01 | -0.0034 | 4.67E-01 | 0.0002 | 9.70E-01 |
| cg18146152 | *TSR3* | 16 | 1400772 | 0.0003 | 7.77E-01 | -0.0069 | 2.01E-01 | 0.0181 | 4.88E-04 | -0.0062 | 2.38E-01 |
| cg01522083 | *TSR3* | 16 | 1400861 | -0.0007 | 5.72E-01 | -0.0055 | 3.60E-01 | -0.0051 | 3.88E-01 | 0.0191 | 1.74E-04 |
| cg08032483 | *CLCN7* | 16 | 1495363 | 0.0009 | 4.19E-01 | 0.0012 | 8.18E-01 | 0.0169 | 7.94E-04 | 0.0067 | 1.91E-01 |
| cg26552163 | *IFT140* | 16 | 1589859 | -0.0017 | 8.73E-02 | -0.0183 | 7.28E-05 | -0.0026 | 5.65E-01 | -0.0090 | 4.86E-02 |
| cg02898051 | *MTRNR2L4* | 16 | 3415256 | 0.0018 | 7.25E-02 | 0.0046 | 3.44E-01 | 0.0157 | 1.04E-03 | -0.0077 | 1.15E-01 |
| cg03402351 | *SNN* | 16 | 11759492 | -0.0032 | 5.44E-02 | -0.0069 | 3.89E-01 | -0.0050 | 5.18E-01 | 0.0396 | 1.69E-07 |
| cg03102879 | *SCNN1B* | 16 | 23313295 | 0.0016 | 1.15E-01 | 0.0022 | 6.48E-01 | 0.0159 | 7.23E-04 | 0.0052 | 2.79E-01 |
| cg27078729 | *TNRC6A* | 16 | 24835194 | 0.0003 | 7.48E-01 | 0.0166 | 3.39E-04 | -0.0008 | 8.56E-01 | 0.0046 | 3.22E-01 |
| cg08033935 | *C16orf58* | 16 | 31505179 | -0.0002 | 8.48E-01 | 0.0064 | 2.28E-01 | -0.0042 | 4.15E-01 | 0.0174 | 9.75E-05 |
| cg27505745 | *UBE2MP1* | 16 | 34411816 | 0.0047 | 3.63E-04 | 0.0192 | 2.31E-03 | 0.0032 | 6.08E-01 | -0.0016 | 8.01E-01 |
| cg16746369 | *SALL1* | 16 | 51192174 | -0.0017 | 9.30E-02 | -0.0027 | 5.73E-01 | -0.0155 | 8.85E-04 | 0.0005 | 9.13E-01 |
| cg07006398 | *IRX6* | 16 | 55406516 | 0.0038 | 1.21E-01 | 0.0406 | 5.15E-04 | -0.0158 | 1.75E-01 | -0.0040 | 6.96E-01 |
| cg16660891 | *USP10* | 16 | 84828913 | -0.0014 | 2.29E-01 | -0.0021 | 6.97E-01 | -0.0172 | 1.14E-03 | -0.0037 | 4.47E-01 |
| cg16544989 | *GSE1* | 16 | 85600639 | -0.0049 | 3.26E-04 | -0.0083 | 2.11E-01 | -0.0093 | 1.51E-01 | -0.0035 | 5.91E-01 |
| cg02484673 | *JPH3* | 16 | 87674713 | -0.0029 | 8.04E-03 | -0.0059 | 2.67E-01 | -0.0018 | 7.26E-01 | 0.0197 | 7.74E-05 |
| cg05053718 | *ZNF469* | 16 | 88313910 | -0.0014 | 1.78E-01 | 0.0027 | 5.84E-01 | -0.0167 | 4.56E-04 | 0.0085 | 8.48E-02 |
| cg26747517 | *ANKRD11* | 16 | 89344383 | -0.0038 | 2.78E-04 | -0.0069 | 1.68E-01 | -0.0089 | 7.15E-02 | 0.0110 | 2.05E-02 |
| cg26217441 | *CPNE7* | 16 | 89643414 | 0.0005 | 7.82E-01 | -0.0014 | 8.64E-01 | -0.0071 | 3.85E-01 | 0.0237 | 3.45E-05 |
| cg19272348 | *SLC43A2* | 17 | 1521405 | -0.0012 | 5.43E-01 | -0.0096 | 3.28E-01 | -0.0355 | 1.95E-04 | -0.0013 | 8.94E-01 |
| cg18964590 | *SLC43A2* | 17 | 1521483 | -0.0023 | 2.59E-01 | -0.0103 | 2.83E-01 | -0.0389 | 3.34E-05 | -0.0066 | 4.94E-01 |
| cg00966411 | *DNAH2* | 17 | 7622819 | -0.0027 | 8.32E-03 | -0.0082 | 9.88E-02 | -0.0169 | 5.01E-04 | 0.0051 | 2.70E-01 |
| cg17993985 | *CHD3* | 17 | 7809534 | -0.0025 | 7.50E-03 | -0.0026 | 5.56E-01 | -0.0144 | 8.31E-04 | 0.0055 | 2.13E-01 |
| cg19774236 | *GUCY2D* | 17 | 7919022 | 0.0005 | 6.99E-01 | 0.0076 | 2.30E-01 | 0.0038 | 5.47E-01 | 0.0217 | 1.45E-04 |
| cg05388761 | *PIK3R5* | 17 | 8798664 | -0.0049 | 2.77E-04 | -0.0112 | 8.64E-02 | -0.0113 | 7.86E-02 | 0.0036 | 5.82E-01 |

| | | | | | | | | | | | |
|---|---|---|---|---|---|---|---|---|---|---|---|
| cg04653170 | *SPECC1* | 17 | 19958833 | -0.0045 | 2.97E-04 | -0.0059 | 3.17E-01 | 0.0005 | 9.30E-01 | 0.0056 | 3.55E-01 |
| cg24696861 | *TTLL6* | 17 | 46871582 | -0.0038 | 7.38E-05 | -0.0066 | 1.67E-01 | -0.0019 | 6.81E-01 | 0.0054 | 2.44E-01 |
| cg08806120 | *TEX14* | 17 | 56679904 | -0.0015 | 2.05E-01 | 0.0027 | 6.36E-01 | -0.0186 | 1.15E-03 | -0.0041 | 4.77E-01 |
| cg00250454 | *RGS9* | 17 | 63181565 | -0.0056 | 7.93E-06 | -0.0009 | 8.81E-01 | -0.0058 | 3.31E-01 | 0.0028 | 6.45E-01 |
| cg16594906 | *CACNG4* | 17 | 65026907 | 0.0017 | 1.03E-01 | 0.0204 | 5.40E-05 | -0.0003 | 9.59E-01 | 0.0091 | 6.70E-02 |
| cg26463800 | *DNAI2* | 17 | 72300792 | 0.0005 | 5.75E-01 | 0.0155 | 6.09E-04 | -0.0038 | 3.98E-01 | 0.0016 | 7.26E-01 |
| cg21997766 | *GRIN2C* | 17 | 72846113 | -0.0042 | 2.70E-04 | -0.0066 | 2.29E-01 | -0.0030 | 5.82E-01 | 0.0094 | 8.34E-02 |
| cg25277353 | *MGAT5B* | 17 | 74911529 | -0.0004 | 6.24E-01 | -0.0061 | 1.06E-01 | 0.0118 | 1.18E-03 | -0.0031 | 3.90E-01 |
| cg00802224 | *RBFOX3* | 17 | 77194129 | -0.0036 | 3.67E-04 | -0.0129 | 8.58E-03 | -0.0094 | 5.24E-02 | 0.0004 | 9.41E-01 |
| cg07307078 | *TUBB6* | 18 | 12307630 | -0.0004 | 7.97E-01 | -0.0005 | 9.41E-01 | 0.0049 | 4.86E-01 | 0.0222 | 6.84E-05 |
| cg03722909 | *SETBP1* | 18 | 42324965 | 0.0009 | 3.53E-01 | 0.0180 | 1.98E-04 | -0.0035 | 4.71E-01 | -0.0006 | 9.10E-01 |
| cg26571002 | *SIGLEC15* | 18 | 43416018 | 0.0002 | 8.44E-01 | -0.0030 | 6.24E-01 | -0.0029 | 6.22E-01 | 0.0215 | 3.32E-05 |
| cg26772788 | *GALR1* | 18 | 75609054 | -0.0003 | 7.74E-01 | 0.0009 | 8.63E-01 | -0.0014 | 7.93E-01 | 0.0198 | 1.29E-04 |
| cg25803139 | *GNG7* | 19 | 2539626 | -0.0032 | 1.06E-04 | 0.0017 | 6.74E-01 | -0.0007 | 8.53E-01 | -0.0012 | 7.59E-01 |
| cg08603735 | *APBA3* | 19 | 3754256 | -0.0005 | 5.59E-01 | 0.0029 | 4.72E-01 | -0.0144 | 2.60E-04 | 0.0070 | 8.02E-02 |
| cg04356699 | *ARRDC5* | 19 | 4888701 | -0.0012 | 1.12E-01 | 0.0041 | 2.75E-01 | -0.0125 | 7.19E-04 | 0.0015 | 6.89E-01 |
| cg13620110 | *EIF3G* | 19 | 10226628 | -0.0035 | 6.07E-03 | -0.0063 | 3.03E-01 | -0.0195 | 1.11E-03 | -0.0071 | 2.49E-01 |
| cg03722295 | *CDC37* | 19 | 10519375 | -0.0023 | 1.01E-01 | -0.0246 | 3.76E-04 | 0.0102 | 1.30E-01 | -0.0009 | 8.88E-01 |
| cg18685158 | *SMARCA4* | 19 | 11168915 | -0.0030 | 2.94E-04 | -0.0092 | 2.12E-02 | -0.0014 | 7.28E-01 | 0.0077 | 5.56E-02 |
| cg16512895 | *CACNA1A* | 19 | 13410117 | 0.0006 | 5.11E-01 | -0.0022 | 6.02E-01 | 0.0137 | 7.90E-04 | 0.0076 | 6.07E-02 |
| cg06596711 | *ARMC6* | 19 | 19148759 | -0.0009 | 3.73E-01 | 0.0015 | 7.55E-01 | -0.0176 | 2.99E-04 | -0.0022 | 6.53E-01 |
| cg10743390 | *VSTM2B* | 19 | 30020728 | 0.0022 | 7.99E-02 | 0.0208 | 4.89E-04 | 0.0042 | 4.75E-01 | -0.0067 | 2.64E-01 |
| cg27139805 | *DMKN* | 19 | 35992791 | -0.0028 | 7.16E-02 | -0.0024 | 7.56E-01 | 0.0007 | 9.22E-01 | 0.0233 | 8.78E-05 |
| cg14946950 | *GAPDHS* | 19 | 36023909 | 0.0012 | 5.24E-01 | 0.0120 | 1.93E-01 | 0.0320 | 4.36E-04 | -0.0026 | 7.82E-01 |
| cg15586563 | *TEX101* | 19 | 43916594 | 0.0021 | 9.38E-02 | 0.0206 | 5.37E-04 | 0.0083 | 1.58E-01 | -0.0032 | 5.92E-01 |
| cg27175069 | *ZNF649* | 19 | 52395190 | -0.0035 | 8.33E-05 | 0.0016 | 6.98E-01 | -0.0052 | 2.10E-01 | -0.0045 | 2.93E-01 |
| cg18236936 | *ZNF417* | 19 | 58419776 | 0.0002 | 8.69E-01 | 0.0176 | 2.16E-04 | 0.0014 | 7.67E-01 | -0.0027 | 5.80E-01 |
| cg12609362 | *PTPRA* | 20 | 2850640 | 0.0012 | 1.41E-01 | 0.0075 | 5.21E-02 | -0.0034 | 3.69E-01 | 0.0146 | 1.44E-04 |
| cg04368843 | *ADAM33* | 20 | 3660815 | -0.0002 | 9.46E-01 | 0.0084 | 4.91E-01 | -0.0083 | 4.90E-01 | 0.0258 | 2.10E-05 |
| cg24763243 | *HSPA12B* | 20 | 3713506 | -0.0007 | 1.17E-01 | 0.0004 | 8.73E-01 | -0.0073 | 7.16E-04 | 0.0023 | 3.00E-01 |
| cg27178677 | *PLCB1* | 20 | 8834803 | -0.0052 | 5.61E-05 | -0.0153 | 2.15E-02 | -0.0003 | 9.58E-01 | -0.0028 | 6.42E-01 |
| cg11806178 | *MAFB* | 20 | 39319929 | -0.0008 | 7.04E-01 | 0.0021 | 8.43E-01 | 0.0097 | 3.21E-01 | 0.0372 | 6.62E-05 |
| cg04575427 | *SPINT3* | 20 | 44144711 | -0.0018 | 3.75E-02 | -0.0149 | 3.94E-04 | 0.0043 | 2.90E-01 | 0.0046 | 2.71E-01 |
| cg07612655 | *PTGIS* | 20 | 48185517 | -0.0014 | 1.87E-01 | -0.0005 | 9.21E-01 | -0.0187 | 1.63E-04 | 0.0087 | 8.30E-02 |
| cg03501387 | *SNAI1* | 20 | 48601882 | -0.0034 | 7.82E-05 | -0.0065 | 1.15E-01 | -0.0082 | 4.28E-02 | 0.0039 | 3.44E-01 |
| cg11400707 | *SLCO4A1* | 20 | 61318513 | 0.0003 | 7.85E-01 | 0.0006 | 8.92E-01 | -0.0031 | 4.89E-01 | 0.0174 | 6.43E-05 |
| cg08989212 | *LINC00160* | 21 | 36096051 | -0.0031 | 1.98E-04 | -0.0071 | 7.81E-02 | -0.0035 | 3.76E-01 | -0.0029 | 4.56E-01 |
| cg01907098 | *PDXK* | 21 | 45149141 | 0.0015 | 2.66E-01 | 0.0056 | 3.71E-01 | 0.0022 | 7.18E-01 | 0.0196 | 3.66E-05 |
| cg25563983 | *CECR7* | 22 | 17517329 | -0.0024 | 9.16E-02 | -0.0231 | 6.57E-04 | 0.0046 | 4.96E-01 | -0.0004 | 9.57E-01 |
| cg23469117 | *YDJC* | 22 | 21985646 | -0.0026 | 1.78E-02 | -0.0181 | 5.52E-04 | 0.0052 | 3.23E-01 | 0.0132 | 1.16E-02 |
| cg24824734 | *SLC2A11* | 22 | 24195377 | -0.0021 | 4.91E-02 | 0.0084 | 1.01E-01 | -0.0184 | 2.55E-04 | 0.0031 | 5.55E-01 |
| cg18617091 | *TMPRSS6* | 22 | 37495019 | -0.0052 | 1.29E-04 | -0.0094 | 1.53E-01 | -0.0087 | 1.78E-01 | 0.0029 | 6.40E-01 |
| cg00191625 | *CACNA1I* | 22 | 40021996 | 0.0006 | 5.94E-01 | -0.0081 | 1.56E-01 | 0.0022 | 6.97E-01 | 0.0207 | 8.94E-05 |
| cg04078118 | *TNRC6B* | 22 | 40439969 | -0.0028 | 5.09E-02 | -0.0025 | 7.20E-01 | -0.0241 | 3.50E-04 | 0.0006 | 9.29E-01 |
| cg22650942 | *NDUFA6-AS1* | 22 | 42527158 | -0.0045 | 1.34E-04 | 0.0019 | 7.38E-01 | -0.0084 | 1.33E-01 | 0.0094 | 8.79E-02 |
| cg26799416 | *TCF20* | 22 | 42611379 | 0.0013 | 1.21E-01 | 0.0134 | 6.13E-04 | 0.0066 | 9.21E-02 | 0.0033 | 4.04E-01 |
| cg07349890 | *RRP7A* | 22 | 42913926 | 0.0032 | 8.69E-05 | 0.0118 | 3.61E-03 | 0.0060 | 1.34E-01 | 0.0066 | 1.04E-01 |
| cg20209891 | *MPPED1* | 22 | 43806614 | 0.0007 | 5.92E-01 | 0.0035 | 5.72E-01 | -0.0033 | 5.88E-01 | 0.0231 | 1.94E-04 |
| cg07366660 | *UPK3A* | 22 | 45680498 | 0.0006 | 5.91E-01 | 0.0060 | 2.27E-01 | -0.0016 | 7.46E-01 | 0.0196 | 6.45E-05 |
| cg25141264 | *TBC1D22A* | 22 | 47560578 | 0.0013 | 4.33E-01 | 0.0046 | 5.48E-01 | 0.0092 | 2.24E-01 | 0.0212 | 1.40E-04 |
| cg07675700 | *MIR3201* | 22 | 48728901 | 0.0011 | 3.88E-01 | -0.0047 | 4.57E-01 | 0.0031 | 6.21E-01 | 0.0238 | 1.32E-04 |

ETV: Exposure to violence. ETGV: Exposure to gun violence. CCDS: Checklist of Children's Distress Symptoms. PSS: Perceived Stress Scale.

**Table S3. Top 100 CpG sites from each of the four ETV/stress EWAS (total 336 CpG sites = 400 CpGs – 4 CpGs overlapped in two EWAS – 60 CpGs missing in Viva) and their corresponding results of atopic asthma in both separate and meta-analyses**

| CpG | Gene | CHR | Position | Beta (EVAPR) | *P* (EVAPR) | Beta (Viva) | *P* (Viva) | Beta (PIAMA) | *P* (PIAMA) | Beta Meta | *P* Meta | *FDR-P* Meta |
|---|---|---|---|---|---|---|---|---|---|---|---|---|
| cg02695349 | *STARD3NL* | chr7 | 38269241 | 0.0205 | 1.04E-02 | 0.0488 | 1.10E-05 | 0.0138 | 7.44E-02 | 0.0237 | 1.40E-06 | **2.80E-04** |
| cg04990977 | *SLC35F4* | chr14 | 58221774 | -0.0478 | 4.67E-06 | -0.0134 | 1.87E-02 | -0.0170 | 2.62E-02 | -0.0199 | 1.66E-06 | **2.80E-04** |
| cg18146152 | *TSR3* | chr16 | 1400772 | -0.0042 | 5.24E-01 | 0.0073 | 8.90E-03 | 0.0131 | 5.79E-05 | 0.0084 | 2.62E-05 | **2.93E-03** |
| cg03541903 | *CDC42SE2* | chr5 | 130651434 | 0.0071 | 5.39E-01 | 0.0095 | 8.64E-03 | 0.0209 | 1.90E-03 | 0.0117 | 1.24E-04 | **1.05E-02** |
| cg21376795 | *KLHL25* | chr15 | 86302121 | 0.0050 | 3.25E-01 | 0.0027 | 4.01E-01 | 0.0154 | 7.41E-06 | 0.0080 | 1.74E-04 | **1.17E-02** |
| cg27178677 | *PLCB1* | chr20 | 8834803 | -0.0328 | 3.60E-05 | 0.0028 | 7.65E-01 | -0.0119 | 2.03E-02 | -0.0144 | 2.30E-04 | **1.29E-02** |
| cg17076485 | *BUD13* | chr11 | 116580712 | -0.0112 | 1.18E-01 | -0.0014 | 8.88E-01 | -0.0225 | 2.28E-04 | -0.0150 | 4.09E-04 | **1.96E-02** |
| cg17335499 | *OR2B3* | chr6 | 29054325 | -0.0179 | 9.49E-03 | -0.0085 | 1.07E-02 | -0.0040 | 2.90E-01 | -0.0078 | 7.54E-04 | **3.13E-02** |
| cg26772788 | *GALR1* | chr18 | 75609054 | 0.0042 | 4.81E-01 | 0.0045 | 3.84E-01 | 0.0224 | 3.35E-05 | 0.0106 | 8.39E-04 | **3.13E-02** |
| cg03729152 | *TMEM196* | chr7 | 19961027 | 0.0247 | 3.27E-03 | 0.0054 | 2.28E-01 | 0.0148 | 2.00E-02 | 0.0110 | 9.56E-04 | **3.15E-02** |
| cg01039401 | *TEAD4* | chr12 | 3070406 | -0.0612 | 1.80E-05 | 0.0036 | 8.14E-01 | -0.0192 | 1.15E-01 | -0.0260 | 1.03E-03 | **3.15E-02** |
| cg16848072 | *ANAPC13* | chr3 | 134182665 | 0.0074 | 1.91E-01 | 0.0044 | 1.58E-01 | 0.0100 | 6.30E-03 | 0.0068 | 1.72E-03 | **4.80E-02** |
| cg16312872 | *TULP4* | chr6 | 158923181 | 0.0115 | 4.91E-02 | 0.0053 | 8.72E-02 | 0.0078 | 6.01E-02 | 0.0070 | 2.11E-03 | 5.44E-02 |
| cg15080232 | *TM4SF20* | chr2 | 228228336 | -0.0064 | 5.78E-01 | 0.0061 | 8.45E-03 | 0.0122 | 5.00E-02 | 0.0064 | 2.47E-03 | 5.60E-02 |
| cg17200999 | *SYNPO2* | chr4 | 119902588 | -0.0452 | 2.65E-03 | -0.0087 | 1.22E-01 | -0.0128 | 9.10E-02 | -0.0130 | 2.50E-03 | 5.60E-02 |
| cg12556325 | *LRRC61* | chr7 | 150026731 | 0.0201 | 9.27E-03 | 0.0215 | 1.13E-02 | 0.0010 | 8.88E-01 | 0.0127 | 3.70E-03 | 7.78E-02 |
| cg22601058 | *LOC283070* | chr10 | 12874207 | -0.0373 | 1.03E-02 | -0.0053 | 2.97E-01 | -0.0253 | 6.78E-03 | -0.0121 | 4.25E-03 | 8.41E-02 |
| cg12433084 | *GDF10* | chr10 | 48430289 | -0.0250 | 8.69E-02 | -0.0092 | 7.13E-02 | -0.0086 | 9.43E-02 | -0.0098 | 4.98E-03 | 9.09E-02 |
| cg24314877 | *MYC* | chr8 | 128656055 | -0.0130 | 6.07E-02 | -0.0035 | 4.17E-01 | -0.0104 | 2.10E-02 | -0.0079 | 5.71E-03 | 9.09E-02 |
| cg09825429 | *ZC3H15* | chr2 | 187385867 | 0.0000 | 9.99E-01 | 0.0023 | 1.37E-01 | 0.0141 | 1.33E-04 | 0.0037 | 5.86E-03 | 9.09E-02 |
| cg10169812 | *CD38* | chr4 | 15779392 | 0.0067 | 1.97E-01 | 0.0025 | 4.88E-02 | 0.0046 | 8.31E-02 | 0.0031 | 5.87E-03 | 9.09E-02 |
| cg18964590 | *SLC43A2* | chr17 | 1521483 | 0.0015 | 8.99E-01 | 0.0205 | 5.82E-03 | 0.0138 | 2.00E-01 | 0.0147 | 5.96E-03 | 9.09E-02 |
| cg08792416 | *SH3BGRL2* | chr6 | 80404639 | 0.0040 | 3.85E-01 | 0.0036 | 2.00E-01 | 0.0093 | 9.39E-03 | 0.0055 | 6.23E-03 | 9.09E-02 |
| cg00397575 | *RASL12* | chr15 | 65359932 | -0.0072 | 3.87E-02 | -0.0001 | 9.89E-01 | -0.0142 | 1.57E-02 | -0.0072 | 7.20E-03 | 1.00E-01 |
| cg24696861 | *TTLL6* | chr17 | 46871582 | -0.0172 | 2.25E-03 | -0.0079 | 4.28E-02 | 0.0028 | 6.00E-01 | -0.0073 | 7.48E-03 | 1.00E-01 |
| cg04388919 | *ZNF777* | chr7 | 149119133 | -0.0044 | 6.21E-01 | -0.0146 | 2.60E-01 | -0.0236 | 5.12E-03 | -0.0146 | 8.53E-03 | 1.02E-01 |
| cg01266390 | *BUB3* | chr10 | 125034002 | -0.0229 | 2.62E-02 | -0.0081 | 6.21E-01 | -0.0160 | 1.25E-01 | -0.0176 | 8.55E-03 | 1.02E-01 |
| cg19272348 | *SLC43A2* | chr17 | 1521405 | 0.0051 | 6.52E-01 | 0.0211 | 2.14E-02 | 0.0158 | 1.18E-01 | 0.0152 | 8.76E-03 | 1.02E-01 |
| cg01832757 | *FNDC1* | chr6 | 159547894 | -0.0235 | 1.55E-02 | -0.0023 | 5.71E-01 | -0.0122 | 1.45E-02 | -0.0078 | 8.81E-03 | 1.02E-01 |
| cg17701991 | *CHN2* | chr7 | 29305699 | -0.0056 | 3.25E-01 | -0.0081 | 4.37E-02 | -0.0054 | 1.64E-01 | -0.0065 | 9.19E-03 | 1.03E-01 |
| cg21924070 | *SPSB4* | chr3 | 140788453 | 0.0008 | 8.55E-01 | 0.0040 | 3.95E-02 | 0.0053 | 8.07E-02 | 0.0040 | 9.53E-03 | 1.03E-01 |
| cg17445948 | *LINC01226* | chr1 | 31997420 | 0.0054 | 5.63E-01 | 0.0054 | 2.27E-02 | 0.0074 | 3.03E-01 | 0.0056 | 9.99E-03 | 1.05E-01 |
| cg13620110 | *EIF3G* | chr19 | 10226628 | 0.0040 | 5.74E-01 | 0.0053 | 6.36E-01 | 0.0111 | 9.13E-03 | 0.0089 | 1.08E-02 | 1.07E-01 |
| cg14392168 | *GAL3ST4* | chr7 | 99766665 | -0.0617 | 7.04E-05 | -0.0132 | 2.53E-01 | -0.0050 | 4.69E-01 | -0.0141 | 1.09E-02 | 1.07E-01 |
| cg04368843 | *ADAM33* | chr20 | 3660815 | -0.0222 | 1.37E-01 | -0.0111 | 1.16E-01 | -0.0289 | 9.81E-02 | -0.0149 | 1.23E-02 | 1.18E-01 |
| cg08032483 | *CLCN7* | chr16 | 1495363 | -0.0037 | 5.32E-01 | 0.0084 | 2.52E-01 | 0.0111 | 2.89E-03 | 0.0072 | 1.36E-02 | 1.24E-01 |
| cg12037784 | *SH3BP4* | chr2 | 235962605 | 0.0033 | 5.48E-01 | 0.0058 | 2.38E-02 | 0.0033 | 3.65E-01 | 0.0048 | 1.41E-02 | 1.24E-01 |
| cg12283658 | *WDR60* | chr7 | 158702581 | -0.0003 | 9.63E-01 | 0.0025 | 2.75E-01 | 0.0112 | 2.29E-03 | 0.0046 | 1.43E-02 | 1.24E-01 |
| cg25876057 | *CCDC168* | chr13 | 103387002 | 0.0055 | 5.77E-01 | 0.0055 | 9.46E-02 | 0.0105 | 6.49E-02 | 0.0066 | 1.47E-02 | 1.24E-01 |
| cg27139805 | *DMKN* | chr19 | 35992791 | -0.0299 | 1.42E-03 | -0.0096 | 4.60E-02 | 0.0066 | 3.96E-01 | -0.0091 | 1.48E-02 | 1.24E-01 |
| cg13075951 | *KCNK16* | chr6 | 39282393 | 0.0342 | 4.57E-02 | 0.0139 | 6.96E-02 | 0.0007 | 9.79E-01 | 0.0163 | 1.57E-02 | 1.29E-01 |
| cg23626986 | *TMEM170B* | chr6 | 11487376 | -0.0014 | 7.67E-01 | 0.0018 | 3.87E-01 | 0.0072 | 2.62E-03 | 0.0036 | 1.66E-02 | 1.32E-01 |
| cg15678392 | *TMEM182* | chr2 | 103378135 | 0.0100 | 4.37E-01 | 0.0055 | 1.09E-01 | 0.0207 | 2.69E-02 | 0.0074 | 1.69E-02 | 1.32E-01 |
| cg05072559 | *SNAI2* | chr8 | 49813855 | -0.0137 | 2.73E-01 | 0.0061 | 2.87E-01 | 0.0190 | 2.69E-03 | 0.0092 | 2.14E-02 | 1.63E-01 |
| cg19482754 | *TRPV4* | chr12 | 110224547 | 0.0118 | 4.18E-02 | 0.0007 | 8.14E-01 | 0.0083 | 2.19E-02 | 0.0048 | 2.39E-02 | 1.78E-01 |
| cg13996593 | *DNER* | chr2 | 230574596 | -0.0514 | 1.41E-03 | -0.0053 | 3.83E-01 | -0.0092 | 2.47E-01 | -0.0104 | 2.44E-02 | 1.78E-01 |
| cg13483797 | *PRRX2* | chr9 | 132431581 | -0.0133 | 1.09E-01 | 0.0029 | 8.77E-01 | -0.0201 | 6.83E-02 | -0.0136 | 2.86E-02 | 2.01E-01 |
| cg09296697 | *MSI1* | chr12 | 120833809 | -0.0084 | 1.66E-01 | 0.0052 | 3.52E-03 | -0.0004 | 8.99E-01 | 0.0033 | 3.04E-02 | 2.01E-01 |
| cg21997766 | *GRIN2C* | chr17 | 72846113 | 0.0136 | 4.05E-02 | -0.0010 | 7.67E-01 | 0.0110 | 6.20E-03 | 0.0052 | 3.05E-02 | 2.01E-01 |
| cg06278714 | *HECW1* | chr7 | 43287968 | -0.0112 | 1.07E-01 | 0.0025 | 6.96E-01 | -0.0129 | 1.72E-02 | -0.0076 | 3.08E-02 | 2.01E-01 |
| cg00661205 | *VWA8* | chr13 | 42400902 | 0.0048 | 4.48E-01 | 0.0017 | 7.41E-01 | 0.0125 | 1.20E-02 | 0.0068 | 3.10E-02 | 2.01E-01 |
| cg04078118 | *TNRC6B* | chr22 | 40439969 | 0.0039 | 6.47E-01 | 0.0060 | 3.60E-01 | 0.0080 | 5.05E-02 | 0.0069 | 3.10E-02 | 2.01E-01 |
| cg03501387 | *SNAI1* | chr20 | 48601882 | 0.0113 | 2.97E-02 | 0.0032 | 2.01E-01 | 0.0025 | 4.93E-01 | 0.0041 | 3.19E-02 | 2.02E-01 |
| cg23028654 | *GRIP2* | chr3 | 14597729 | -0.0072 | 1.71E-01 | -0.0056 | 4.77E-01 | -0.0093 | 1.33E-01 | -0.0076 | 3.39E-02 | 2.08E-01 |
| cg04471092 | *SMOC2* | chr6 | 169246889 | -0.0029 | 5.21E-01 | 0.0052 | 1.38E-01 | 0.0111 | 8.18E-03 | 0.0049 | 3.41E-02 | 2.08E-01 |
| cg04732596 | *MEIS1* | chr2 | 66789740 | 0.0109 | 4.50E-01 | 0.0060 | 1.17E-01 | 0.0380 | 3.38E-02 | 0.0075 | 3.50E-02 | 2.10E-01 |
| cg04356699 | *ARRDC5* | chr19 | 4888701 | 0.0062 | 1.63E-01 | 0.0030 | 2.56E-01 | 0.0041 | 2.30E-01 | 0.0039 | 3.81E-02 | 2.25E-01 |
| cg15950251 | *TBXAS1* | chr7 | 139658345 | 0.0194 | 1.65E-02 | 0.0059 | 1.18E-01 | -0.0001 | 9.82E-01 | 0.0057 | 4.45E-02 | 2.55E-01 |
| cg07147958 | *LINC00536* | chr8 | 116822442 | -0.0123 | 2.11E-01 | 0.0081 | 6.98E-02 | 0.0318 | 6.03E-03 | 0.0076 | 4.64E-02 | 2.55E-01 |
| cg00402668 | *HLA-DOB* | chr6 | 32768079 | -0.0148 | 8.31E-03 | 0.0029 | 7.55E-02 | 0.0087 | 9.54E-03 | 0.0028 | 4.68E-02 | 2.55E-01 |
| cg09154166 | *SLC35G1* | chr10 | 95608979 | 0.0137 | 1.36E-02 | -0.0003 | 9.43E-01 | 0.0055 | 1.19E-01 | 0.0045 | 4.75E-02 | 2.55E-01 |
| cg14220055 | *LOC158434* | chr9 | 98885705 | -0.0242 | 7.19E-04 | -0.0019 | 3.20E-01 | -0.0036 | 4.45E-01 | -0.0034 | 4.75E-02 | 2.55E-01 |
| cg08986871 | *GIGYF1* | chr7 | 100279334 | -0.0003 | 9.58E-01 | 0.0019 | 1.54E-01 | 0.0067 | 5.01E-02 | 0.0023 | 4.78E-02 | 2.55E-01 |

| | | | | | | | | | | | | |
|---|---|---|---|---|---|---|---|---|---|---|---|---|
| cg18883643 | *ZNF512* | chr2 | 27845233 | -0.0104 | 3.83E-02 | 0.0015 | 1.51E-01 | 0.0059 | 1.04E-02 | 0.0018 | 5.22E-02 | 2.73E-01 |
| cg17025642 | *TRIM5* | chr11 | 5709468 | -0.0097 | 3.96E-01 | 0.0093 | 3.58E-02 | 0.0158 | 2.48E-01 | 0.0076 | 5.30E-02 | 2.73E-01 |
| cg05537316 | *SEMA7A* | chr15 | 74706314 | -0.0074 | 1.45E-01 | -0.0070 | 5.02E-02 | -0.0004 | 9.16E-01 | -0.0042 | 5.40E-02 | 2.73E-01 |
| cg04921109 | *PLEKHD1* | chr14 | 69952104 | 0.0268 | 2.57E-03 | 0.0105 | 4.05E-01 | -0.0078 | 4.33E-01 | 0.0113 | 5.44E-02 | 2.73E-01 |
| cg16205352 | *PRPH* | chr12 | 49688176 | -0.0076 | 3.15E-01 | 0.0029 | 6.48E-01 | -0.0168 | 6.78E-03 | -0.0073 | 5.62E-02 | 2.78E-01 |
| cg18783033 | *LIPK* | chr10 | 90488297 | -0.0209 | 9.68E-02 | -0.0012 | 7.81E-01 | -0.0146 | 2.85E-02 | -0.0066 | 6.25E-02 | 3.03E-01 |
| cg16577132 | *STRN3* | chr14 | 31408344 | 0.0038 | 5.84E-01 | 0.0018 | 2.04E-01 | 0.0086 | 5.98E-02 | 0.0025 | 6.31E-02 | 3.03E-01 |
| cg11927233 | *NPM1* | chr5 | 170816542 | 0.0170 | 4.67E-02 | 0.0088 | 5.28E-01 | 0.0040 | 5.82E-01 | 0.0094 | 6.81E-02 | 3.19E-01 |
| cg16097532 | *AHNAK2* | chr14 | 105416166 | 0.0030 | 7.48E-01 | 0.0353 | 1.57E-01 | 0.0127 | 8.84E-02 | 0.0102 | 6.96E-02 | 3.19E-01 |
| cg02484673 | *JPH3* | chr16 | 87674713 | -0.0208 | 1.23E-03 | -0.0099 | 2.12E-01 | 0.0004 | 9.01E-01 | -0.0052 | 6.98E-02 | 3.19E-01 |
| cg14685575 | *RTN1* | chr14 | 60193832 | -0.0077 | 1.24E-01 | -0.0035 | 1.99E-01 | -0.0015 | 7.24E-01 | -0.0038 | 7.04E-02 | 3.19E-01 |
| cg11302992 | *ELP4* | chr11 | 31789446 | -0.0043 | 5.78E-01 | 0.0033 | 7.95E-02 | 0.0091 | 2.88E-01 | 0.0031 | 7.58E-02 | 3.40E-01 |
| cg05314679 | *PABPC1* | chr8 | 101821998 | 0.0130 | 6.60E-02 | 0.0043 | 4.53E-01 | 0.0042 | 6.96E-01 | 0.0073 | 7.81E-02 | 3.45E-01 |
| cg01181415 | *LMO3* | chr12 | 16757954 | 0.0261 | 1.64E-01 | 0.0130 | 1.73E-01 | -0.0002 | 9.94E-01 | 0.0139 | 8.15E-02 | 3.56E-01 |
| cg22366698 | *ZNF195* | chr11 | 3400543 | 0.0059 | 3.22E-01 | 0.0062 | 6.40E-02 | -0.0013 | 8.01E-01 | 0.0044 | 8.34E-02 | 3.59E-01 |
| cg18189288 | *SPATS2L* | chr2 | 201262081 | 0.0123 | 1.71E-01 | 0.0022 | 5.60E-01 | 0.0131 | 7.65E-02 | 0.0053 | 8.72E-02 | 3.69E-01 |
| cg00988179 | *HMGCLL1* | chr6 | 55314478 | -0.0258 | 1.51E-02 | -0.0024 | 5.37E-01 | -0.0063 | 3.31E-01 | -0.0054 | 8.78E-02 | 3.69E-01 |
| cg13585930 | *NPFFR1* | chr10 | 72027357 | 0.0063 | 4.49E-01 | 0.0316 | 5.07E-03 | -0.0013 | 8.70E-01 | 0.0085 | 9.56E-02 | 3.97E-01 |
| cg19290938 | *MRPL23* | chr11 | 1988758 | -0.0221 | 6.99E-03 | -0.0029 | 2.46E-01 | 0.0059 | 4.93E-01 | -0.0038 | 9.92E-02 | 4.07E-01 |
| cg18838059 | *PDGFA* | chr7 | 534834 | 0.0067 | 3.20E-01 | -0.0004 | 8.88E-01 | 0.0071 | 3.01E-02 | 0.0035 | 1.04E-01 | 4.20E-01 |
| cg14375050 | *CYP2U1* | chr4 | 108848533 | -0.0058 | 5.49E-01 | 0.0018 | 4.10E-01 | 0.0162 | 4.90E-03 | 0.0032 | 1.09E-01 | 4.37E-01 |
| cg08381504 | *PRSS50* | chr3 | 46792357 | 0.0096 | 3.35E-01 | -0.0085 | 5.65E-01 | 0.0150 | 7.38E-02 | 0.0093 | 1.11E-01 | 4.40E-01 |
| cg16544989 | *GSE1* | chr16 | 85600639 | -0.0056 | 4.94E-01 | -0.0004 | 9.77E-01 | -0.0137 | 8.62E-02 | -0.0083 | 1.14E-01 | 4.44E-01 |
| cg09125924 | *PRKAG2* | chr7 | 151452190 | -0.0097 | 1.84E-01 | -0.0066 | 5.58E-02 | 0.0017 | 6.57E-01 | -0.0038 | 1.22E-01 | 4.67E-01 |
| cg11863854 | *RBMS3* | chr3 | 29471980 | -0.0144 | 5.37E-01 | 0.0043 | 6.15E-01 | -0.0290 | 4.70E-03 | -0.0098 | 1.22E-01 | 4.67E-01 |
| cg04575427 | *SPINT3* | chr20 | 44144711 | 0.0030 | 5.40E-01 | -0.0080 | 2.14E-02 | -0.0018 | 6.53E-01 | -0.0036 | 1.25E-01 | 4.71E-01 |
| cg21189146 | *TNXB* | chr6 | 32069841 | 0.0101 | 3.24E-02 | 0.0009 | 7.01E-01 | 0.0022 | 4.30E-01 | 0.0026 | 1.27E-01 | 4.74E-01 |
| cg03722295 | *CDC37* | chr19 | 10519375 | 0.0138 | 8.47E-02 | 0.0031 | 5.29E-01 | 0.0046 | 6.21E-01 | 0.0058 | 1.28E-01 | 4.74E-01 |
| cg20891116 | *BTNL2* | chr6 | 32363228 | -0.0040 | 5.10E-01 | 0.0037 | 1.54E-01 | 0.0044 | 2.46E-01 | 0.0030 | 1.30E-01 | 4.76E-01 |
| cg00306063 | *UNQ6494* | chr9 | 92289643 | 0.0053 | 2.38E-01 | 0.0010 | 8.18E-01 | 0.0035 | 2.65E-01 | 0.0034 | 1.36E-01 | 4.85E-01 |
| cg14260530 | *AKNA* | chr9 | 117157871 | -0.0056 | 4.34E-01 | -0.0151 | 5.10E-02 | 0.0024 | 7.76E-01 | -0.0066 | 1.37E-01 | 4.85E-01 |
| cg13674558 | *UBE3D* | chr6 | 83775899 | -0.0470 | 4.31E-03 | 0.0089 | 5.78E-01 | -0.0059 | 6.38E-01 | -0.0125 | 1.38E-01 | 4.85E-01 |
| cg17482424 | *BAMBI* | chr10 | 28970998 | 0.0029 | 6.93E-01 | 0.0017 | 3.99E-01 | 0.0094 | 7.26E-02 | 0.0027 | 1.38E-01 | 4.85E-01 |
| cg11806178 | *MAFB* | chr20 | 39319929 | -0.0180 | 1.30E-01 | 0.0031 | 6.78E-01 | -0.0230 | 3.30E-02 | -0.0079 | 1.44E-01 | 4.95E-01 |
| cg11947245 | *SLC46A3* | chr13 | 29328979 | -0.0121 | 3.37E-02 | -0.0007 | 9.19E-01 | -0.0016 | 7.29E-01 | -0.0046 | 1.46E-01 | 4.95E-01 |
| cg26430597 | *GLUL* | chr1 | 182354699 | 0.0118 | 1.90E-01 | 0.0028 | 5.17E-01 | 0.0095 | 3.09E-01 | 0.0052 | 1.46E-01 | 4.95E-01 |
| cg10625666 | *SLC39A10* | chr2 | 196521354 | -0.0225 | 1.65E-01 | -0.0043 | 4.84E-01 | -0.0118 | 3.48E-01 | -0.0076 | 1.51E-01 | 5.06E-01 |
| cg07720990 | *ADD2* | chr2 | 70998383 | -0.0086 | 3.57E-01 | -0.0042 | 3.01E-01 | -0.0030 | 5.38E-01 | -0.0042 | 1.55E-01 | 5.06E-01 |
| cg19547124 | *MOB2* | chr11 | 1669376 | -0.0038 | 6.10E-01 | -0.0029 | 4.75E-01 | -0.0100 | 1.56E-01 | -0.0045 | 1.56E-01 | 5.06E-01 |
| cg19064846 | *PTPRN2* | chr7 | 157273391 | -0.0245 | 1.23E-03 | 0.0004 | 9.49E-01 | 0.0007 | 9.02E-01 | -0.0051 | 1.57E-01 | 5.06E-01 |
| cg20991983 | *EPHA8* | chr1 | 22927528 | -0.0014 | 8.16E-01 | -0.0006 | 8.80E-01 | 0.0066 | 3.12E-02 | 0.0032 | 1.57E-01 | 5.06E-01 |
| cg17993985 | *CHD3* | chr17 | 7809534 | -0.0028 | 5.91E-01 | -0.0011 | 6.60E-01 | 0.0054 | 1.25E-02 | 0.0022 | 1.61E-01 | 5.15E-01 |
| cg20522006 | *MUC2* | chr11 | 1097259 | -0.0082 | 1.41E-01 | 0.0057 | 1.49E-01 | 0.0063 | 8.66E-02 | 0.0033 | 1.67E-01 | 5.30E-01 |
| cg00196474 | *EIF4E3* | chr3 | 71702735 | -0.0024 | 6.90E-01 | 0.0022 | 1.49E-01 | 0.0013 | 6.82E-01 | 0.0018 | 1.74E-01 | 5.46E-01 |
| cg25417917 | *RND3* | chr2 | 151452323 | -0.0166 | 7.08E-02 | 0.0046 | 4.11E-02 | -0.0028 | 6.57E-01 | 0.0028 | 1.75E-01 | 5.46E-01 |
| cg04368939 | *KCNIP2* | chr10 | 103588914 | -0.0063 | 3.10E-01 | -0.0006 | 9.27E-01 | -0.0170 | 1.27E-01 | -0.0058 | 1.78E-01 | 5.50E-01 |
| cg09801334 | *LTBP3* | chr11 | 65315625 | 0.0040 | 5.08E-01 | 0.0011 | 7.83E-01 | 0.0125 | 6.91E-02 | 0.0040 | 1.81E-01 | 5.50E-01 |
| cg12382418 | *METTL6* | chr3 | 15466952 | -0.0298 | 4.71E-03 | 0.0051 | 1.53E-01 | 0.0126 | 4.74E-02 | 0.0040 | 1.83E-01 | 5.50E-01 |
| cg00499831 | *CSMD2* | chr1 | 34627314 | -0.0136 | 3.68E-02 | -0.0012 | 7.81E-01 | -0.0019 | 7.25E-01 | -0.0039 | 1.83E-01 | 5.50E-01 |
| cg16358741 | *ST14* | chr11 | 130076257 | -0.0051 | 4.72E-01 | -0.0005 | 9.45E-01 | -0.0087 | 1.49E-01 | -0.0050 | 1.86E-01 | 5.51E-01 |
| cg08827764 | *TNIK* | chr3 | 171181416 | -0.0083 | 1.82E-01 | 0.0024 | 1.06E-01 | 0.0013 | 7.31E-01 | 0.0017 | 1.87E-01 | 5.51E-01 |
| cg18188150 | *PPRC1* | chr10 | 103895431 | -0.0062 | 4.39E-01 | 0.0053 | 2.43E-01 | 0.0074 | 1.76E-01 | 0.0042 | 1.89E-01 | 5.52E-01 |
| cg08603735 | *APBA3* | chr19 | 3754256 | -0.0024 | 5.93E-01 | 0.0036 | 1.32E-01 | 0.0028 | 4.66E-01 | 0.0024 | 1.91E-01 | 5.52E-01 |
| cg02546828 | *FAM49B* | chr8 | 130838883 | 0.0027 | 7.48E-01 | -0.0051 | 2.96E-01 | -0.0087 | 2.05E-01 | -0.0047 | 1.93E-01 | 5.55E-01 |
| cg11400707 | *SLCO4A1* | chr20 | 61318513 | -0.0032 | 5.75E-01 | -0.0031 | 2.96E-01 | 0.0132 | 1.80E-04 | 0.0027 | 1.96E-01 | 5.56E-01 |
| cg23349982 | *DIRC3* | chr2 | 218135498 | 0.0090 | 2.08E-01 | 0.0020 | 6.13E-01 | 0.0051 | 4.41E-01 | 0.0039 | 1.98E-01 | 5.56E-01 |
| cg01011757 | *EIF4E2* | chr2 | 233416425 | -0.0162 | 2.82E-03 | -0.0011 | 6.73E-01 | 0.0000 | 9.93E-01 | -0.0023 | 1.99E-01 | 5.56E-01 |
| cg11639168 | *POM121* | chr7 | 72393693 | -0.0106 | 2.35E-01 | 0.0022 | 1.96E-01 | 0.0065 | 3.13E-01 | 0.0020 | 2.02E-01 | 5.57E-01 |
| cg25141264 | *TBC1D22A* | chr22 | 47560578 | -0.0047 | 6.05E-01 | 0.0066 | 9.83E-02 | 0.0011 | 9.18E-01 | 0.0044 | 2.02E-01 | 5.57E-01 |
| cg11902863 | *TSSC1* | chr2 | 3044737 | 0.0251 | 3.38E-02 | 0.0055 | 5.84E-01 | 0.0010 | 8.69E-01 | 0.0062 | 2.04E-01 | 5.58E-01 |
| cg05857060 | *HIP1* | chr7 | 75267630 | -0.0285 | 7.55E-03 | 0.0016 | 7.36E-01 | -0.0156 | 1.41E-01 | -0.0050 | 2.06E-01 | 5.58E-01 |
| cg25803139 | *GNG7* | chr19 | 2539626 | -0.0010 | 8.23E-01 | 0.0037 | 6.52E-02 | -0.0011 | 7.59E-01 | 0.0020 | 2.08E-01 | 5.58E-01 |
| cg07410217 | *TBC1D1* | chr4 | 38153736 | -0.0052 | 3.42E-01 | 0.0009 | 6.93E-01 | 0.0061 | 3.41E-02 | 0.0022 | 2.09E-01 | 5.58E-01 |
| cg03034180 | *PACRG* | chr6 | 163721894 | 0.0070 | 1.95E-01 | 0.0036 | 6.72E-01 | 0.0017 | 8.10E-01 | 0.0047 | 2.17E-01 | 5.74E-01 |
| cg14445389 | *AFF3* | chr2 | 100426973 | -0.0702 | 8.36E-03 | 0.0045 | 2.83E-01 | -0.0374 | 1.83E-05 | -0.0046 | 2.19E-01 | 5.74E-01 |
| cg10300293 | *ERICH1* | chr8 | 643753 | -0.0046 | 4.06E-01 | -0.0115 | 6.18E-02 | 0.0010 | 8.16E-01 | -0.0036 | 2.28E-01 | 5.94E-01 |
| cg24763243 | *HSPA12B* | chr20 | 3713506 | -0.0033 | 2.00E-01 | -0.0019 | 5.90E-01 | -0.0001 | 9.84E-01 | -0.0022 | 2.32E-01 | 5.95E-01 |
| cg07321354 | *DLX2* | chr2 | 173078588 | -0.0117 | 2.30E-01 | -0.0019 | 3.93E-01 | -0.0036 | 6.32E-01 | -0.0025 | 2.33E-01 | 5.95E-01 |
| cg18003659 | *CHRNA1* | chr2 | 175629267 | -0.0067 | 3.82E-01 | -0.0024 | 2.52E-01 | 0.0003 | 9.44E-01 | -0.0022 | 2.35E-01 | 5.95E-01 |
| cg22650942 | *NDUFA6-AS1* | chr22 | 42527158 | -0.0014 | 8.36E-01 | 0.0094 | 1.65E-01 | 0.0078 | 3.61E-01 | 0.0049 | 2.36E-01 | 5.95E-01 |

| | | | | | | | | | | | | |
|---|---|---|---|---|---|---|---|---|---|---|---|---|
| cg08806120 | *TEX14* | chr17 | 56679904 | 0.0013 | 8.47E-01 | -0.0045 | 7.02E-02 | 0.0002 | 9.30E-01 | -0.0021 | 2.39E-01 | 5.95E-01 |
| cg11950778 | *UGT8* | chr4 | 115519006 | -0.0081 | 4.38E-01 | -0.0097 | 4.55E-01 | -0.0050 | 5.67E-01 | -0.0070 | 2.40E-01 | 5.95E-01 |
| cg19838358 | *ANKLE2* | chr12 | 133304061 | 0.0042 | 3.76E-01 | 0.0003 | 9.04E-01 | 0.0046 | 1.81E-01 | 0.0022 | 2.41E-01 | 5.95E-01 |
| cg26858238 | *DAXX* | chr6 | 33289208 | -0.0073 | 4.14E-01 | 0.0099 | 4.47E-02 | 0.0003 | 9.67E-01 | 0.0041 | 2.44E-01 | 5.98E-01 |
| cg26518884 | *CPE* | chr4 | 166322679 | -0.0078 | 2.36E-01 | 0.0039 | 2.64E-02 | -0.0038 | 2.52E-01 | 0.0017 | 2.46E-01 | 5.98E-01 |
| cg06827611 | *ITPRIPL1* | chr2 | 96993311 | 0.0033 | 4.66E-01 | -0.0001 | 9.83E-01 | 0.0069 | 1.16E-01 | 0.0026 | 2.48E-01 | 6.00E-01 |
| cg02104112 | *CCND1* | chr11 | 69418071 | -0.0152 | 1.17E-01 | -0.0008 | 7.46E-01 | -0.0095 | 1.66E-01 | -0.0027 | 2.51E-01 | 6.01E-01 |
| cg25204743 | *KCNQ1* | chr11 | 2715837 | 0.0001 | 9.95E-01 | 0.0118 | 1.57E-01 | 0.0054 | 6.25E-01 | 0.0061 | 2.52E-01 | 6.01E-01 |
| cg16264537 | *LOC284661* | chr1 | 4468051 | -0.0044 | 4.09E-01 | 0.0008 | 8.20E-01 | -0.0045 | 1.72E-01 | -0.0025 | 2.62E-01 | 6.18E-01 |
| cg27314569 | *BSX* | chr11 | 122852461 | -0.0045 | 3.59E-01 | -0.0022 | 3.81E-01 | -0.0003 | 9.47E-01 | -0.0023 | 2.63E-01 | 6.18E-01 |
| cg07461215 | *CNTNAP5* | chr2 | 124444215 | -0.0016 | 8.72E-01 | 0.0012 | 7.96E-01 | 0.0074 | 1.30E-01 | 0.0035 | 2.67E-01 | 6.24E-01 |
| cg01494441 | *CERKL* | chr2 | 182523146 | -0.0228 | 1.07E-02 | 0.0006 | 8.95E-01 | -0.0020 | 6.63E-01 | -0.0035 | 2.69E-01 | 6.24E-01 |
| cg10224088 | *S100A13* | chr1 | 153597891 | -0.0036 | 4.40E-01 | 0.0012 | 4.99E-01 | 0.0050 | 1.01E-01 | 0.0016 | 2.75E-01 | 6.33E-01 |
| cg02409090 | *EXOC2* | chr6 | 709725 | 0.0008 | 9.08E-01 | -0.0056 | 4.55E-02 | 0.0021 | 5.35E-01 | -0.0022 | 2.85E-01 | 6.48E-01 |
| cg12017061 | *ALOX5AP* | chr13 | 31271512 | 0.0082 | 4.71E-01 | 0.0054 | 6.15E-01 | 0.0059 | 5.28E-01 | 0.0063 | 2.87E-01 | 6.48E-01 |
| cg13804467 | *LINGO2* | chr9 | 28907222 | -0.0102 | 3.37E-01 | 0.0033 | 5.10E-01 | -0.0121 | 4.23E-02 | -0.0038 | 2.87E-01 | 6.48E-01 |
| cg15586563 | *TEX101* | chr19 | 43916594 | -0.0088 | 2.16E-01 | 0.0029 | 2.61E-01 | 0.0031 | 3.64E-01 | 0.0021 | 2.94E-01 | 6.53E-01 |
| cg22032961 | *FCAMR* | chr1 | 207152909 | -0.0251 | 2.96E-03 | 0.0087 | 1.85E-01 | -0.0038 | 4.76E-01 | -0.0039 | 2.95E-01 | 6.53E-01 |
| cg05376362 | *CD109* | chr6 | 74826447 | -0.0025 | 7.17E-01 | -0.0010 | 7.31E-01 | -0.0036 | 2.73E-01 | -0.0021 | 2.97E-01 | 6.53E-01 |
| cg08033935 | *C16orf58* | chr16 | 31505179 | -0.0038 | 5.49E-01 | 0.0033 | 2.33E-01 | 0.0036 | 5.28E-01 | 0.0024 | 2.99E-01 | 6.53E-01 |
| cg11869150 | *EXOC3* | chr5 | 465665 | -0.0042 | 4.31E-01 | 0.0026 | 5.54E-01 | 0.0076 | 8.10E-02 | 0.0028 | 3.01E-01 | 6.53E-01 |
| cg04653170 | *SPECC1* | chr17 | 19958833 | -0.0098 | 1.63E-01 | -0.0024 | 1.72E-01 | 0.0073 | 1.15E-01 | -0.0016 | 3.02E-01 | 6.53E-01 |
| cg14946950 | *GAPDHS* | chr19 | 36023909 | 0.0090 | 4.14E-01 | -0.0008 | 9.46E-01 | 0.0080 | 3.56E-01 | 0.0061 | 3.03E-01 | 6.53E-01 |
| cg08734395 | *MAT2B* | chr5 | 163086064 | -0.0122 | 1.79E-01 | -0.0006 | 9.27E-01 | -0.0031 | 5.79E-01 | -0.0039 | 3.10E-01 | 6.63E-01 |
| cg07006398 | *IRX6* | chr16 | 55406516 | -0.0238 | 1.06E-01 | -0.0047 | 6.41E-01 | -0.0005 | 9.60E-01 | -0.0061 | 3.19E-01 | 6.72E-01 |
| cg19513744 | *ARHGEF3* | chr3 | 56836209 | -0.0034 | 4.16E-01 | -0.0010 | 5.49E-01 | -0.0050 | 4.65E-01 | -0.0015 | 3.20E-01 | 6.72E-01 |
| cg00980804 | *RND3* | chr2 | 150977441 | -0.0029 | 6.61E-01 | 0.0082 | 1.19E-01 | 0.0021 | 7.60E-01 | 0.0035 | 3.20E-01 | 6.72E-01 |
| cg02552137 | *ITGA4* | chr2 | 182320901 | -0.0010 | 8.49E-01 | 0.0022 | 1.68E-01 | -0.0020 | 5.96E-01 | 0.0014 | 3.26E-01 | 6.72E-01 |
| cg02987054 | *INO80B-WBP1* | chr2 | 74680627 | 0.0018 | 7.67E-01 | 0.0018 | 2.46E-01 | -0.0005 | 8.66E-01 | 0.0013 | 3.31E-01 | 6.72E-01 |
| cg06119575 | *TAL2* | chr9 | 108424905 | 0.0012 | 8.39E-01 | 0.0005 | 7.36E-01 | 0.0069 | 7.67E-02 | 0.0013 | 3.32E-01 | 6.72E-01 |
| cg23839479 | *MYEOV* | chr11 | 69181986 | -0.0058 | 3.54E-01 | 0.0050 | 1.34E-01 | 0.0020 | 6.87E-01 | 0.0024 | 3.32E-01 | 6.72E-01 |
| cg02668819 | *TNXB* | chr6 | 32057053 | -0.0049 | 3.64E-01 | 0.0021 | 2.58E-01 | 0.0017 | 5.54E-01 | 0.0015 | 3.32E-01 | 6.72E-01 |
| cg09265016 | *TBX3* | chr12 | 115170515 | 0.0015 | 8.72E-01 | -0.0100 | 3.30E-01 | -0.0052 | 4.18E-01 | -0.0046 | 3.32E-01 | 6.72E-01 |
| cg08932683 | *RNFT2* | chr12 | 117253648 | 0.0041 | 5.36E-01 | -0.0027 | 2.57E-01 | -0.0014 | 6.50E-01 | -0.0017 | 3.34E-01 | 6.72E-01 |
| cg02749511 | *SP3* | chr2 | 174696522 | 0.0043 | 4.94E-01 | -0.0018 | 7.57E-01 | 0.0106 | 1.48E-01 | 0.0035 | 3.44E-01 | 6.88E-01 |
| cg00802224 | *RBFOX3* | chr17 | 77194129 | -0.0095 | 1.02E-01 | -0.0006 | 9.28E-01 | 0.0022 | 7.52E-01 | -0.0035 | 3.47E-01 | 6.90E-01 |
| cg19508967 | *NDUFS6* | chr5 | 1840347 | -0.0113 | 1.07E-01 | -0.0002 | 9.36E-01 | -0.0030 | 4.78E-01 | -0.0021 | 3.50E-01 | 6.91E-01 |
| cg08458678 | *SHANK2* | chr11 | 70565177 | -0.0070 | 1.90E-01 | -0.0012 | 8.03E-01 | -0.0008 | 8.49E-01 | -0.0025 | 3.55E-01 | 6.97E-01 |
| cg19942977 | *DLG2* | chr11 | 83394759 | -0.0130 | 5.81E-02 | -0.0013 | 6.47E-01 | 0.0000 | 9.94E-01 | -0.0019 | 3.60E-01 | 7.01E-01 |
| cg13550751 | *ABL2* | chr1 | 179194565 | -0.0151 | 1.52E-01 | 0.0022 | 2.57E-01 | 0.0018 | 7.73E-01 | 0.0016 | 3.62E-01 | 7.01E-01 |
| cg23184477 | *DOCK2* | chr5 | 169064897 | 0.0211 | 2.35E-01 | -0.0008 | 9.48E-01 | 0.0264 | 3.14E-01 | 0.0083 | 3.63E-01 | 7.01E-01 |
| cg19616851 | *LINC01005* | chr7 | 63363811 | -0.0236 | 7.00E-02 | 0.0306 | 7.65E-03 | -0.0169 | 3.79E-02 | -0.0053 | 3.65E-01 | 7.01E-01 |
| cg10236247 | *IL17D* | chr13 | 21276233 | -0.0076 | 2.37E-01 | -0.0012 | 6.15E-01 | 0.0133 | 8.31E-04 | 0.0018 | 3.67E-01 | 7.01E-01 |
| cg18617091 | *TMPRSS6* | chr22 | 37495019 | 0.0102 | 1.95E-01 | -0.0019 | 8.20E-01 | 0.0052 | 6.63E-01 | 0.0045 | 3.76E-01 | 7.14E-01 |
| cg05388761 | *PIK3R5* | chr17 | 8798664 | -0.0094 | 2.46E-01 | -0.0042 | 2.65E-01 | 0.0034 | 5.04E-01 | -0.0025 | 3.79E-01 | 7.16E-01 |
| cg17255108 | *PTPRN2* | chr7 | 157338433 | -0.0093 | 1.38E-01 | 0.0053 | 3.42E-01 | -0.0056 | 2.90E-01 | -0.0029 | 3.84E-01 | 7.21E-01 |
| cg24545125 | *TNP1* | chr2 | 217724866 | 0.0005 | 9.49E-01 | -0.0014 | 6.96E-01 | 0.0121 | 2.54E-02 | 0.0024 | 3.92E-01 | 7.32E-01 |
| cg13150061 | *WDR20* | chr14 | 102685953 | -0.0013 | 8.29E-01 | -0.0013 | 7.48E-01 | 0.0080 | 6.44E-02 | 0.0023 | 3.99E-01 | 7.41E-01 |
| cg02501086 | *ADRA1A* | chr8 | 26717875 | -0.0001 | 9.79E-01 | 0.0010 | 8.31E-01 | 0.0037 | 2.97E-01 | 0.0021 | 4.04E-01 | 7.46E-01 |
| cg25277353 | *MGAT5B* | chr17 | 74911529 | -0.0026 | 5.50E-01 | 0.0034 | 2.16E-01 | 0.0012 | 6.93E-01 | 0.0015 | 4.09E-01 | 7.48E-01 |
| cg26799416 | *TCF20* | chr22 | 42611379 | -0.0010 | 8.49E-01 | 0.0005 | 5.80E-01 | 0.0031 | 2.68E-01 | 0.0007 | 4.11E-01 | 7.48E-01 |
| cg05134054 | *CUX2* | chr12 | 111751588 | 0.0020 | 7.52E-01 | -0.0067 | 6.36E-01 | -0.0152 | 1.00E-01 | -0.0040 | 4.18E-01 | 7.48E-01 |
| cg20982112 | *PVRL3* | chr3 | 110911278 | 0.0041 | 4.66E-01 | -0.0006 | 7.15E-01 | 0.0072 | 3.56E-02 | 0.0012 | 4.19E-01 | 7.48E-01 |
| cg23932798 | *MYOT* | chr5 | 137142029 | -0.0007 | 8.91E-01 | -0.0070 | 1.66E-01 | 0.0064 | 4.26E-02 | 0.0019 | 4.20E-01 | 7.48E-01 |
| cg17104151 | *C1QC* | chr1 | 22970132 | -0.0112 | 1.08E-01 | 0.0120 | 1.76E-01 | -0.0068 | 4.06E-01 | -0.0037 | 4.21E-01 | 7.48E-01 |
| cg00191625 | *CACNA1I* | chr22 | 40021996 | -0.0076 | 2.62E-01 | 0.0027 | 7.61E-01 | -0.0035 | 6.92E-01 | -0.0037 | 4.23E-01 | 7.48E-01 |
| cg00250454 | *RGS9* | chr17 | 63181565 | -0.0087 | 1.85E-01 | -0.0014 | 2.80E-01 | 0.0062 | 1.27E-01 | -0.0010 | 4.23E-01 | 7.48E-01 |
| cg23469117 | *YDJC* | chr22 | 21985646 | 0.0025 | 6.92E-01 | 0.0009 | 8.35E-01 | 0.0048 | 3.99E-01 | 0.0025 | 4.27E-01 | 7.51E-01 |
| cg15423862 | *KCNAB1* | chr3 | 155838109 | -0.0216 | 6.23E-03 | -0.0020 | 3.69E-01 | 0.0103 | 5.55E-02 | -0.0016 | 4.34E-01 | 7.56E-01 |
| cg16512895 | *CACNA1A* | chr19 | 13410117 | 0.0021 | 6.59E-01 | 0.0000 | 9.91E-01 | 0.0076 | 2.31E-01 | 0.0022 | 4.34E-01 | 7.56E-01 |
| cg20377447 | *UNC5CL* | chr6 | 41010217 | -0.0014 | 7.77E-01 | -0.0034 | 3.51E-01 | -0.0003 | 9.29E-01 | -0.0017 | 4.40E-01 | 7.57E-01 |
| cg06946314 | *SH3BP4* | chr2 | 236092957 | 0.0072 | 1.99E-01 | -0.0009 | 8.31E-01 | 0.0029 | 6.17E-01 | 0.0022 | 4.40E-01 | 7.57E-01 |
| cg03076855 | *CEP162* | chr6 | 85132322 | 0.0213 | 2.86E-01 | 0.0020 | 8.62E-01 | -0.0086 | 1.79E-01 | -0.0041 | 4.44E-01 | 7.58E-01 |
| cg21186708 | *SOX1* | chr13 | 112716728 | -0.0011 | 8.23E-01 | -0.0014 | 6.95E-01 | -0.0095 | 2.82E-01 | -0.0021 | 4.46E-01 | 7.58E-01 |
| cg19489704 | *EHD3* | chr2 | 31508053 | -0.0050 | 3.16E-01 | 0.0008 | 5.90E-01 | 0.0041 | 1.91E-01 | 0.0010 | 4.47E-01 | 7.58E-01 |
| cg22884082 | *SMIM8* | chr6 | 88039023 | 0.0025 | 7.30E-01 | 0.0015 | 5.77E-01 | 0.0022 | 6.86E-01 | 0.0017 | 4.52E-01 | 7.63E-01 |
| cg18552983 | *ASB18* | chr2 | 237128101 | -0.0012 | 8.18E-01 | -0.0001 | 9.79E-01 | 0.0052 | 1.24E-01 | 0.0014 | 4.58E-01 | 7.66E-01 |
| cg02801405 | *LOC100507384* | chr11 | 45727997 | -0.0123 | 4.00E-02 | 0.0005 | 9.10E-01 | 0.0028 | 6.29E-01 | -0.0023 | 4.58E-01 | 7.66E-01 |
| cg27300028 | *CLYBL* | chr13 | 100312810 | -0.0031 | 5.95E-01 | -0.0012 | 6.61E-01 | 0.0087 | 2.40E-02 | 0.0015 | 4.71E-01 | 7.77E-01 |
| cg08989212 | *LINC00160* | chr21 | 36096051 | 0.0052 | 2.71E-01 | 0.0000 | 9.93E-01 | 0.0020 | 6.01E-01 | 0.0014 | 4.71E-01 | 7.77E-01 |

| | | | | | | | | | | | | |
|---|---|---|---|---|---|---|---|---|---|---|---|---|
| cg26552163 | *IFT140* | chr16 | 1589859 | 0.0035 | 5.11E-01 | -0.0006 | 8.45E-01 | 0.0033 | 3.51E-01 | 0.0016 | 4.73E-01 | 7.77E-01 |
| cg13632655 | *PTPRN2* | chr7 | 158266097 | 0.0064 | 4.75E-01 | -0.0002 | 9.81E-01 | 0.0099 | 5.22E-01 | 0.0043 | 4.74E-01 | 7.77E-01 |
| cg26907313 | *ATP6V1B1* | chr2 | 71167117 | -0.0077 | 1.54E-01 | 0.0057 | 1.41E-01 | 0.0028 | 4.97E-01 | 0.0018 | 4.79E-01 | 7.81E-01 |
| cg06595154 | *MRVI1* | chr11 | 10716164 | -0.0389 | 5.66E-03 | 0.0023 | 6.88E-01 | -0.0020 | 7.72E-01 | -0.0029 | 4.81E-01 | 7.81E-01 |
| cg02235663 | *B3GALNT1* | chr3 | 160825607 | -0.0051 | 2.73E-01 | 0.0016 | 1.79E-01 | -0.0021 | 4.50E-01 | 0.0007 | 4.96E-01 | 8.01E-01 |
| cg15009913 | *MORN1* | chr1 | 2287741 | 0.0019 | 6.72E-01 | -0.0004 | 8.29E-01 | -0.0033 | 2.23E-01 | -0.0009 | 5.00E-01 | 8.04E-01 |
| cg07307078 | *TUBB6* | chr18 | 12307630 | -0.0024 | 7.74E-01 | 0.0065 | 4.26E-01 | -0.0144 | 8.76E-02 | -0.0032 | 5.03E-01 | 8.05E-01 |
| cg07611816 | *AGAP1* | chr2 | 236760371 | -0.0097 | 1.78E-01 | 0.0009 | 7.76E-01 | 0.0063 | 1.21E-01 | 0.0015 | 5.07E-01 | 8.07E-01 |
| cg12810264 | *BMS1* | chr10 | 43329092 | 0.0043 | 3.91E-01 | -0.0026 | 3.60E-01 | -0.0016 | 5.71E-01 | -0.0012 | 5.10E-01 | 8.08E-01 |
| cg08231096 | *THSD7B* | chr2 | 137748460 | 0.0038 | 6.36E-01 | -0.0005 | 8.85E-01 | 0.0047 | 2.83E-01 | 0.0016 | 5.21E-01 | 8.20E-01 |
| cg10629128 | *HDAC4* | chr2 | 240280038 | -0.0194 | 3.39E-02 | 0.0032 | 5.07E-01 | -0.0025 | 6.16E-01 | -0.0021 | 5.24E-01 | 8.20E-01 |
| cg07675700 | *MIR3201* | chr22 | 48728901 | -0.0131 | 7.89E-02 | 0.0012 | 8.92E-01 | 0.0035 | 6.09E-01 | -0.0028 | 5.24E-01 | 8.20E-01 |
| cg18685158 | *SMARCA4* | chr19 | 11168915 | -0.0003 | 9.45E-01 | 0.0008 | 7.44E-01 | 0.0023 | 4.76E-01 | 0.0011 | 5.38E-01 | 8.37E-01 |
| cg25247122 | *SOX1* | chr13 | 112652556 | -0.0068 | 3.17E-01 | 0.0017 | 5.64E-01 | 0.0037 | 3.63E-01 | 0.0014 | 5.41E-01 | 8.37E-01 |
| cg11892307 | *EXOC2* | chr6 | 470595 | 0.0055 | 3.09E-01 | -0.0023 | 4.52E-01 | -0.0039 | 3.67E-01 | -0.0014 | 5.43E-01 | 8.37E-01 |
| cg19234171 | *LOC90246* | chr3 | 128226071 | -0.0087 | 2.33E-01 | 0.0023 | 3.78E-01 | -0.0052 | 1.16E-01 | -0.0012 | 5.49E-01 | 8.42E-01 |
| cg16501235 | *C1orf54* | chr1 | 150245988 | -0.0109 | 7.46E-02 | 0.0025 | 4.12E-01 | -0.0020 | 4.38E-01 | -0.0011 | 5.52E-01 | 8.42E-01 |
| cg18034995 | *VPS41* | chr7 | 38805409 | -0.0086 | 2.42E-01 | -0.0010 | 5.04E-01 | 0.0052 | 3.16E-01 | -0.0008 | 5.57E-01 | 8.42E-01 |
| cg03537802 | *CEP126* | chr11 | 101868388 | -0.0052 | 5.90E-01 | 0.0026 | 4.15E-01 | 0.0006 | 8.77E-01 | 0.0014 | 5.58E-01 | 8.42E-01 |
| cg24775027 | *ZNF804A* | chr2 | 185459721 | -0.0218 | 2.22E-03 | 0.0022 | 2.11E-01 | 0.0032 | 6.54E-01 | 0.0010 | 5.59E-01 | 8.42E-01 |
| cg10327980 | *FITM1* | chr14 | 24601001 | -0.0074 | 2.38E-01 | 0.0069 | 5.85E-01 | 0.0066 | 1.57E-01 | 0.0021 | 5.68E-01 | 8.45E-01 |
| cg22481770 | *SSR3* | chr3 | 156324118 | 0.0004 | 9.64E-01 | -0.0060 | 5.73E-01 | -0.0084 | 5.34E-01 | -0.0034 | 5.70E-01 | 8.45E-01 |
| cg02865595 | *SH3D21* | chr1 | 36771714 | -0.0430 | 1.41E-02 | 0.0031 | 6.22E-01 | -0.0253 | 3.17E-01 | -0.0032 | 5.71E-01 | 8.45E-01 |
| cg27368726 | *CYP1B1-AS1* | chr2 | 38385687 | 0.0013 | 8.40E-01 | 0.0010 | 7.33E-01 | 0.0018 | 6.67E-01 | 0.0013 | 5.73E-01 | 8.45E-01 |
| cg04127998 | *IFT43* | chr14 | 76518992 | -0.0058 | 2.90E-01 | -0.0031 | 1.12E-01 | 0.0060 | 4.56E-02 | -0.0009 | 5.73E-01 | 8.45E-01 |
| cg23205775 | *HIST1H2AD* | chr6 | 26199576 | 0.0127 | 2.15E-02 | 0.0023 | 3.94E-01 | -0.0183 | 2.71E-03 | 0.0012 | 5.84E-01 | 8.57E-01 |
| cg26463800 | *DNAI2* | chr17 | 72300792 | -0.0113 | 3.84E-02 | 0.0027 | 1.94E-01 | 0.0006 | 8.13E-01 | 0.0008 | 5.95E-01 | 8.66E-01 |
| cg18236936 | *ZNF417* | chr19 | 58419776 | -0.0097 | 6.57E-02 | 0.0006 | 5.49E-01 | 0.0033 | 3.16E-01 | 0.0005 | 5.96E-01 | 8.66E-01 |
| cg26502105 | *FXYD6-FXYD2* | chr11 | 117701909 | 0.0035 | 5.27E-01 | -0.0018 | 7.04E-01 | 0.0055 | 3.90E-01 | 0.0016 | 5.98E-01 | 8.66E-01 |
| cg03102879 | *SCNN1B* | chr16 | 23313295 | 0.0013 | 8.07E-01 | 0.0016 | 2.93E-01 | -0.0133 | 2.69E-02 | 0.0007 | 6.05E-01 | 8.73E-01 |
| cg03722909 | *SETBP1* | chr18 | 42324965 | 0.0006 | 9.12E-01 | -0.0049 | 1.94E-02 | 0.0066 | 2.07E-02 | -0.0008 | 6.09E-01 | 8.74E-01 |
| cg19774236 | *GUCY2D* | chr17 | 7919022 | -0.0036 | 6.42E-01 | -0.0007 | 8.54E-01 | -0.0014 | 7.26E-01 | -0.0013 | 6.11E-01 | 8.74E-01 |
| cg06440348 | *PRDM8* | chr4 | 81111527 | -0.0065 | 3.73E-01 | 0.0201 | 9.24E-02 | 0.0048 | 5.26E-01 | 0.0024 | 6.17E-01 | 8.77E-01 |
| cg14896948 | *COBL* | chr7 | 51096885 | 0.0083 | 1.37E-01 | 0.0032 | 5.20E-01 | -0.0031 | 4.19E-01 | 0.0013 | 6.22E-01 | 8.77E-01 |
| cg09816180 | *GC* | chr4 | 72650088 | -0.0067 | 2.99E-01 | -0.0019 | 3.11E-01 | 0.0051 | 1.55E-01 | -0.0008 | 6.27E-01 | 8.77E-01 |
| cg22810546 | *PAIP1* | chr5 | 43559688 | -0.0137 | 1.40E-01 | 0.0015 | 8.32E-01 | 0.0033 | 7.42E-01 | -0.0024 | 6.29E-01 | 8.77E-01 |
| cg15217784 | *FAM83B* | chr6 | 54722227 | -0.0047 | 6.33E-01 | 0.0009 | 8.40E-01 | -0.0071 | 3.45E-01 | -0.0018 | 6.33E-01 | 8.77E-01 |
| cg00750481 | *SPRY2* | chr13 | 80755568 | 0.0125 | 6.10E-01 | -0.0001 | 9.98E-01 | 0.0056 | 8.78E-01 | 0.0090 | 6.35E-01 | 8.77E-01 |
| cg00966411 | *DNAH2* | chr17 | 7622819 | 0.0039 | 5.05E-01 | 0.0018 | 5.77E-01 | -0.0010 | 7.85E-01 | 0.0011 | 6.37E-01 | 8.77E-01 |
| cg02847897 | *KLHL8* | chr4 | 88142275 | 0.0092 | 4.49E-01 | -0.0204 | 3.28E-01 | 0.0098 | 5.39E-01 | 0.0041 | 6.38E-01 | 8.77E-01 |
| cg14905665 | *NRP2* | chr2 | 206661494 | 0.0075 | 1.99E-01 | -0.0001 | 9.51E-01 | 0.0013 | 6.64E-01 | 0.0007 | 6.46E-01 | 8.77E-01 |
| cg25090315 | *GPR68* | chr14 | 91705005 | -0.0078 | 1.31E-01 | 0.0032 | 3.92E-01 | 0.0023 | 4.19E-01 | 0.0009 | 6.46E-01 | 8.77E-01 |
| cg21449463 | *MRPL15* | chr8 | 55091759 | -0.0356 | 4.30E-02 | 0.0037 | 6.46E-01 | -0.0023 | 8.41E-01 | -0.0028 | 6.46E-01 | 8.77E-01 |
| cg27506254 | *XKR4* | chr8 | 56436472 | -0.0090 | 1.45E-01 | 0.0013 | 4.20E-01 | 0.0011 | 8.19E-01 | 0.0007 | 6.48E-01 | 8.77E-01 |
| cg24434987 | *REG3G* | chr2 | 79221130 | 0.0122 | 2.57E-01 | 0.0138 | 3.23E-01 | -0.0122 | 2.51E-01 | 0.0030 | 6.49E-01 | 8.77E-01 |
| cg20209891 | *MPPED1* | chr22 | 43806614 | -0.0042 | 5.55E-01 | 0.0073 | 3.91E-01 | 0.0034 | 5.71E-01 | 0.0018 | 6.50E-01 | 8.77E-01 |
| cg01542423 | *TRIB1* | chr8 | 126668119 | -0.0114 | 9.08E-02 | 0.0035 | 3.37E-01 | -0.0023 | 4.87E-01 | -0.0010 | 6.57E-01 | 8.83E-01 |
| cg24309215 | *POP5* | chr12 | 121021560 | 0.0045 | 6.02E-01 | 0.0023 | 8.84E-01 | -0.0129 | 1.71E-01 | -0.0026 | 6.60E-01 | 8.83E-01 |
| cg13558105 | *C10orf71* | chr10 | 50530868 | -0.0105 | 6.88E-02 | -0.0007 | 8.90E-01 | 0.0108 | 2.22E-02 | 0.0013 | 6.66E-01 | 8.83E-01 |
| cg02086125 | *LIAS* | chr4 | 39463608 | -0.0087 | 2.07E-01 | -0.0018 | 4.37E-01 | 0.0067 | 1.53E-01 | -0.0008 | 6.71E-01 | 8.83E-01 |
| cg05053718 | *ZNF469* | chr16 | 88313910 | 0.0035 | 5.41E-01 | -0.0019 | 2.68E-01 | 0.0044 | 2.80E-01 | -0.0006 | 6.72E-01 | 8.83E-01 |
| cg09683730 | *USP48* | chr1 | 22013127 | -0.0043 | 3.82E-01 | 0.0008 | 6.29E-01 | -0.0016 | 3.83E-01 | -0.0005 | 6.73E-01 | 8.83E-01 |
| cg00513205 | *NEUROG1* | chr5 | 134872649 | 0.0130 | 3.60E-01 | -0.0050 | 6.36E-01 | 0.0162 | 4.58E-01 | 0.0033 | 6.75E-01 | 8.83E-01 |
| cg07399532 | *MSRA* | chr8 | 10001612 | -0.0060 | 3.32E-01 | 0.0032 | 1.74E-01 | -0.0037 | 3.77E-01 | 0.0008 | 6.75E-01 | 8.83E-01 |
| cg19463078 | *TNIP3* | chr4 | 122099598 | 0.0091 | 1.95E-01 | 0.0049 | 3.14E-01 | -0.0056 | 2.28E-01 | 0.0012 | 6.83E-01 | 8.85E-01 |
| cg20872252 | *LOC283177* | chr11 | 134287326 | -0.0167 | 5.07E-02 | 0.0060 | 2.77E-01 | -0.0025 | 6.41E-01 | -0.0014 | 6.86E-01 | 8.85E-01 |
| cg05812430 | *INTS1* | chr7 | 1542218 | -0.0023 | 5.90E-01 | -0.0002 | 9.06E-01 | 0.0073 | 7.56E-02 | 0.0006 | 6.92E-01 | 8.85E-01 |
| cg14012365 | *FAM110B* | chr8 | 59059187 | -0.0033 | 4.87E-01 | 0.0007 | 8.17E-01 | 0.0068 | 2.09E-01 | 0.0009 | 7.01E-01 | 8.85E-01 |
| cg05623411 | *ZDHHC21* | chr9 | 14691289 | -0.0198 | 2.08E-01 | -0.0045 | 4.24E-01 | 0.0194 | 9.72E-02 | -0.0019 | 7.01E-01 | 8.85E-01 |
| cg11893488 | *COL4A2* | chr13 | 111144464 | -0.0008 | 8.62E-01 | -0.0017 | 6.75E-01 | -0.0004 | 8.78E-01 | -0.0007 | 7.03E-01 | 8.85E-01 |
| cg07349890 | *RRP7A* | chr22 | 42913926 | -0.0042 | 3.64E-01 | -0.0014 | 5.86E-01 | 0.0015 | 5.94E-01 | -0.0007 | 7.07E-01 | 8.85E-01 |
| cg18448949 | *HOXD8* | chr2 | 176993089 | 0.0035 | 4.68E-01 | 0.0006 | 9.33E-01 | -0.0047 | 5.90E-01 | 0.0013 | 7.11E-01 | 8.85E-01 |
| cg06596711 | *ARMC6* | chr19 | 19148759 | -0.0061 | 2.92E-01 | -0.0073 | 2.45E-01 | 0.0048 | 1.15E-01 | 0.0009 | 7.11E-01 | 8.85E-01 |
| cg16660891 | *USP10* | chr16 | 84828913 | -0.0002 | 9.76E-01 | 0.0104 | 2.41E-01 | -0.0010 | 8.60E-01 | 0.0014 | 7.13E-01 | 8.85E-01 |
| cg25162794 | *ANKRD23* | chr2 | 97509831 | -0.0054 | 3.27E-01 | 0.0006 | 7.31E-01 | 0.0036 | 3.74E-01 | 0.0006 | 7.15E-01 | 8.85E-01 |
| cg18961589 | *LINC01164* | chr10 | 133598669 | -0.0096 | 1.60E-01 | 0.0007 | 6.43E-01 | 0.0021 | 5.67E-01 | 0.0005 | 7.15E-01 | 8.85E-01 |
| cg11313335 | *TAP1* | chr6 | 32814921 | 0.0007 | 8.90E-01 | -0.0003 | 8.10E-01 | 0.0044 | 1.59E-01 | 0.0004 | 7.18E-01 | 8.85E-01 |
| cg16594906 | *CACNG4* | chr17 | 65026907 | 0.0018 | 7.63E-01 | -0.0010 | 6.99E-01 | -0.0012 | 7.64E-01 | -0.0007 | 7.19E-01 | 8.85E-01 |
| cg08530934 | *PTPRN2* | chr7 | 158236160 | -0.0113 | 5.56E-02 | -0.0006 | 7.41E-01 | 0.0036 | 3.16E-01 | -0.0005 | 7.20E-01 | 8.85E-01 |
| cg08649360 | *TADA3* | chr3 | 9833841 | -0.0019 | 7.57E-01 | 0.0059 | 5.31E-01 | 0.0051 | 5.76E-01 | 0.0016 | 7.21E-01 | 8.85E-01 |

| | | | | | | | | | | | | |
|---|---|---|---|---|---|---|---|---|---|---|---|---|
| cg02323550 | *PTPRN2* | chr7 | 157660321 | -0.0065 | 1.53E-01 | -0.0003 | 8.91E-01 | 0.0032 | 1.61E-01 | 0.0005 | 7.22E-01 | 8.85E-01 |
| cg21985862 | *THBS2* | chr6 | 169559628 | 0.0009 | 8.51E-01 | -0.0034 | 1.71E-01 | 0.0037 | 2.65E-01 | -0.0006 | 7.33E-01 | 8.92E-01 |
| cg19774899 | *DLK1* | chr14 | 101203896 | 0.0030 | 5.94E-01 | -0.0004 | 7.47E-01 | -0.0021 | 6.05E-01 | -0.0004 | 7.35E-01 | 8.92E-01 |
| cg01134274 | *CLK2* | chr1 | 155239493 | -0.0057 | 2.61E-01 | -0.0004 | 7.61E-01 | 0.0067 | 2.93E-02 | 0.0004 | 7.37E-01 | 8.92E-01 |
| cg02563137 | *PCDHGA1* | chr5 | 140778546 | 0.0023 | 7.68E-01 | -0.0045 | 7.32E-01 | -0.0167 | 2.95E-01 | -0.0021 | 7.38E-01 | 8.92E-01 |
| cg19521311 | *F13A1* | chr6 | 6322347 | -0.0089 | 1.85E-01 | 0.0011 | 4.89E-01 | -0.0001 | 9.86E-01 | 0.0005 | 7.44E-01 | 8.96E-01 |
| cg14078070 | *RPS6KA2* | chr6 | 167051518 | 0.0015 | 7.99E-01 | -0.0021 | 4.78E-01 | 0.0014 | 7.62E-01 | -0.0007 | 7.59E-01 | 9.11E-01 |
| cg24687432 | *ATP10A* | chr15 | 26109608 | -0.0027 | 7.24E-01 | 0.0041 | 5.89E-01 | -0.0054 | 4.77E-01 | -0.0013 | 7.68E-01 | 9.12E-01 |
| cg15159017 | *GALNT9* | chr12 | 132854526 | 0.0028 | 6.92E-01 | 0.0002 | 9.79E-01 | 0.0005 | 9.05E-01 | 0.0010 | 7.68E-01 | 9.12E-01 |
| cg18703983 | *KCNS3* | chr2 | 18097337 | 0.0131 | 1.11E-01 | -0.0014 | 5.91E-01 | 0.0024 | 5.54E-01 | 0.0006 | 7.71E-01 | 9.12E-01 |
| cg22361604 | *NFIA* | chr1 | 61435148 | -0.0092 | 5.01E-01 | -0.0099 | 3.23E-01 | 0.0118 | 1.24E-01 | 0.0016 | 7.72E-01 | 9.12E-01 |
| cg16244889 | *PRIMA1* | chr14 | 94211818 | 0.0021 | 7.97E-01 | -0.0010 | 8.97E-01 | 0.0043 | 6.67E-01 | 0.0014 | 7.73E-01 | 9.12E-01 |
| cg25799241 | *FAM83H* | chr8 | 144805740 | 0.0027 | 7.38E-01 | -0.0021 | 5.07E-01 | 0.0025 | 6.86E-01 | -0.0007 | 7.85E-01 | 9.17E-01 |
| cg09582880 | *IL17A* | chr6 | 52050133 | -0.0022 | 6.42E-01 | -0.0003 | 8.22E-01 | 0.0002 | 9.47E-01 | -0.0003 | 7.88E-01 | 9.17E-01 |
| cg09632585 | *ZNF131* | chr5 | 43125540 | -0.0174 | 7.93E-03 | 0.0022 | 4.81E-01 | 0.0016 | 7.33E-01 | -0.0006 | 7.95E-01 | 9.17E-01 |
| cg03365705 | *GREB1* | chr2 | 11751071 | -0.0080 | 1.26E-01 | 0.0000 | 9.93E-01 | 0.0066 | 9.15E-02 | 0.0005 | 7.96E-01 | 9.17E-01 |
| cg27449651 | *SYNGAP1* | chr6 | 33395433 | 0.0073 | 3.24E-01 | 0.0002 | 9.63E-01 | -0.0067 | 2.20E-01 | -0.0008 | 7.98E-01 | 9.17E-01 |
| cg27062927 | *USH1C* | chr11 | 17555142 | -0.0104 | 3.96E-02 | -0.0017 | 8.02E-01 | 0.0055 | 8.25E-02 | 0.0006 | 7.99E-01 | 9.17E-01 |
| cg19827716 | *GM140* | chr1 | 181287967 | -0.0025 | 7.34E-01 | -0.0050 | 3.48E-01 | 0.0071 | 2.98E-01 | -0.0009 | 8.00E-01 | 9.17E-01 |
| cg18298197 | *MRPS18B* | chr6 | 30593314 | 0.0012 | 8.03E-01 | -0.0002 | 9.42E-01 | -0.0010 | 6.80E-01 | -0.0005 | 8.01E-01 | 9.17E-01 |
| cg11429111 | *TIFAB* | chr5 | 134813329 | 0.0013 | 8.22E-01 | -0.0008 | 9.46E-01 | 0.0015 | 8.46E-01 | 0.0010 | 8.03E-01 | 9.17E-01 |
| cg00513781 | *THAP4* | chr2 | 242571652 | -0.0011 | 8.67E-01 | -0.0012 | 3.88E-01 | 0.0042 | 1.59E-01 | -0.0003 | 8.08E-01 | 9.17E-01 |
| cg22435335 | *RTP5* | chr2 | 242841392 | -0.0058 | 3.51E-01 | 0.0023 | 3.34E-01 | -0.0023 | 5.80E-01 | 0.0005 | 8.08E-01 | 9.17E-01 |
| cg16525987 | *TEX26-AS1* | chr13 | 31506376 | 0.0098 | 3.10E-01 | -0.0025 | 2.56E-01 | 0.0140 | 5.74E-03 | 0.0005 | 8.15E-01 | 9.22E-01 |
| cg05149097 | *OSBP* | chr11 | 59325534 | 0.0015 | 8.83E-01 | 0.0078 | 4.17E-01 | -0.0033 | 6.66E-01 | 0.0012 | 8.22E-01 | 9.27E-01 |
| cg08726801 | *CACNA2D4* | chr12 | 1974122 | -0.0019 | 8.28E-01 | -0.0132 | 4.13E-01 | 0.0016 | 8.20E-01 | -0.0011 | 8.27E-01 | 9.29E-01 |
| cg01281501 | *ADGRD1* | chr12 | 131494451 | -0.0064 | 2.76E-01 | -0.0004 | 9.54E-01 | 0.0061 | 3.61E-01 | -0.0008 | 8.30E-01 | 9.29E-01 |
| cg26529239 | *ZBED9* | chr6 | 28550343 | -0.0290 | 4.79E-03 | -0.0003 | 9.08E-01 | 0.0077 | 1.85E-01 | -0.0005 | 8.35E-01 | 9.32E-01 |
| cg20926049 | *DLL1* | chr6 | 170591749 | -0.0103 | 4.81E-02 | 0.0010 | 5.85E-01 | 0.0029 | 4.34E-01 | 0.0003 | 8.39E-01 | 9.34E-01 |
| cg03050981 | *LEPR* | chr1 | 65906670 | -0.0047 | 6.93E-01 | -0.0080 | 5.61E-01 | 0.0088 | 5.11E-01 | -0.0015 | 8.45E-01 | 9.37E-01 |
| cg10518144 | *NUF2* | chr1 | 163581520 | -0.0085 | 6.99E-02 | 0.0054 | 6.39E-02 | -0.0014 | 6.37E-01 | 0.0004 | 8.49E-01 | 9.37E-01 |
| cg05276226 | *ADAM18* | chr8 | 39441968 | -0.0205 | 5.52E-02 | 0.0017 | 6.03E-01 | 0.0021 | 6.70E-01 | 0.0005 | 8.53E-01 | 9.37E-01 |
| cg08027748 | *UROC1* | chr3 | 126237319 | -0.0027 | 5.24E-01 | 0.0009 | 5.12E-01 | -0.0029 | 4.44E-01 | 0.0002 | 8.53E-01 | 9.37E-01 |
| cg08343671 | *SNORD115-7* | chr15 | 25427509 | -0.0028 | 7.05E-01 | 0.0095 | 5.13E-01 | 0.0011 | 8.14E-01 | 0.0007 | 8.61E-01 | 9.43E-01 |
| cg18011253 | *ISLR2* | chr15 | 74424054 | 0.0009 | 8.16E-01 | 0.0049 | 4.45E-01 | -0.0076 | 3.28E-01 | 0.0005 | 8.67E-01 | 9.45E-01 |
| cg11071155 | *ARID1B* | chr6 | 157012049 | 0.0020 | 7.08E-01 | -0.0002 | 9.35E-01 | 0.0006 | 8.79E-01 | 0.0003 | 8.71E-01 | 9.45E-01 |
| cg03922126 | *KCNQ5* | chr6 | 73330297 | 0.0079 | 4.27E-01 | -0.0002 | 9.60E-01 | -0.0050 | 4.44E-01 | -0.0005 | 8.72E-01 | 9.45E-01 |
| cg01522083 | *TSR3* | chr16 | 1400861 | -0.0024 | 7.39E-01 | 0.0061 | 1.95E-01 | -0.0073 | 1.52E-01 | -0.0005 | 8.78E-01 | 9.46E-01 |
| cg26832915 | *FAM53A* | chr4 | 1577932 | -0.0204 | 2.08E-03 | 0.0025 | 4.05E-01 | 0.0032 | 5.41E-01 | -0.0004 | 8.81E-01 | 9.46E-01 |
| cg27639662 | *PRDM8* | chr4 | 81111393 | -0.0128 | 2.72E-01 | 0.0283 | 1.69E-01 | 0.0110 | 4.88E-01 | 0.0013 | 8.81E-01 | 9.46E-01 |
| cg17491365 | *TMEM232* | chr5 | 109962391 | -0.0104 | 1.36E-01 | 0.0003 | 8.85E-01 | 0.0115 | 1.15E-01 | 0.0003 | 8.88E-01 | 9.51E-01 |
| cg00029821 | *STXBP5-AS1* | chr6 | 147374493 | -0.0225 | 1.33E-02 | 0.0008 | 7.00E-01 | 0.0053 | 3.80E-01 | 0.0002 | 8.97E-01 | 9.55E-01 |
| cg21799053 | *RHOBTB2* | chr8 | 22856801 | 0.0032 | 5.16E-01 | 0.0009 | 7.05E-01 | -0.0024 | 4.60E-01 | 0.0002 | 8.98E-01 | 9.55E-01 |
| cg01974817 | *PTPRU* | chr1 | 29802252 | -0.0217 | 9.75E-03 | 0.0015 | 6.94E-01 | 0.0050 | 3.63E-01 | -0.0004 | 9.03E-01 | 9.55E-01 |
| cg16506432 | *CDC20B* | chr5 | 54428863 | -0.0112 | 4.28E-02 | 0.0001 | 9.25E-01 | 0.0033 | 2.52E-01 | 0.0001 | 9.05E-01 | 9.55E-01 |
| cg13580380 | *MSC* | chr8 | 72667937 | 0.0009 | 8.63E-01 | 0.0049 | 2.98E-01 | -0.0131 | 9.03E-02 | 0.0004 | 9.07E-01 | 9.55E-01 |
| cg00116430 | *BCAR3* | chr1 | 94188268 | -0.0021 | 8.52E-01 | -0.0001 | 9.92E-01 | 0.0005 | 9.77E-01 | -0.0008 | 9.12E-01 | 9.55E-01 |
| cg10510558 | *MTX3* | chr5 | 79290786 | -0.0174 | 3.21E-03 | 0.0002 | 9.40E-01 | 0.0044 | 1.26E-01 | 0.0002 | 9.12E-01 | 9.55E-01 |
| cg15599483 | *SLC40A1* | chr2 | 190442823 | -0.0061 | 4.30E-01 | -0.0003 | 9.60E-01 | 0.0052 | 5.04E-01 | -0.0005 | 9.15E-01 | 9.55E-01 |
| cg09154062 | *WNT4* | chr1 | 22582789 | 0.0076 | 3.61E-01 | -0.0012 | 7.77E-01 | -0.0035 | 6.10E-01 | -0.0003 | 9.19E-01 | 9.56E-01 |
| cg27505745 | *UBE2MP1* | chr16 | 34411816 | -0.0026 | 7.30E-01 | 0.0079 | 3.14E-01 | -0.0054 | 4.50E-01 | -0.0004 | 9.24E-01 | 9.58E-01 |
| cg05487105 | *CLDN18* | chr3 | 137729296 | -0.0026 | 5.40E-01 | 0.0003 | 8.40E-01 | -0.0006 | 8.09E-01 | -0.0001 | 9.33E-01 | 9.65E-01 |
| cg07612655 | *PTGIS* | chr20 | 48185517 | 0.0002 | 9.65E-01 | 0.0100 | 4.42E-01 | -0.0065 | 4.65E-01 | -0.0003 | 9.46E-01 | 9.73E-01 |
| cg26571002 | *SIGLEC15* | chr18 | 43416018 | -0.0113 | 1.27E-01 | 0.0029 | 3.93E-01 | -0.0005 | 9.23E-01 | 0.0002 | 9.49E-01 | 9.73E-01 |
| cg02140508 | *RAD51AP1* | chr12 | 4659038 | -0.0078 | 3.33E-01 | -0.0002 | 9.70E-01 | 0.0040 | 5.09E-01 | -0.0002 | 9.50E-01 | 9.73E-01 |
| cg24837397 | *CUX1* | chr7 | 101632568 | -0.0018 | 5.99E-01 | 0.0016 | 6.07E-01 | 0.0002 | 9.60E-01 | 0.0001 | 9.53E-01 | 9.73E-01 |
| cg00612828 | *DCTD* | chr4 | 183815971 | -0.0055 | 2.61E-01 | -0.0003 | 8.52E-01 | 0.0033 | 3.03E-01 | -0.0001 | 9.57E-01 | 9.75E-01 |
| cg19166759 | *IGF2R* | chr6 | 160501174 | 0.0014 | 8.24E-01 | -0.0064 | 5.12E-01 | 0.0001 | 9.69E-01 | -0.0001 | 9.63E-01 | 9.77E-01 |
| cg03455964 | *PATE1* | chr11 | 125615317 | -0.0023 | 6.53E-01 | 0.0007 | 7.85E-01 | -0.0004 | 9.22E-01 | -0.0001 | 9.79E-01 | 9.86E-01 |
| cg04124888 | *CAPZB* | chr1 | 19788288 | -0.0052 | 4.04E-01 | -0.0012 | 4.77E-01 | 0.0039 | 1.33E-01 | 0.0000 | 9.82E-01 | 9.86E-01 |
| cg27078729 | *TNRC6A* | chr16 | 24835194 | -0.0034 | 5.49E-01 | -0.0001 | 9.40E-01 | 0.0021 | 5.65E-01 | 0.0000 | 9.82E-01 | 9.86E-01 |
| cg18666249 | *FOXN3* | chr14 | 89670502 | -0.0079 | 4.60E-01 | -0.0014 | 9.16E-01 | 0.0060 | 4.89E-01 | 0.0001 | 9.83E-01 | 9.86E-01 |
| cg26154235 | *PDZRN3* | chr3 | 73622059 | -0.0083 | 2.75E-01 | 0.0033 | 2.16E-01 | -0.0071 | 1.25E-01 | 0.0000 | 9.89E-01 | 9.89E-01 |

**Table S4.** Nominally significant *cis*-acting eQTMs for the 12 CpG sites associated with atopic asthma (total 182 CpG-gene pairs, from Table 2) in nasal epithelial cells from participants in EVA-PR

| | | EVA-PR | |
|---|---|---|---|
| **CpG** | **Gene** | **Beta** | ***P*-value** |
| cg27178677 | *TMX4* | 0.1078 | 9.29E-04 |
| cg02695349 | *VPS41* | -0.0362 | 6.15E-03 |
| cg18146152 | *NPW* | -0.1232 | 1.09E-02 |
| cg17335499 | *TRIM27* | -0.0383 | 1.55E-02 |
| cg02695349 | *TRG-AS1* | -0.0959 | 1.57E-02 |
| cg03541903 | *HINT1* | -0.0304 | 1.95E-02 |
| cg18146152 | *NME4* | -0.0778 | 2.32E-02 |
| cg17335499 | *ZKSCAN8* | 0.0454 | 2.46E-02 |
| cg17335499 | *HLA-F* | 0.1270 | 2.54E-02 |
| cg02695349 | *STARD3NL* | -0.0365 | 3.05E-02 |
| cg18146152 | *RPS2* | -0.0523 | 3.06E-02 |
| cg18146152 | *TMEM8A* | 0.0598 | 3.76E-02 |
| cg01039401 | *RHNO1* | 0.0400 | 4.32E-02 |
| cg01039401 | *DCP1B* | 0.0427 | 4.44E-02 |
| cg17335499 | *ZNF311* | 0.0621 | 4.45E-02 |
| cg02695349 | *TARP* | -0.1192 | 4.88E-02 |
